# Reduced amplification efficiency of the RNA-dependent-RNA-polymerase (RdRp) target enables tracking of the Delta SARS-CoV-2 variant using routine diagnostic tests

**DOI:** 10.1101/2021.10.01.21264408

**Authors:** Ziyaad Valley-Omar, Gert Marais, Arash Iranzadeh, Michelle Naidoo, Stephen Korsman, Tongai Maponga, Hannah Hussey, Mary-Ann Davies, Andrew Boulle, Deelan Doolabh, Mariska Laubscher, JD Deetlefs, Jean Maritz, Lesley Scott, Nokukhanya Msomi, Houriiyah Tegally, Tulio de Oliveira, Jinal Bhiman, Carolyn Williamson, Wolfgang Preiser, Diana Hardie, Nei-yuan Hsiao

## Abstract

Routine SARS-CoV-2 surveillance in the Western Cape region of South Africa (January-August 2021) found a reduced PCR amplification efficiency of the RdRp gene target of the Seegene, Allplex 2019-nCoV diagnostic assay when detecting the Delta variant. We propose that this can be used as a surrogate for variant detection.

## The Study

Genomic surveillance of the severe acute respiratory syndrome coronavirus 2 (SARS-CoV-2) by whole genome sequencing (WGS) has played a critical role in identifying and monitoring the dissemination of variants of concern (VOCs) [1, 2]. However, WGS is costly and time-consuming, and not all countries have the resources to do this at scale to get detailed VOC epidemiological information.

Multiplex PCR-based diagnostic tests for SARS-CoV-2, which simultaneously amplify and detect multiple SARS-CoV-2 gene targets, such as the Spike (S)-, Nucleocapsid (N)-, Envelope (E)- and RNA-dependent-RNA-polymerase (RdRp) genes, are the primary tools used to define cases and epidemic waves. Current commercial tests were designed to detect ancestral SARS-CoV-2 sequences and have not been optimised for the viral genomic diversity that has subsequently emerged [3]. Despite this, built-in assay redundancy when detecting multiple gene targets, these tests continue to be an effective tool for diagnosing infections by contemporary variants. For example, deletions of amino acids 69 and 70 within the S-gene of SARS-CoV-2 Alpha variants was associated with failure to detect this target by some commonly used commercial diagnostic assays (e.g. ThermoFisher TaqPath) [4]. Fortuitously, because the other targets are largely unaffected, this S-gene target failure (S-GTF) serves as a surrogate marker for monitoring prevalence of SARS-CoV-2 Alpha variant [4, 5]. As the Delta variant becomes the predominant VOC across the world, a similar diagnostic PCR-based marker has potential to enable similar rapid epidemiological assessment and research. An evaluation of routine diagnostic results together with WGS, we show that infection with the Delta variant is associated with reduced RdRp target amplification efficiency and propose that this can be used for tracking the prevalence of this variant.

Routine SARS-CoV-2 diagnostic testing activities of the non-insured population in South Africa is performed by the National Health Laboratory Service (NHLS). These laboratories also form part of the network of genomic surveillance of South Africa (NGS-SA) where a subset of routine diagnostic samples are sequenced as part of the genomic surveillance activities [6]. WGS was performed using either GridION X5 (Oxford Nanopore Technologies) or MiSeq (Illumina) sequencing platforms [7]. To ensure accurate clade assignment, we only included sequences with less than 3000 bases missing, no frameshift mutations and no misplaced stop codons. All sequences were deposited in real time on the GISAID SARS-CoV-2 sequence repository (https://www.gisaid.org/) (Supplementary Table 1).

Between March 2020 and August 2021, the Seegene Allplex 2019-nCoV assay, a single-tube multiplex assay with 3 SARS-CoV-2 gene targets: E, RdRp and N, was one of the most commonly used RT-PCR platforms across the NHLS. From May 2021, we observed an increase in the number of samples testing positive with the Seegene Allplex nCoV-19 assay that had either delayed or absent RdRp target detection (cycle threshold (Ct) value) relative to E gene and N gene targets. Prior to this period, the average difference between the Ct values for RdRp and E-gene targets (RΔE) was around 2. From June 2021, the RΔE in diagnostic samples increased to a median of 5 (Figure 1). We investigated if the efficiency of RdRP target amplification and detection was affected by mutations in the predominant circulating variant.

**Figure 1.**
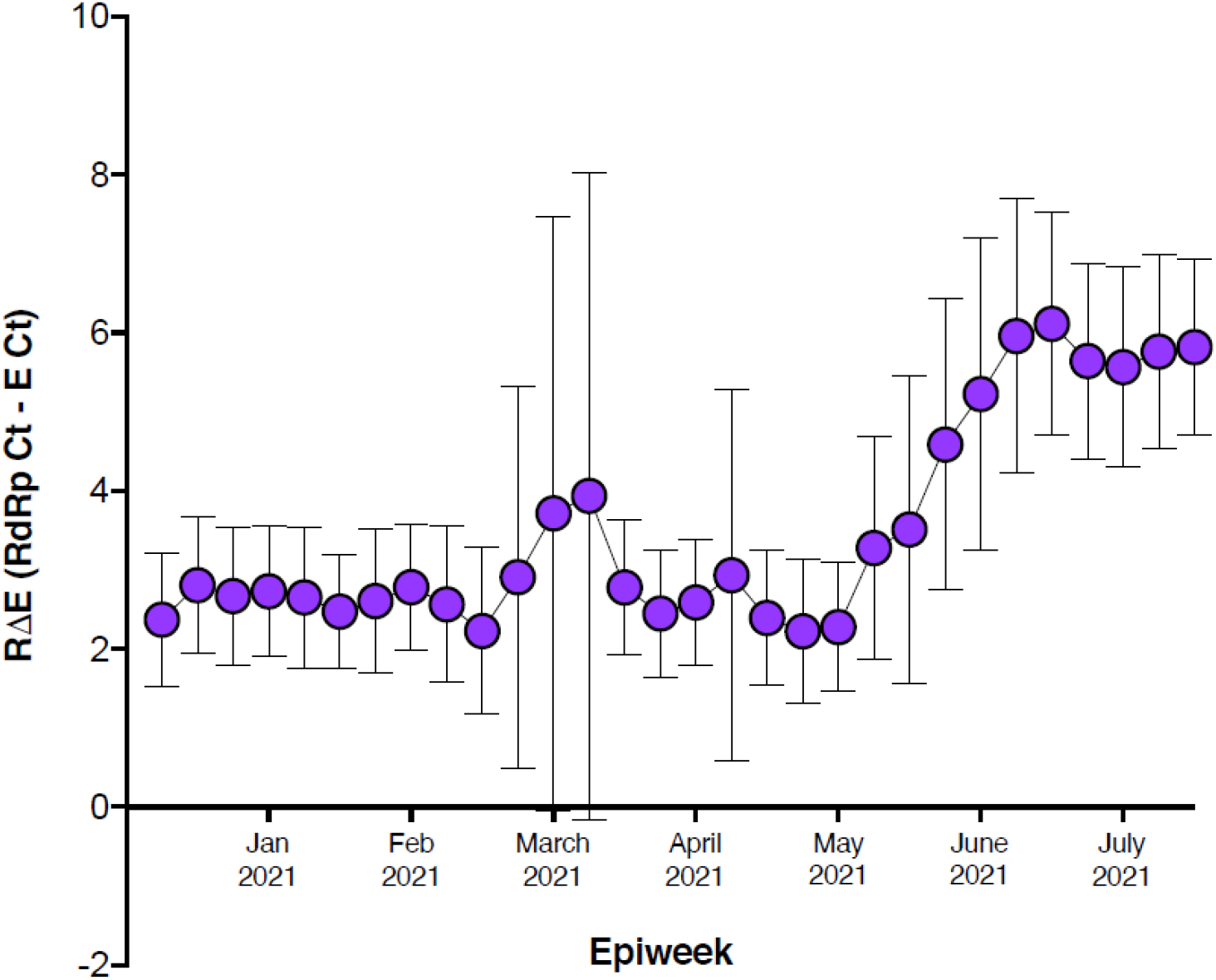
Mean ΔCt by week and genomic surveillance data. The mean Δ Ct (Seegene Allplex 2019-nCoV assay RdRp gene Ct value minus E gene value) is shown for samples testing positive for both E and RdRp targets for each week of 2021 up to 31 July. Only samples tested in the Western Cape region of South Africa are included in the analysis. Error bars represent 1 standard deviation.

In Western Cape province where the study was conducted, the region entered its third wave on 21 June 2021 (week 25) where more than 1020 cases had been reported daily and there was >20% week on week increase in the 7-day moving average of new cases. During the same period, genomic surveillance data showed a rapid increase in Delta variant, which (Figure 2) replaced Beta as the predominant VOC in the region. As the cases surged, the proportion of Delta variant increased from just over 20% of total samples sequenced in May 2021, to >90% in August 2021.

**Figure 2.**
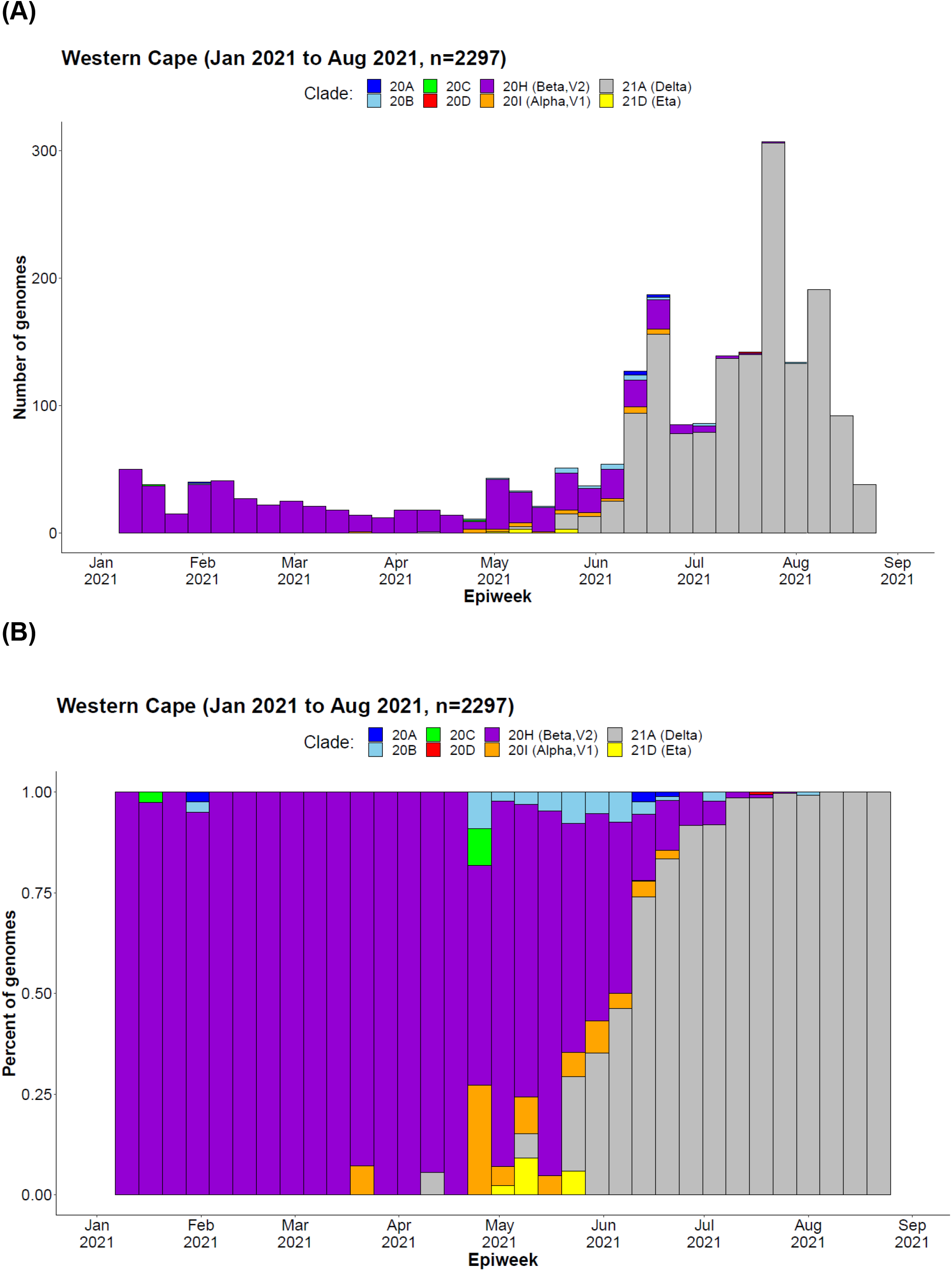
Weekly frequency and distribution of SARS-CoV-2 variants circulating in the Western Cape region of South Africa between 1 January and 31 August 2021. (A) Absolute count of genomes sequenced, (B) Proportion of genomes sequenced.

To investigate the observed RdRp target delay which coincided with the emergence of Delta, RdRp gene sequences of samples tested with the Seegene, Allplex 2019-nCoV assay were evaluated for mutations that could interfere with PCR probe or primer binding. We identified a non-synonymous G15451A mutation (codon 671S) within RdRp gene of Delta variants, potentially responsible for the reduced amplification efficiency of the assay RdRp target. This highly conserved G15451A mutation was present in 100% (362/362) of Delta variant sequences and not observed in Alpha (n=9), Beta (n=731) or Eta (n=5) variants sequenced at this time (Supplementary Figure 1). This specific mutation was significant as it resulted in a single nucleotide mismatch in the second last base of the WHO-recommended forward primer binding site for RdRp amplification for most diagnostic assays targeting RdRP, including Seegene Allplex nCoV-19 [8]. In addition, we previously found that a synonymous G15452C mutation (codon 671S) in a minority of Beta variant sequences (3’-end of this forward primer binding site) also affected RdRp gene target detection when using the Seegene, Allplex 2019-nCoV assay (Supplementary Figure 1).

To systematically assess the effect of this Delta variant G15451A mutation on Seegene assay RdRp target amplification efficiency relative to that of ancestral strain and Beta variant, we compared the median relative RdRp and E gene target Ct values (RΔE) among the different Nextstrain clades detected in diagnostic samples over this time period. Of the 1455 sequences retrieved, 11, 749, 369, 329 were Alpha, Beta, Delta and other variants respectively. After removing 39 sequences where target Ct data was incomplete, Delta variant samples had a significantly increased RΔE (n =360, median =5.74, IQR **4.76-6.55**) when compared to non-Delta variant samples (n=1056, median = 2.54, IQR2.13-3, p<0.001) (Supplementary figure 2). The diagnostic ability of the RΔE value to identify the Delta variant was evaluated using a receiver operating characteristic (ROC) curve (Supplementary figure 3). Using a RΔE threshold of > 3.5 cycles we were able to accurately identify delta variant positive samples within the sample set with 93.6% sensitivity and 89.7% specificity, correctly classifying 90% of cases (AUC = 0.9663). When the threshold is increased to 4 and 4.5 cycles, the specificity was improved to 96.5% and 98% respectively but the sensitivity was reduced to 86.7% and 80% respectively.

In this article we show that reduced amplification efficiency of the RdRp gene target of the Seegene, Allplex 2019-nCoV assay can be used as an indirect measure of SARS-CoV-2 Delta variant prevalence in a population. Using the RΔE value could therefore serve as a reliable surrogate for genomic sequencing to approximate the spread of the SARS-CoV-2 Delta variant. While using the RΔE is only a surrogate marker of Delta variant identity, assay Ct values have proven useful in the past to identify Alpha variant prevalence as well as serving as an independent predictor of disease severity [9, 10]. Limitations to our study concern the proprietary nature of the Seegene, Allplex 2019-nCoV assay. We are unable to confirm the genomic loci amplified by the assay primers and probes. Evidence including the RdRp target delay in Delta variants from this study, and RdRp complete GTF in a minority of Beta variants, suggest that these diagnostic primers overlap with the WHO-recommended PCR primer sets stemming from Corman *et al*., 2020 [8]. This study highlights the need for continued monitoring of the efficacy of current commercial SARS-CoV-2 diagnostic assays in a setting where we are observing the constant genetic drift of a novel pathogen.

## Data Availability

All study sequences are listed in the supplementary table

## Acknowledgments

We gratefully acknowledge the efforts of the staff of the NHLS and UCT (Nokuzola Mbhele, Fezokuhle Khumalo, Thabang Serakge and Innocent Mudau) laboratories responsible for obtaining the specimens.

This work was supported by grants from the Wellcome Centre for Infectious Diseases Research in Africa (CIDRI-Africa), Institute of Infectious Disease and Molecular Medicine (core grant number is 203135/Z/16/Z), Division of Medical Virology in Department of Pathology, University of Cape Town, Observatory 7925, Republic of South Africa, the Grand Challenges ICODA pilot initiative delivered by Health Data Research UK and funded by the Bill & Melinda Gates and the Minderoo Foundations, and by a research Flagship grant from the South African Medical Research Council (MRC-RFA-UFSP-01-2013/UKZN HIVEPI).

This study is an extension to current genomic surveillance activity of Network for Genomic Surveillance in South Africa (NGS-SA) and forms part of the “*Molecular Surveillance of SARS-CoV-2 in the Western Cape: Understanding Spread to Inform Intervention Strategies*” study within the UCT Division of Medical Virology. Study protocols have been approved by the UCT Faculty of Health Sciences Human Research Ethics Committee (HREC:383/2020).

