## Supplementary figures for "Reduced amplification efficiency of the RNA-dependent-RNA-polymerase (RdRp) target enables tracking of the Delta SARS-CoV-2 variant using routine diagnostic tests"

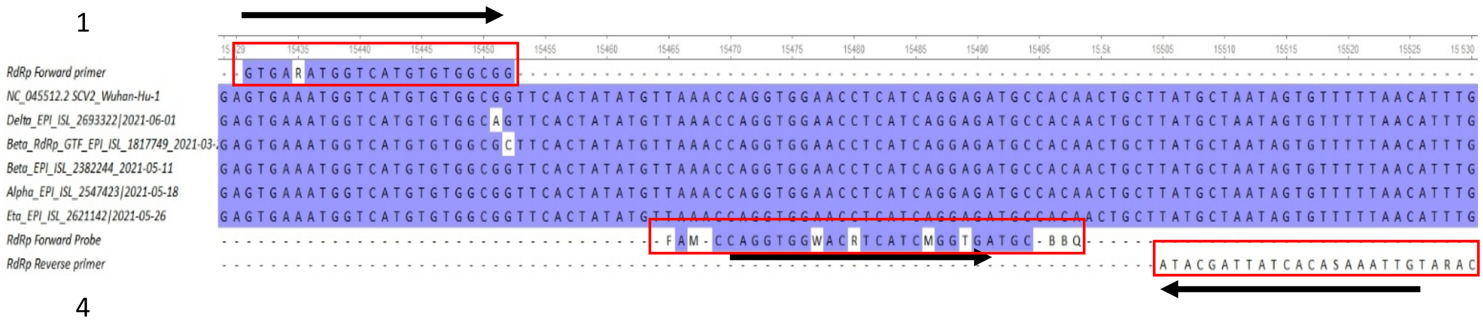


**Supplementary Figure 1.** Nucleic acid alignment showing binding sites of WHO-recommended RdRp PCR primer and probe sets stemming from Corman *et al*., 2020 for contemporary SARS-CoV-2 variants of concern.

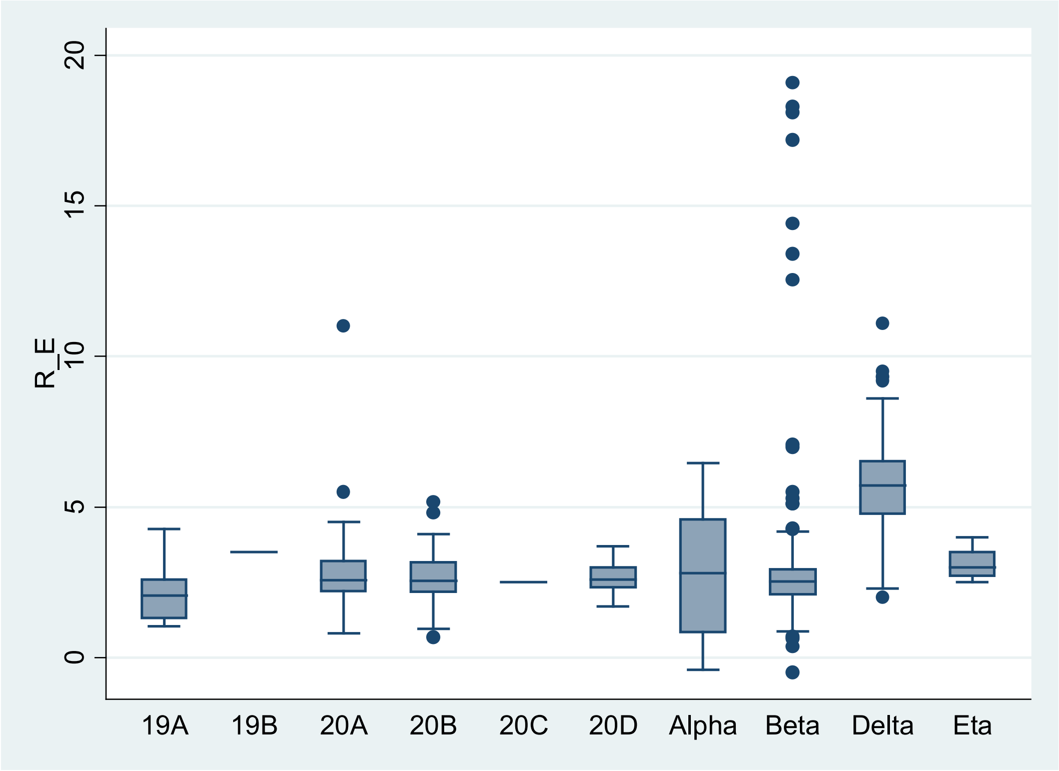


**Supplementary Figure 2.** The relative RdRp Ct (RΔE) of Nextstrain clades. Of the 1416 Western Cape sequences retrieved from GISAID with good coverage (<3000 missing bases) and complete Ct data, the median Ct difference of RdRp-E in samples assigned by Nextstrain as Delta variant was (5.73, IQR4.76-6.53), substantially higher than the medians of the other variants (2.54, IQR 2.13-3, p<0.001). Some of the outliers observed in the beta variant were previous found to have the rare G15451C mutation which result in complete RdRp gene target failure.

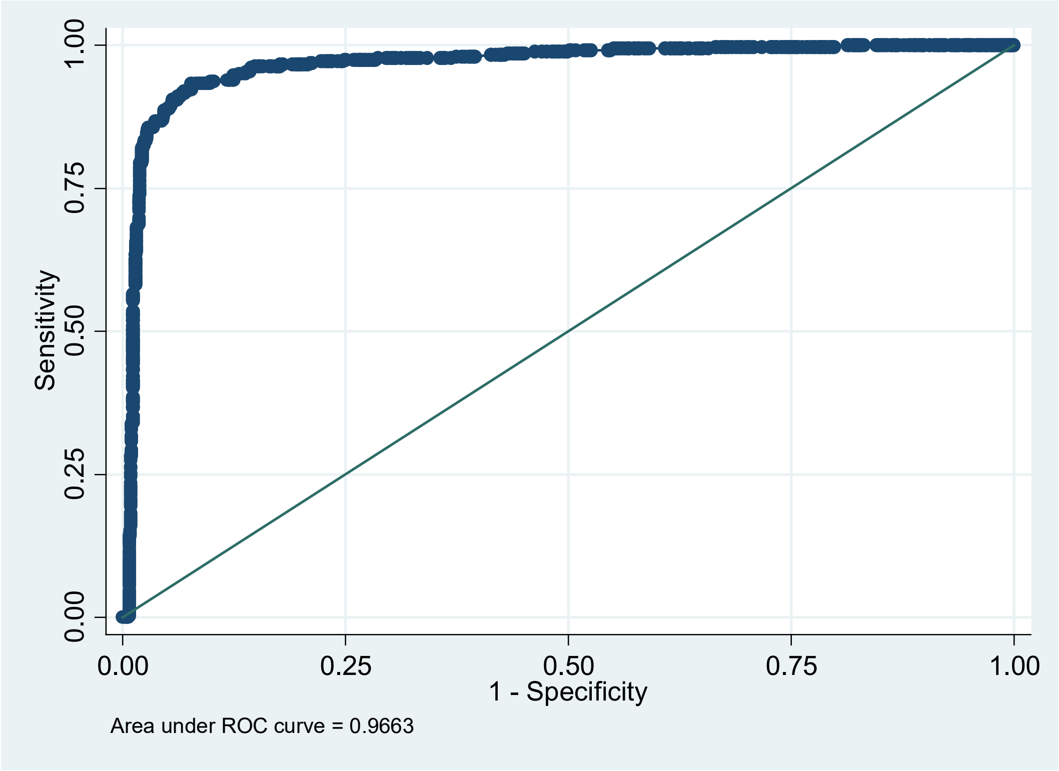


**Supplementary Figure 3.** Receiver operating characteristic (ROC) curve analysis of using relative RdRp target to identify Delta variant in sequence confirmed samples.
