## Supplementary Table 1 for "Reduced amplification efficiency of the RNA-dependent-RNA-polymerase (RdRp) target enables tracking of the Delta SARS-CoV-2 variant using routine diagnostic tests"

| seqName | clade |
|---|---|
| hCoV-19/SouthAfrica/Tygerberg_2113/2021\|EPI_ISL_3847894\|2021-08-11 | 21A (Delta) |
| hCoV-19/SouthAfrica/Tygerberg_2114/2021\|EPI_ISL_3847810\|2021-08-11 | 21A (Delta) |
| hCoV-19/SouthAfrica/Tygerberg_2115/2021\|EPI_ISL_3847897\|2021-08-10 | 21A (Delta) |
| hCoV-19/SouthAfrica/Tygerberg_2116/2021\|EPI_ISL_3847919\|2021-08-10 | 21A (Delta) |
| hCoV-19/SouthAfrica/Tygerberg_2117/2021\|EPI_ISL_3847846\|2021-08-10 | 21A (Delta) |
| hCoV-19/SouthAfrica/Tygerberg_2118/2021\|EPI_ISL_3847901\|2021-08-11 | 21A (Delta) |
| hCoV-19/SouthAfrica/Tygerberg_2119/2021\|EPI_ISL_3847826\|2021-08-11 | 21A (Delta) |
| hCoV-19/SouthAfrica/Tygerberg_2121/2021\|EPI_ISL_3847801\|2021-08-10 | 21A (Delta) |
| hCoV-19/SouthAfrica/Tygerberg_2122/2021\|EPI_ISL_3847904\|2021-08-11 | 21A (Delta) |
| hCoV-19/SouthAfrica/Tygerberg_2123/2021\|EPI_ISL_3847907\|2021-08-11 | 21A (Delta) |
| hCoV-19/SouthAfrica/Tygerberg_2124/2021\|EPI_ISL_3847910\|2021-08-10 | 21A (Delta) |
| hCoV-19/SouthAfrica/Tygerberg_2125/2021\|EPI_ISL_3847804\|2021-08-11 | 21A (Delta) |
| hCoV-19/SouthAfrica/Tygerberg_2126/2021\|EPI_ISL_3847848\|2021-08-10 | 21A (Delta) |
| hCoV-19/SouthAfrica/Tygerberg_2127/2021\|EPI_ISL_3847783\|2021-08-11 | 21A (Delta) |
| hCoV-19/SouthAfrica/Tygerberg_2129/2021\|EPI_ISL_3847786\|2021-08-11 | 21A (Delta) |
| hCoV-19/SouthAfrica/Tygerberg_2130/2021\|EPI_ISL_3847850\|2021-08-10 | 21A (Delta) |
| hCoV-19/SouthAfrica/Tygerberg_2131/2021\|EPI_ISL_3847756\|2021-08-11 | 21A (Delta) |
| hCoV-19/SouthAfrica/Tygerberg_2132/2021\|EPI_ISL_3847852\|2021-08-10 | 21A (Delta) |
| hCoV-19/SouthAfrica/Tygerberg_2133/2021\|EPI_ISL_3847837\|2021-08-11 | 21A (Delta) |
| hCoV-19/SouthAfrica/Tygerberg_2134/2021\|EPI_ISL_3847854\|2021-08-10 | 21A (Delta) |
| hCoV-19/SouthAfrica/Tygerberg_2135/2021\|EPI_ISL_3847841\|2021-08-11 | 21A (Delta) |
| hCoV-19/SouthAfrica/Tygerberg_2136/2021\|EPI_ISL_3847768\|2021-08-10 | 21A (Delta) |
| hCoV-19/SouthAfrica/Tygerberg_2137/2021\|EPI_ISL_3847723\|2021-08-11 | 21A (Delta) |
| hCoV-19/SouthAfrica/Tygerberg_2138/2021\|EPI_ISL_3847733\|2021-08-11 | 21A (Delta) |
| hCoV-19/SouthAfrica/Tygerberg_2139/2021\|EPI_ISL_3847856\|2021-08-10 | 21A (Delta) |
| hCoV-19/SouthAfrica/Tygerberg_2140/2021\|EPI_ISL_3847915\|2021-08-11 | 21A (Delta) |
| hCoV-19/SouthAfrica/Tygerberg_2141/2021\|EPI_ISL_3847828\|2021-08-10 | 21A (Delta) |
| hCoV-19/SouthAfrica/Tygerberg_2142/2021\|EPI_ISL_3847871\|2021-08-10 | 21A (Delta) |
| hCoV-19/SouthAfrica/Tygerberg_2143/2021\|EPI_ISL_3847749\|2021-08-10 | 21A (Delta) |
| hCoV-19/SouthAfrica/Tygerberg_2145/2021\|EPI_ISL_3847748\|2021-08-10 | 21A (Delta) |
| hCoV-19/SouthAfrica/Tygerberg_2148/2021\|EPI_ISL_3847867\|2021-08-10 | 21A (Delta) |
| hCoV-19/SouthAfrica/Tygerberg_2150/2021\|EPI_ISL_3847780\|2021-08-10 | 21A (Delta) |
| hCoV-19/SouthAfrica/Tygerberg_2151/2021\|EPI_ISL_3847816\|2021-08-10 | 21A (Delta) |
| hCoV-19/SouthAfrica/Tygerberg_2153/2021\|EPI_ISL_3847869\|2021-08-10 | 21A (Delta) |
| hCoV-19/SouthAfrica/Tygerberg_2155/2021\|EPI_ISL_3847789\|2021-08-11 | 21A (Delta) |
| hCoV-19/SouthAfrica/Tygerberg_2156/2021\|EPI_ISL_3847771\|2021-08-10 | 21A (Delta) |
| hCoV-19/SouthAfrica/Tygerberg_2157/2021\|EPI_ISL_3847845\|2021-08-11 | 21A (Delta) |
| hCoV-19/SouthAfrica/Tygerberg_2158/2021\|EPI_ISL_3847742\|2021-08-10 | 21A (Delta) |
| hCoV-19/SouthAfrica/Tygerberg_2159/2021\|EPI_ISL_3847823\|2021-08-11 | 21A (Delta) |
| hCoV-19/SouthAfrica/Tygerberg_2161/2021\|EPI_ISL_3847830\|2021-08-11 | 21A (Delta) |
| hCoV-19/SouthAfrica/Tygerberg_2162/2021\|EPI_ISL_3847858\|2021-08-10 | 21A (Delta) |
| hCoV-19/SouthAfrica/Tygerberg_2163/2021\|EPI_ISL_3847739\|2021-08-10 | 21A (Delta) |
| hCoV-19/SouthAfrica/Tygerberg_2164/2021\|EPI_ISL_3847753\|2021-08-10 | 21A (Delta) |
| hCoV-19/SouthAfrica/Tygerberg_2165/2021\|EPI_ISL_3847873\|2021-08-11 | 21A (Delta) |
| hCoV-19/SouthAfrica/Tygerberg_2166/2021\|EPI_ISL_3847874\|2021-08-10 | 21A (Delta) |
| hCoV-19/SouthAfrica/Tygerberg_2167/2021\|EPI_ISL_3847754\|2021-08-10 | 21A (Delta) |
| hCoV-19/SouthAfrica/Tygerberg_2169/2021\|EPI_ISL_3847876\|2021-08-10 | 21A (Delta) |
| hCoV-19/SouthAfrica/Tygerberg_2170/2021\|EPI_ISL_3847774\|2021-08-11 | 21A (Delta) |
| hCoV-19/SouthAfrica/Tygerberg_2171/2021\|EPI_ISL_3847813\|2021-08-10 | 21A (Delta) |

| | |
|---|---|
| hCoV-19/SouthAfrica/Tygerberg_2172/2021\|EPI_ISL_3847807\|2021-08-10 | 21A (Delta) |
| hCoV-19/SouthAfrica/Tygerberg_2173/2021\|EPI_ISL_3847933\|2021-08-10 | 21A (Delta) |
| hCoV-19/SouthAfrica/Tygerberg_2174/2021\|EPI_ISL_3847880\|2021-08-10 | 21A (Delta) |
| hCoV-19/SouthAfrica/Tygerberg_2175/2021\|EPI_ISL_3847883\|2021-08-09 | 21A (Delta) |
| hCoV-19/SouthAfrica/Tygerberg_2177/2021\|EPI_ISL_3847792\|2021-08-10 | 21A (Delta) |
| hCoV-19/SouthAfrica/Tygerberg_2178/2021\|EPI_ISL_3847746\|2021-08-10 | 21A (Delta) |
| hCoV-19/SouthAfrica/Tygerberg_2179/2021\|EPI_ISL_3847886\|2021-08-10 | 21A (Delta) |
| hCoV-19/SouthAfrica/Tygerberg_2180/2021\|EPI_ISL_3847889\|2021-08-10 | 21A (Delta) |
| hCoV-19/SouthAfrica/Tygerberg_2181/2021\|EPI_ISL_3847935\|2021-08-10 | 21A (Delta) |
| hCoV-19/SouthAfrica/Tygerberg_2182/2021\|EPI_ISL_3847727\|2021-08-10 | 21A (Delta) |
| hCoV-19/SouthAfrica/Tygerberg_2183/2021\|EPI_ISL_3847795\|2021-08-10 | 21A (Delta) |
| hCoV-19/SouthAfrica/Tygerberg_2185/2021\|EPI_ISL_3847890\|2021-08-10 | 21A (Delta) |
| hCoV-19/SouthAfrica/Tygerberg_2186/2021\|EPI_ISL_3847914\|2021-08-10 | 21A (Delta) |
| hCoV-19/SouthAfrica/Tygerberg_2187/2021\|EPI_ISL_3847859\|2021-08-11 | 21A (Delta) |
| hCoV-19/SouthAfrica/Tygerberg_2188/2021\|EPI_ISL_3847777\|2021-08-11 | 21A (Delta) |
| hCoV-19/SouthAfrica/Tygerberg_2189/2021\|EPI_ISL_3847891\|2021-08-11 | 21A (Delta) |
| hCoV-19/SouthAfrica/Tygerberg_2190/2021\|EPI_ISL_3847798\|2021-08-11 | 21A (Delta) |
| hCoV-19/SouthAfrica/Tygerberg_2191/2021\|EPI_ISL_3847736\|2021-08-11 | 21A (Delta) |
| hCoV-19/SouthAfrica/Tygerberg_2194/2021\|EPI_ISL_3847923\|2021-08-11 | 21A (Delta) |
| hCoV-19/SouthAfrica/Tygerberg_2195/2021\|EPI_ISL_3847917\|2021-08-11 | 21A (Delta) |
| hCoV-19/SouthAfrica/Tygerberg_2196/2021\|EPI_ISL_3847863\|2021-08-11 | 21A (Delta) |
| hCoV-19/SouthAfrica/Tygerberg_2197/2021\|EPI_ISL_3847834\|2021-08-10 | 21A (Delta) |
| hCoV-19/SouthAfrica/Tygerberg_2198/2021\|EPI_ISL_3847762\|2021-08-11 | 21A (Delta) |
| hCoV-19/SouthAfrica/Tygerberg_2199/2021\|EPI_ISL_3847912\|2021-08-11 | 21A (Delta) |
| hCoV-19/SouthAfrica/Tygerberg_2201/2021\|EPI_ISL_3847730\|2021-08-11 | 21A (Delta) |
| hCoV-19/SouthAfrica/Tygerberg_2202/2021\|EPI_ISL_3847820\|2021-08-11 | 21A (Delta) |
| hCoV-19/SouthAfrica/Tygerberg_2204/2021\|EPI_ISL_3856198\|2021-08-12 | 21A (Delta) |
| hCoV-19/SouthAfrica/Tygerberg_2205/2021\|EPI_ISL_3856261\|2021-08-12 | 21A (Delta) |
| hCoV-19/SouthAfrica/Tygerberg_2207/2021\|EPI_ISL_3856301\|2021-08-11 | 21A (Delta) |
| hCoV-19/SouthAfrica/Tygerberg_2208/2021\|EPI_ISL_3856185\|2021-08-11 | 21A (Delta) |
| hCoV-19/SouthAfrica/Tygerberg_2209/2021\|EPI_ISL_3856263\|2021-08-11 | 21A (Delta) |
| hCoV-19/SouthAfrica/Tygerberg_2210/2021\|EPI_ISL_3856176\|2021-08-11 | 21A (Delta) |
| hCoV-19/SouthAfrica/Tygerberg_2211/2021\|EPI_ISL_3856170\|2021-08-11 | 21A (Delta) |
| hCoV-19/SouthAfrica/Tygerberg_2212/2021\|EPI_ISL_3856191\|2021-08-11 | 21A (Delta) |
| hCoV-19/SouthAfrica/Tygerberg_2213/2021\|EPI_ISL_3856182\|2021-08-12 | 21A (Delta) |
| hCoV-19/SouthAfrica/Tygerberg_2214/2021\|EPI_ISL_3856286\|2021-08-12 | 21A (Delta) |
| hCoV-19/SouthAfrica/Tygerberg_2215/2021\|EPI_ISL_3856288\|2021-08-11 | 21A (Delta) |
| hCoV-19/SouthAfrica/Tygerberg_2216/2021\|EPI_ISL_3856290\|2021-08-15 | 21A (Delta) |
| hCoV-19/SouthAfrica/Tygerberg_2217/2021\|EPI_ISL_3856278\|2021-08-15 | 21A (Delta) |
| hCoV-19/SouthAfrica/Tygerberg_2218/2021\|EPI_ISL_3856336\|2021-08-15 | 21A (Delta) |
| hCoV-19/SouthAfrica/Tygerberg_2219/2021\|EPI_ISL_3856211\|2021-08-15 | 21A (Delta) |
| hCoV-19/SouthAfrica/Tygerberg_2220/2021\|EPI_ISL_3856206\|2021-08-15 | 21A (Delta) |
| hCoV-19/SouthAfrica/Tygerberg_2221/2021\|EPI_ISL_3856187\|2021-08-15 | 21A (Delta) |
| hCoV-19/SouthAfrica/Tygerberg_2222/2021\|EPI_ISL_3856189\|2021-08-15 | 21A (Delta) |
| hCoV-19/SouthAfrica/Tygerberg_2223/2021\|EPI_ISL_3856196\|2021-08-12 | 21A (Delta) |
| hCoV-19/SouthAfrica/Tygerberg_2224/2021\|EPI_ISL_3856207\|2021-08-15 | 21A (Delta) |
| hCoV-19/SouthAfrica/Tygerberg_2225/2021\|EPI_ISL_3856338\|2021-08-15 | 21A (Delta) |
| hCoV-19/SouthAfrica/Tygerberg_2227/2021\|EPI_ISL_3856203\|2021-08-16 | 21A (Delta) |
| hCoV-19/SouthAfrica/Tygerberg_2228/2021\|EPI_ISL_3856228\|2021-08-16 | 21A (Delta) |
| hCoV-19/SouthAfrica/Tygerberg_2229/2021\|EPI_ISL_3856230\|2021-08-16 | 21A (Delta) |

| | |
|---|---|
| hCoV-19/SouthAfrica/Tygerberg_2230/2021\|EPI_ISL_3856200\|2021-08-14 | 21A (Delta) |
| hCoV-19/SouthAfrica/Tygerberg_2231/2021\|EPI_ISL_3856291\|2021-08-15 | 21A (Delta) |
| hCoV-19/SouthAfrica/Tygerberg_2232/2021\|EPI_ISL_3856217\|2021-08-15 | 21A (Delta) |
| hCoV-19/SouthAfrica/Tygerberg_2233/2021\|EPI_ISL_3856239\|2021-08-15 | 21A (Delta) |
| hCoV-19/SouthAfrica/Tygerberg_2235/2021\|EPI_ISL_3856234\|2021-08-15 | 21A (Delta) |
| hCoV-19/SouthAfrica/Tygerberg_2237/2021\|EPI_ISL_3856267\|2021-08-14 | 21A (Delta) |
| hCoV-19/SouthAfrica/Tygerberg_2238/2021\|EPI_ISL_3856269\|2021-08-14 | 21A (Delta) |
| hCoV-19/SouthAfrica/Tygerberg_2239/2021\|EPI_ISL_3856271\|2021-08-14 | 21A (Delta) |
| hCoV-19/SouthAfrica/Tygerberg_2240/2021\|EPI_ISL_3856192\|2021-08-14 | 21A (Delta) |
| hCoV-19/SouthAfrica/Tygerberg_2241/2021\|EPI_ISL_3856273\|2021-08-17 | 21A (Delta) |
| hCoV-19/SouthAfrica/Tygerberg_2244/2021\|EPI_ISL_3856321\|2021-08-16 | 21A (Delta) |
| hCoV-19/SouthAfrica/Tygerberg_2245/2021\|EPI_ISL_3856284\|2021-08-16 | 21A (Delta) |
| hCoV-19/SouthAfrica/Tygerberg_2246/2021\|EPI_ISL_3856326\|2021-08-16 | 21A (Delta) |
| hCoV-19/SouthAfrica/Tygerberg_2247/2021\|EPI_ISL_3856249\|2021-08-16 | 21A (Delta) |
| hCoV-19/SouthAfrica/Tygerberg_2248/2021\|EPI_ISL_3856202\|2021-08-16 | 21A (Delta) |
| hCoV-19/SouthAfrica/Tygerberg_2249/2021\|EPI_ISL_3856306\|2021-08-16 | 21A (Delta) |
| hCoV-19/SouthAfrica/Tygerberg_2250/2021\|EPI_ISL_3856178\|2021-08-16 | 21A (Delta) |
| hCoV-19/SouthAfrica/Tygerberg_2252/2021\|EPI_ISL_3856265\|2021-08-16 | 21A (Delta) |
| hCoV-19/SouthAfrica/Tygerberg_2253/2021\|EPI_ISL_3856226\|2021-08-16 | 21A (Delta) |
| hCoV-19/SouthAfrica/Tygerberg_2254/2021\|EPI_ISL_3856194\|2021-08-16 | 21A (Delta) |
| hCoV-19/SouthAfrica/Tygerberg_2255/2021\|EPI_ISL_3856293\|2021-08-16 | 21A (Delta) |
| hCoV-19/SouthAfrica/Tygerberg_2257/2021\|EPI_ISL_3856309\|2021-08-16 | 21A (Delta) |
| hCoV-19/SouthAfrica/Tygerberg_2260/2021\|EPI_ISL_3856282\|2021-08-16 | 21A (Delta) |
| hCoV-19/SouthAfrica/Tygerberg_2261/2021\|EPI_ISL_3856251\|2021-08-16 | 21A (Delta) |
| hCoV-19/SouthAfrica/Tygerberg_2262/2021\|EPI_ISL_3856330\|2021-08-16 | 21A (Delta) |
| hCoV-19/SouthAfrica/Tygerberg_2268/2021\|EPI_ISL_3856232\|2021-08-16 | 21A (Delta) |
| hCoV-19/SouthAfrica/Tygerberg_2270/2021\|EPI_ISL_3856295\|2021-08-19 | 21A (Delta) |
| hCoV-19/SouthAfrica/Tygerberg_2271/2021\|EPI_ISL_3856297\|2021-08-16 | 21A (Delta) |
| hCoV-19/SouthAfrica/Tygerberg_2272/2021\|EPI_ISL_3856209\|2021-08-16 | 21A (Delta) |
| hCoV-19/SouthAfrica/Tygerberg_2275/2021\|EPI_ISL_3856184\|2021-08-18 | 21A (Delta) |
| hCoV-19/SouthAfrica/Tygerberg_2276/2021\|EPI_ISL_3856253\|2021-08-18 | 21A (Delta) |
| hCoV-19/SouthAfrica/Tygerberg_2277/2021\|EPI_ISL_3856255\|2021-08-18 | 21A (Delta) |
| hCoV-19/SouthAfrica/Tygerberg_2278/2021\|EPI_ISL_3856257\|2021-08-18 | 21A (Delta) |
| hCoV-19/SouthAfrica/Tygerberg_2279/2021\|EPI_ISL_3856213\|2021-08-20 | 21A (Delta) |
| hCoV-19/SouthAfrica/Tygerberg_2280/2021\|EPI_ISL_3856174\|2021-08-20 | 21A (Delta) |
| hCoV-19/SouthAfrica/Tygerberg_2281/2021\|EPI_ISL_3856317\|2021-08-20 | 21A (Delta) |
| hCoV-19/SouthAfrica/Tygerberg_2283/2021\|EPI_ISL_3856180\|2021-08-19 | 21A (Delta) |
| hCoV-19/SouthAfrica/Tygerberg_2284/2021\|EPI_ISL_3856241\|2021-08-19 | 21A (Delta) |
| hCoV-19/SouthAfrica/Tygerberg_2285/2021\|EPI_ISL_3856313\|2021-08-19 | 21A (Delta) |
| hCoV-19/SouthAfrica/Tygerberg_2286/2021\|EPI_ISL_3856259\|2021-08-19 | 21A (Delta) |
| hCoV-19/SouthAfrica/Tygerberg_2287/2021\|EPI_ISL_3856341\|2021-08-19 | 21A (Delta) |
| hCoV-19/SouthAfrica/Tygerberg_2288/2021\|EPI_ISL_3856237\|2021-08-20 | 21A (Delta) |
| hCoV-19/SouthAfrica/Tygerberg_2289/2021\|EPI_ISL_3856340\|2021-08-20 | 21A (Delta) |
| hCoV-19/SouthAfrica/Tygerberg_2290/2021\|EPI_ISL_3856221\|2021-08-20 | 21A (Delta) |
| hCoV-19/SouthAfrica/Tygerberg_2294/2021\|EPI_ISL_3856222\|2021-08-20 | 21A (Delta) |
| hCoV-19/SouthAfrica/Tygerberg_2295/2021\|EPI_ISL_3856224\|2021-08-19 | 21A (Delta) |
| hCoV-19/South_Africa/CERI-KRISP-K020104/2021\|EPI_ISL_3267725\|2021-06-30 | 21A (Delta) |
| hCoV-19/South_Africa/CERI-KRISP-Tyg1627/2021\|EPI_ISL_3827559\|2021-07-26 | 21A (Delta) |
| hCoV-19/South_Africa/CERI-KRISP-Tyg1675/2021\|EPI_ISL_3827648\|2021-07-26 | 21A (Delta) |
| hCoV-19/South_Africa/CERI-KRISP-Tyg1689/2021\|EPI_ISL_3827649\|2021-07-27 | 21A (Delta) |

| | |
|---|---|
| hCoV-19/South_Africa/CERI-KRISP-Tyg1690/2021\|EPI_ISL_3827650\|2021-07-27 | 21A (Delta) |
| hCoV-19/South_Africa/CERI-KRISP-Tyg1691/2021\|EPI_ISL_3827651\|2021-07-27 | 21A (Delta) |
| hCoV-19/South_Africa/CERI-KRISP-Tyg1692/2021\|EPI_ISL_3827652\|2021-07-27 | 21A (Delta) |
| hCoV-19/South_Africa/CERI-KRISP-Tyg1693/2021\|EPI_ISL_3827653\|2021-07-27 | 21A (Delta) |
| hCoV-19/South_Africa/CERI-KRISP-Tyg1694/2021\|EPI_ISL_3827654\|2021-07-27 | 21A (Delta) |
| hCoV-19/South_Africa/CERI-KRISP-Tyg1695/2021\|EPI_ISL_3827655\|2021-07-27 | 21A (Delta) |
| hCoV-19/South_Africa/CERI-KRISP-Tyg1696/2021\|EPI_ISL_3827597\|2021-07-27 | 21A (Delta) |
| hCoV-19/South_Africa/CERI-KRISP-Tyg1697/2021\|EPI_ISL_3827656\|2021-07-26 | 21A (Delta) |
| hCoV-19/South_Africa/CERI-KRISP-Tyg1698/2021\|EPI_ISL_3827657\|2021-07-27 | 21A (Delta) |
| hCoV-19/South_Africa/CERI-KRISP-Tyg1699/2021\|EPI_ISL_3827590\|2021-07-27 | 21A (Delta) |
| hCoV-19/South_Africa/CERI-KRISP-Tyg1700/2021\|EPI_ISL_3827658\|2021-07-27 | 21A (Delta) |
| hCoV-19/South_Africa/CERI-KRISP-Tyg1701/2021\|EPI_ISL_3827659\|2021-07-27 | 21A (Delta) |
| hCoV-19/South_Africa/CERI-KRISP-Tyg1702/2021\|EPI_ISL_3827660\|2021-07-27 | 21A (Delta) |
| hCoV-19/South_Africa/CERI-KRISP-Tyg1703/2021\|EPI_ISL_3827589\|2021-07-27 | 21A (Delta) |
| hCoV-19/South_Africa/CERI-KRISP-Tyg1704/2021\|EPI_ISL_3827581\|2021-07-27 | 21A (Delta) |
| hCoV-19/South_Africa/CERI-KRISP-Tyg1705/2021\|EPI_ISL_3827646\|2021-07-27 | 21A (Delta) |
| hCoV-19/South_Africa/CERI-KRISP-Tyg1706/2021\|EPI_ISL_3827661\|2021-07-27 | 21A (Delta) |
| hCoV-19/South_Africa/CERI-KRISP-Tyg1707/2021\|EPI_ISL_3827662\|2021-07-27 | 21A (Delta) |
| hCoV-19/South_Africa/CERI-KRISP-Tyg1708/2021\|EPI_ISL_3827663\|2021-07-27 | 21A (Delta) |
| hCoV-19/South_Africa/CERI-KRISP-Tyg1709/2021\|EPI_ISL_3827664\|2021-07-27 | 21A (Delta) |
| hCoV-19/South_Africa/CERI-KRISP-Tyg1710/2021\|EPI_ISL_3827594\|2021-07-27 | 21A (Delta) |
| hCoV-19/South_Africa/CERI-KRISP-Tyg1711/2021\|EPI_ISL_3827665\|2021-07-27 | 21A (Delta) |
| hCoV-19/South_Africa/CERI-KRISP-Tyg1713/2021\|EPI_ISL_3827619\|2021-07-27 | 21A (Delta) |
| hCoV-19/South_Africa/CERI-KRISP-Tyg1714/2021\|EPI_ISL_3827666\|2021-07-27 | 21A (Delta) |
| hCoV-19/South_Africa/CERI-KRISP-Tyg1715/2021\|EPI_ISL_3827667\|2021-07-26 | 21A (Delta) |
| hCoV-19/South_Africa/CERI-KRISP-Tyg1716/2021\|EPI_ISL_3827668\|2021-07-27 | 21A (Delta) |
| hCoV-19/South_Africa/CERI-KRISP-Tyg1717/2021\|EPI_ISL_3827611\|2021-07-27 | 21A (Delta) |
| hCoV-19/South_Africa/CERI-KRISP-Tyg1718/2021\|EPI_ISL_3827579\|2021-07-27 | 21A (Delta) |
| hCoV-19/South_Africa/CERI-KRISP-Tyg1719/2021\|EPI_ISL_3827669\|2021-07-26 | 21A (Delta) |
| hCoV-19/South_Africa/CERI-KRISP-Tyg1720/2021\|EPI_ISL_3827586\|2021-07-27 | 21A (Delta) |
| hCoV-19/South_Africa/CERI-KRISP-Tyg1721/2021\|EPI_ISL_3827612\|2021-07-27 | 21A (Delta) |
| hCoV-19/South_Africa/CERI-KRISP-Tyg1722/2021\|EPI_ISL_3827670\|2021-07-26 | 21A (Delta) |
| hCoV-19/South_Africa/CERI-KRISP-Tyg1723/2021\|EPI_ISL_3827671\|2021-07-26 | 21A (Delta) |
| hCoV-19/South_Africa/CERI-KRISP-Tyg1724/2021\|EPI_ISL_3827672\|2021-07-27 | 21A (Delta) |
| hCoV-19/South_Africa/CERI-KRISP-Tyg1725/2021\|EPI_ISL_3827565\|2021-07-27 | 21A (Delta) |
| hCoV-19/South_Africa/CERI-KRISP-Tyg1726/2021\|EPI_ISL_3827564\|2021-07-27 | 21A (Delta) |
| hCoV-19/South_Africa/CERI-KRISP-Tyg1727/2021\|EPI_ISL_3827568\|2021-07-26 | 21A (Delta) |
| hCoV-19/South_Africa/CERI-KRISP-Tyg1728/2021\|EPI_ISL_3827673\|2021-07-27 | 21A (Delta) |
| hCoV-19/South_Africa/CERI-KRISP-Tyg1729/2021\|EPI_ISL_3827674\|2021-07-27 | 21A (Delta) |
| hCoV-19/South_Africa/CERI-KRISP-Tyg1730/2021\|EPI_ISL_3827601\|2021-07-27 | 21A (Delta) |
| hCoV-19/South_Africa/CERI-KRISP-Tyg1731/2021\|EPI_ISL_3827675\|2021-07-27 | 21A (Delta) |
| hCoV-19/South_Africa/CERI-KRISP-Tyg1732/2021\|EPI_ISL_3827647\|2021-07-27 | 21A (Delta) |
| hCoV-19/South_Africa/CERI-KRISP-Tyg1733/2021\|EPI_ISL_3827676\|2021-07-27 | 21A (Delta) |
| hCoV-19/South_Africa/CERI-KRISP-Tyg1734/2021\|EPI_ISL_3827608\|2021-07-27 | 21A (Delta) |
| hCoV-19/South_Africa/CERI-KRISP-Tyg1735/2021\|EPI_ISL_3827569\|2021-07-27 | 21A (Delta) |
| hCoV-19/South_Africa/CERI-KRISP-Tyg1736/2021\|EPI_ISL_3827606\|2021-07-27 | 21A (Delta) |
| hCoV-19/South_Africa/CERI-KRISP-Tyg1737/2021\|EPI_ISL_3827566\|2021-07-28 | 21A (Delta) |
| hCoV-19/South_Africa/CERI-KRISP-Tyg1738/2021\|EPI_ISL_3827677\|2021-07-28 | 21A (Delta) |
| hCoV-19/South_Africa/CERI-KRISP-Tyg1739/2021\|EPI_ISL_3827678\|2021-07-28 | 21A (Delta) |
| hCoV-19/South_Africa/CERI-KRISP-Tyg1740/2021\|EPI_ISL_3827679\|2021-07-28 | 21A (Delta) |

| | |
|---|---|
| hCoV-19/South_Africa/CERI-KRISP-Tyg1741/2021\|EPI_ISL_3827572\|2021-07-27 | 21A (Delta) |
| hCoV-19/South_Africa/CERI-KRISP-Tyg1742/2021\|EPI_ISL_3827680\|2021-07-27 | 21A (Delta) |
| hCoV-19/South_Africa/CERI-KRISP-Tyg1745/2021\|EPI_ISL_3827620\|2021-07-28 | 21A (Delta) |
| hCoV-19/South_Africa/CERI-KRISP-Tyg1746/2021\|EPI_ISL_3827681\|2021-07-27 | 21A (Delta) |
| hCoV-19/South_Africa/CERI-KRISP-Tyg1747/2021\|EPI_ISL_3827682\|2021-07-28 | 21A (Delta) |
| hCoV-19/South_Africa/CERI-KRISP-Tyg1748/2021\|EPI_ISL_3827621\|2021-07-28 | 21A (Delta) |
| hCoV-19/South_Africa/CERI-KRISP-Tyg1749/2021\|EPI_ISL_3827683\|2021-07-28 | 21A (Delta) |
| hCoV-19/South_Africa/CERI-KRISP-Tyg1751/2021\|EPI_ISL_3827577\|2021-07-28 | 21A (Delta) |
| hCoV-19/South_Africa/CERI-KRISP-Tyg1752/2021\|EPI_ISL_3827684\|2021-07-28 | 21A (Delta) |
| hCoV-19/South_Africa/CERI-KRISP-Tyg1753/2021\|EPI_ISL_3827685\|2021-07-28 | 21A (Delta) |
| hCoV-19/South_Africa/CERI-KRISP-Tyg1754/2021\|EPI_ISL_3827686\|2021-07-28 | 21A (Delta) |
| hCoV-19/South_Africa/CERI-KRISP-Tyg1755/2021\|EPI_ISL_3827687\|2021-07-28 | 21A (Delta) |
| hCoV-19/South_Africa/CERI-KRISP-Tyg1756/2021\|EPI_ISL_3827688\|2021-07-28 | 21A (Delta) |
| hCoV-19/South_Africa/CERI-KRISP-Tyg1757/2021\|EPI_ISL_3827689\|2021-07-28 | 21A (Delta) |
| hCoV-19/South_Africa/CERI-KRISP-Tyg1758/2021\|EPI_ISL_3827615\|2021-07-28 | 21A (Delta) |
| hCoV-19/South_Africa/CERI-KRISP-Tyg1759/2021\|EPI_ISL_3827574\|2021-07-28 | 21A (Delta) |
| hCoV-19/South_Africa/CERI-KRISP-Tyg1761/2021\|EPI_ISL_3827639\|2021-07-28 | 21A (Delta) |
| hCoV-19/South_Africa/CERI-KRISP-Tyg1762/2021\|EPI_ISL_3827690\|2021-07-28 | 21A (Delta) |
| hCoV-19/South_Africa/CERI-KRISP-Tyg1763/2021\|EPI_ISL_3827691\|2021-07-28 | 21A (Delta) |
| hCoV-19/South_Africa/CERI-KRISP-Tyg1765/2021\|EPI_ISL_3827692\|2021-07-27 | 21A (Delta) |
| hCoV-19/South_Africa/CERI-KRISP-Tyg1766/2021\|EPI_ISL_3827693\|2021-07-28 | 21A (Delta) |
| hCoV-19/South_Africa/CERI-KRISP-Tyg1767/2021\|EPI_ISL_3827694\|2021-07-28 | 21A (Delta) |
| hCoV-19/South_Africa/CERI-KRISP-Tyg1768/2021\|EPI_ISL_3827695\|2021-07-28 | 21A (Delta) |
| hCoV-19/South_Africa/CERI-KRISP-Tyg1769/2021\|EPI_ISL_3827696\|2021-07-28 | 21A (Delta) |
| hCoV-19/South_Africa/CERI-KRISP-Tyg1770/2021\|EPI_ISL_3827637\|2021-07-28 | 21A (Delta) |
| hCoV-19/South_Africa/CERI-KRISP-Tyg1771/2021\|EPI_ISL_3827697\|2021-07-28 | 21A (Delta) |
| hCoV-19/South_Africa/CERI-KRISP-Tyg1772/2021\|EPI_ISL_3827698\|2021-07-28 | 21A (Delta) |
| hCoV-19/South_Africa/CERI-KRISP-Tyg1773/2021\|EPI_ISL_3827699\|2021-07-28 | 21A (Delta) |
| hCoV-19/South_Africa/CERI-KRISP-Tyg1774/2021\|EPI_ISL_3827700\|2021-07-28 | 21A (Delta) |
| hCoV-19/South_Africa/CERI-KRISP-Tyg1775/2021\|EPI_ISL_3827701\|2021-07-28 | 21A (Delta) |
| hCoV-19/South_Africa/CERI-KRISP-Tyg1776/2021\|EPI_ISL_3827638\|2021-07-28 | 21A (Delta) |
| hCoV-19/South_Africa/CERI-KRISP-Tyg1777/2021\|EPI_ISL_3827702\|2021-07-27 | 21A (Delta) |
| hCoV-19/South_Africa/CERI-KRISP-Tyg1778/2021\|EPI_ISL_3827703\|2021-07-28 | 21A (Delta) |
| hCoV-19/South_Africa/CERI-KRISP-Tyg1779/2021\|EPI_ISL_3827704\|2021-07-28 | 21A (Delta) |
| hCoV-19/South_Africa/CERI-KRISP-Tyg1780/2021\|EPI_ISL_3827705\|2021-07-28 | 21A (Delta) |
| hCoV-19/South_Africa/CERI-KRISP-Tyg1781/2021\|EPI_ISL_3827706\|2021-07-28 | 21A (Delta) |
| hCoV-19/South_Africa/CERI-KRISP-Tyg1782/2021\|EPI_ISL_3827707\|2021-07-28 | 21A (Delta) |
| hCoV-19/South_Africa/CERI-KRISP-Tyg1783/2021\|EPI_ISL_3827708\|2021-07-28 | 21A (Delta) |
| hCoV-19/South_Africa/CERI-KRISP-Tyg1784/2021\|EPI_ISL_3827709\|2021-07-28 | 21A (Delta) |
| hCoV-19/South_Africa/CERI-KRISP-Tyg1785/2021\|EPI_ISL_3827602\|2021-07-28 | 21A (Delta) |
| hCoV-19/South_Africa/CERI-KRISP-Tyg1786/2021\|EPI_ISL_3827710\|2021-07-28 | 21A (Delta) |
| hCoV-19/South_Africa/CERI-KRISP-Tyg1787/2021\|EPI_ISL_3827711\|2021-07-27 | 21A (Delta) |
| hCoV-19/South_Africa/CERI-KRISP-Tyg1788/2021\|EPI_ISL_3827712\|2021-07-27 | 21A (Delta) |
| hCoV-19/South_Africa/CERI-KRISP-Tyg1789/2021\|EPI_ISL_3827713\|2021-07-28 | 21A (Delta) |
| hCoV-19/South_Africa/CERI-KRISP-Tyg1790/2021\|EPI_ISL_3827714\|2021-07-28 | 21A (Delta) |
| hCoV-19/South_Africa/CERI-KRISP-Tyg1791/2021\|EPI_ISL_3827600\|2021-07-28 | 21A (Delta) |
| hCoV-19/South_Africa/CERI-KRISP-Tyg1792/2021\|EPI_ISL_3827603\|2021-07-28 | 21A (Delta) |
| hCoV-19/South_Africa/CERI-KRISP-Tyg1794/2021\|EPI_ISL_3827642\|2021-07-28 | 21A (Delta) |
| hCoV-19/South_Africa/CERI-KRISP-Tyg1795/2021\|EPI_ISL_3827715\|2021-07-28 | 21A (Delta) |
| hCoV-19/South_Africa/CERI-KRISP-Tyg1796/2021\|EPI_ISL_3827716\|2021-07-28 | 21A (Delta) |

| | |
|---|---|
| hCoV-19/South_Africa/CERI-KRISP-Tyg1797/2021\|EPI_ISL_3827643\|2021-07-28 | 21A (Delta) |
| hCoV-19/South_Africa/CERI-KRISP-Tyg1798/2021\|EPI_ISL_3827717\|2021-07-28 | 21A (Delta) |
| hCoV-19/South_Africa/CERI-KRISP-Tyg1799/2021\|EPI_ISL_3827718\|2021-07-28 | 21A (Delta) |
| hCoV-19/South_Africa/CERI-KRISP-Tyg1800/2021\|EPI_ISL_3827588\|2021-07-28 | 21A (Delta) |
| hCoV-19/South_Africa/CERI-KRISP-Tyg1801/2021\|EPI_ISL_3827719\|2021-07-28 | 21A (Delta) |
| hCoV-19/South_Africa/CERI-KRISP-Tyg1802/2021\|EPI_ISL_3827720\|2021-07-28 | 21A (Delta) |
| hCoV-19/South_Africa/CERI-KRISP-Tyg1803/2021\|EPI_ISL_3827634\|2021-07-28 | 21A (Delta) |
| hCoV-19/South_Africa/CERI-KRISP-Tyg1804/2021\|EPI_ISL_3827721\|2021-07-28 | 21A (Delta) |
| hCoV-19/South_Africa/CERI-KRISP-Tyg1805/2021\|EPI_ISL_3827722\|2021-07-28 | 21A (Delta) |
| hCoV-19/South_Africa/CERI-KRISP-Tyg1806/2021\|EPI_ISL_3827723\|2021-07-28 | 21A (Delta) |
| hCoV-19/South_Africa/CERI-KRISP-Tyg1807/2021\|EPI_ISL_3827575\|2021-07-28 | 21A (Delta) |
| hCoV-19/South_Africa/CERI-KRISP-Tyg1808/2021\|EPI_ISL_3827567\|2021-07-28 | 21A (Delta) |
| hCoV-19/South_Africa/CERI-KRISP-Tyg1809/2021\|EPI_ISL_3827583\|2021-07-28 | 21A (Delta) |
| hCoV-19/South_Africa/CERI-KRISP-Tyg1810/2021\|EPI_ISL_3827724\|2021-07-28 | 21A (Delta) |
| hCoV-19/South_Africa/CERI-KRISP-Tyg1811/2021\|EPI_ISL_3827725\|2021-07-28 | 21A (Delta) |
| hCoV-19/South_Africa/CERI-KRISP-Tyg1812/2021\|EPI_ISL_3827726\|2021-07-28 | 21A (Delta) |
| hCoV-19/South_Africa/CERI-KRISP-Tyg1813/2021\|EPI_ISL_3827727\|2021-07-28 | 21A (Delta) |
| hCoV-19/South_Africa/CERI-KRISP-Tyg1814/2021\|EPI_ISL_3827728\|2021-07-28 | 21A (Delta) |
| hCoV-19/South_Africa/CERI-KRISP-Tyg1815/2021\|EPI_ISL_3827729\|2021-07-28 | 21A (Delta) |
| hCoV-19/South_Africa/CERI-KRISP-Tyg1816/2021\|EPI_ISL_3827613\|2021-07-28 | 21A (Delta) |
| hCoV-19/South_Africa/CERI-KRISP-Tyg1817/2021\|EPI_ISL_3827599\|2021-07-28 | 21A (Delta) |
| hCoV-19/South_Africa/CERI-KRISP-Tyg1818/2021\|EPI_ISL_3827730\|2021-07-28 | 21A (Delta) |
| hCoV-19/South_Africa/CERI-KRISP-Tyg1819/2021\|EPI_ISL_3827731\|2021-07-28 | 21A (Delta) |
| hCoV-19/South_Africa/CERI-KRISP-Tyg1820/2021\|EPI_ISL_3827732\|2021-07-28 | 21A (Delta) |
| hCoV-19/South_Africa/CERI-KRISP-Tyg1821/2021\|EPI_ISL_3827733\|2021-07-28 | 21A (Delta) |
| hCoV-19/South_Africa/CERI-KRISP-Tyg1822/2021\|EPI_ISL_3827636\|2021-07-28 | 21A (Delta) |
| hCoV-19/South_Africa/CERI-KRISP-Tyg1823/2021\|EPI_ISL_3827616\|2021-07-28 | 21A (Delta) |
| hCoV-19/South_Africa/CERI-KRISP-Tyg1824/2021\|EPI_ISL_3827593\|2021-07-28 | 21A (Delta) |
| hCoV-19/South_Africa/CERI-KRISP-Tyg1825/2021\|EPI_ISL_3827734\|2021-07-28 | 21A (Delta) |
| hCoV-19/South_Africa/CERI-KRISP-Tyg1826/2021\|EPI_ISL_3827735\|2021-07-28 | 21A (Delta) |
| hCoV-19/South_Africa/CERI-KRISP-Tyg1827/2021\|EPI_ISL_3827736\|2021-07-28 | 21A (Delta) |
| hCoV-19/South_Africa/CERI-KRISP-Tyg1828/2021\|EPI_ISL_3827737\|2021-07-28 | 21A (Delta) |
| hCoV-19/South_Africa/CERI-KRISP-Tyg1829/2021\|EPI_ISL_3827738\|2021-07-28 | 21A (Delta) |
| hCoV-19/South_Africa/CERI-KRISP-Tyg1831/2021\|EPI_ISL_3827635\|2021-07-28 | 21A (Delta) |
| hCoV-19/South_Africa/CERI-KRISP-Tyg1833/2021\|EPI_ISL_3827739\|2021-07-28 | 21A (Delta) |
| hCoV-19/South_Africa/CERI-KRISP-Tyg1834/2021\|EPI_ISL_3827740\|2021-07-28 | 21A (Delta) |
| hCoV-19/South_Africa/CERI-KRISP-Tyg1835/2021\|EPI_ISL_3827741\|2021-07-28 | 21A (Delta) |
| hCoV-19/South_Africa/CERI-KRISP-Tyg1836/2021\|EPI_ISL_3827742\|2021-07-28 | 21A (Delta) |
| hCoV-19/South_Africa/CERI-KRISP-Tyg1837/2021\|EPI_ISL_3827743\|2021-07-28 | 21A (Delta) |
| hCoV-19/South_Africa/CERI-KRISP-Tyg1838/2021\|EPI_ISL_3827744\|2021-07-29 | 21A (Delta) |
| hCoV-19/South_Africa/CERI-KRISP-Tyg1839/2021\|EPI_ISL_3827582\|2021-07-29 | 21A (Delta) |
| hCoV-19/South_Africa/CERI-KRISP-Tyg1840/2021\|EPI_ISL_3827591\|2021-07-29 | 21A (Delta) |
| hCoV-19/South_Africa/CERI-KRISP-Tyg1841/2021\|EPI_ISL_3827745\|2021-07-29 | 21A (Delta) |
| hCoV-19/South_Africa/CERI-KRISP-Tyg1842/2021\|EPI_ISL_3827598\|2021-07-29 | 21A (Delta) |
| hCoV-19/South_Africa/CERI-KRISP-Tyg1843/2021\|EPI_ISL_3827746\|2021-07-29 | 21A (Delta) |
| hCoV-19/South_Africa/CERI-KRISP-Tyg1844/2021\|EPI_ISL_3827747\|2021-07-29 | 21A (Delta) |
| hCoV-19/South_Africa/CERI-KRISP-Tyg1845/2021\|EPI_ISL_3827644\|2021-07-29 | 21A (Delta) |
| hCoV-19/South_Africa/CERI-KRISP-Tyg1846/2021\|EPI_ISL_3827617\|2021-07-29 | 21A (Delta) |
| hCoV-19/South_Africa/CERI-KRISP-Tyg1847/2021\|EPI_ISL_3827748\|2021-07-29 | 21A (Delta) |
| hCoV-19/South_Africa/CERI-KRISP-Tyg1848/2021\|EPI_ISL_3827749\|2021-07-29 | 21A (Delta) |

| | |
|---|---|
| hCoV-19/South_Africa/CERI-KRISP-Tyg1849/2021\|EPI_ISL_3827750\|2021-07-29 | 21A (Delta) |
| hCoV-19/South_Africa/CERI-KRISP-Tyg1850/2021\|EPI_ISL_3827751\|2021-07-29 | 21A (Delta) |
| hCoV-19/South_Africa/CERI-KRISP-Tyg1851/2021\|EPI_ISL_3827752\|2021-07-29 | 21A (Delta) |
| hCoV-19/South_Africa/CERI-KRISP-Tyg1852/2021\|EPI_ISL_3827753\|2021-07-29 | 21A (Delta) |
| hCoV-19/South_Africa/CERI-KRISP-Tyg1853/2021\|EPI_ISL_3827570\|2021-07-29 | 21A (Delta) |
| hCoV-19/South_Africa/CERI-KRISP-Tyg1855/2021\|EPI_ISL_3827618\|2021-07-29 | 21A (Delta) |
| hCoV-19/South_Africa/CERI-KRISP-Tyg1856/2021\|EPI_ISL_3827571\|2021-07-29 | 21A (Delta) |
| hCoV-19/South_Africa/CERI-KRISP-Tyg1857/2021\|EPI_ISL_3827576\|2021-07-29 | 21A (Delta) |
| hCoV-19/South_Africa/CERI-KRISP-Tyg1858/2021\|EPI_ISL_3827754\|2021-07-29 | 21A (Delta) |
| hCoV-19/South_Africa/CERI-KRISP-Tyg1859/2021\|EPI_ISL_3827755\|2021-07-29 | 21A (Delta) |
| hCoV-19/South_Africa/CERI-KRISP-Tyg1860/2021\|EPI_ISL_3827756\|2021-07-29 | 21A (Delta) |
| hCoV-19/South_Africa/CERI-KRISP-Tyg1861/2021\|EPI_ISL_3827563\|2021-07-29 | 21A (Delta) |
| hCoV-19/South_Africa/CERI-KRISP-Tyg1864/2021\|EPI_ISL_3827562\|2021-07-29 | 21A (Delta) |
| hCoV-19/South_Africa/CERI-KRISP-Tyg1866/2021\|EPI_ISL_3827757\|2021-07-29 | 21A (Delta) |
| hCoV-19/South_Africa/CERI-KRISP-Tyg1867/2021\|EPI_ISL_3827758\|2021-07-29 | 21A (Delta) |
| hCoV-19/South_Africa/CERI-KRISP-Tyg1868/2021\|EPI_ISL_3827759\|2021-07-29 | 21A (Delta) |
| hCoV-19/South_Africa/CERI-KRISP-Tyg1869/2021\|EPI_ISL_3827578\|2021-07-29 | 21A (Delta) |
| hCoV-19/South_Africa/CERI-KRISP-Tyg1870/2021\|EPI_ISL_3827573\|2021-07-29 | 21A (Delta) |
| hCoV-19/South_Africa/CERI-KRISP-Tyg1871/2021\|EPI_ISL_3827604\|2021-07-29 | 21A (Delta) |
| hCoV-19/South_Africa/CERI-KRISP-Tyg1873/2021\|EPI_ISL_3827645\|2021-07-29 | 21A (Delta) |
| hCoV-19/South_Africa/CERI-KRISP-Tyg1874/2021\|EPI_ISL_3827760\|2021-07-29 | 21A (Delta) |
| hCoV-19/South_Africa/CERI-KRISP-Tyg1875/2021\|EPI_ISL_3827761\|2021-07-29 | 21A (Delta) |
| hCoV-19/South_Africa/CERI-KRISP-Tyg1876/2021\|EPI_ISL_3827762\|2021-07-29 | 21A (Delta) |
| hCoV-19/South_Africa/CERI-KRISP-Tyg1877/2021\|EPI_ISL_3827763\|2021-07-29 | 21A (Delta) |
| hCoV-19/South_Africa/CERI-KRISP-Tyg1878/2021\|EPI_ISL_3827560\|2021-07-29 | 21A (Delta) |
| hCoV-19/South_Africa/CERI-KRISP-Tyg1879/2021\|EPI_ISL_3827764\|2021-07-29 | 21A (Delta) |
| hCoV-19/South_Africa/CERI-KRISP-Tyg1880/2021\|EPI_ISL_3827765\|2021-07-29 | 21A (Delta) |
| hCoV-19/South_Africa/CERI-KRISP-Tyg1882/2021\|EPI_ISL_3827766\|2021-07-29 | 21A (Delta) |
| hCoV-19/South_Africa/CERI-KRISP-Tyg1883/2021\|EPI_ISL_3827768\|2021-07-29 | 21A (Delta) |
| hCoV-19/South_Africa/CERI-KRISP-Tyg1884/2021\|EPI_ISL_3827769\|2021-07-29 | 21A (Delta) |
| hCoV-19/South_Africa/CERI-KRISP-Tyg1885/2021\|EPI_ISL_3827770\|2021-07-29 | 21A (Delta) |
| hCoV-19/South_Africa/CERI-KRISP-Tyg1886/2021\|EPI_ISL_3827771\|2021-07-29 | 21A (Delta) |
| hCoV-19/South_Africa/CERI-KRISP-Tyg1887/2021\|EPI_ISL_3827596\|2021-07-29 | 21A (Delta) |
| hCoV-19/South_Africa/CERI-KRISP-Tyg1888/2021\|EPI_ISL_3827772\|2021-07-29 | 21A (Delta) |
| hCoV-19/South_Africa/CERI-KRISP-Tyg1889/2021\|EPI_ISL_3827595\|2021-07-29 | 21A (Delta) |
| hCoV-19/South_Africa/CERI-KRISP-Tyg1890/2021\|EPI_ISL_3827773\|2021-07-29 | 21A (Delta) |
| hCoV-19/South_Africa/CERI-KRISP-Tyg1891/2021\|EPI_ISL_3827774\|2021-07-29 | 21A (Delta) |
| hCoV-19/South_Africa/CERI-KRISP-Tyg1892/2021\|EPI_ISL_3827775\|2021-07-29 | 21A (Delta) |
| hCoV-19/South_Africa/CERI-KRISP-Tyg1893/2021\|EPI_ISL_3827776\|2021-07-29 | 21A (Delta) |
| hCoV-19/South_Africa/CERI-KRISP-Tyg1894/2021\|EPI_ISL_3827610\|2021-07-29 | 21A (Delta) |
| hCoV-19/South_Africa/CERI-KRISP-Tyg1895/2021\|EPI_ISL_3827777\|2021-07-29 | 21A (Delta) |
| hCoV-19/South_Africa/CERI-KRISP-Tyg1896/2021\|EPI_ISL_3827607\|2021-07-29 | 21A (Delta) |
| hCoV-19/South_Africa/CERI-KRISP-Tyg1897/2021\|EPI_ISL_3827778\|2021-07-29 | 21A (Delta) |
| hCoV-19/South_Africa/CERI-KRISP-Tyg1898/2021\|EPI_ISL_3827779\|2021-07-29 | 21A (Delta) |
| hCoV-19/South_Africa/CERI-KRISP-Tyg1899/2021\|EPI_ISL_3827780\|2021-07-29 | 21A (Delta) |
| hCoV-19/South_Africa/CERI-KRISP-Tyg1900/2021\|EPI_ISL_3827781\|2021-07-30 | 21A (Delta) |
| hCoV-19/South_Africa/CERI-KRISP-Tyg1901/2021\|EPI_ISL_3827782\|2021-07-30 | 21A (Delta) |
| hCoV-19/South_Africa/CERI-KRISP-Tyg1903/2021\|EPI_ISL_3827587\|2021-07-30 | 21A (Delta) |
| hCoV-19/South_Africa/CERI-KRISP-Tyg1904/2021\|EPI_ISL_3827783\|2021-07-30 | 21A (Delta) |
| hCoV-19/South_Africa/CERI-KRISP-Tyg1905/2021\|EPI_ISL_3827561\|2021-07-30 | 21A (Delta) |

| | |
|---|---|
| hCoV-19/South_Africa/CERI-KRISP-Tyg1906/2021\|EPI_ISL_3827784\|2021-07-30 | 21A (Delta) |
| hCoV-19/South_Africa/CERI-KRISP-Tyg1907/2021\|EPI_ISL_3827785\|2021-07-30 | 21A (Delta) |
| hCoV-19/South_Africa/CERI-KRISP-Tyg1908/2021\|EPI_ISL_3827786\|2021-07-30 | 21A (Delta) |
| hCoV-19/South_Africa/CERI-KRISP-Tyg1909/2021\|EPI_ISL_3827584\|2021-07-30 | 21A (Delta) |
| hCoV-19/South_Africa/CERI-KRISP-Tyg1910/2021\|EPI_ISL_3827787\|2021-07-30 | 21A (Delta) |
| hCoV-19/South_Africa/CERI-KRISP-Tyg1912/2021\|EPI_ISL_3827788\|2021-07-30 | 21A (Delta) |
| hCoV-19/South_Africa/CERI-KRISP-Tyg1913/2021\|EPI_ISL_3827580\|2021-07-30 | 21A (Delta) |
| hCoV-19/South_Africa/CERI-KRISP-Tyg1914/2021\|EPI_ISL_3827789\|2021-07-30 | 21A (Delta) |
| hCoV-19/South_Africa/CERI-KRISP-Tyg1915/2021\|EPI_ISL_3827790\|2021-07-30 | 21A (Delta) |
| hCoV-19/South_Africa/CERI-KRISP-Tyg1916/2021\|EPI_ISL_3827791\|2021-07-30 | 21A (Delta) |
| hCoV-19/South_Africa/CERI-KRISP-Tyg1917/2021\|EPI_ISL_3827792\|2021-07-30 | 21A (Delta) |
| hCoV-19/South_Africa/CERI-KRISP-Tyg1918/2021\|EPI_ISL_3827793\|2021-07-30 | 21A (Delta) |
| hCoV-19/South_Africa/CERI-KRISP-Tyg1919/2021\|EPI_ISL_3827609\|2021-07-30 | 21A (Delta) |
| hCoV-19/South_Africa/CERI-KRISP-Tyg1920/2021\|EPI_ISL_3827794\|2021-07-30 | 21A (Delta) |
| hCoV-19/South_Africa/CERI-KRISP-Tyg1921/2021\|EPI_ISL_3827795\|2021-07-30 | 21A (Delta) |
| hCoV-19/South_Africa/CERI-KRISP-Tyg1922/2021\|EPI_ISL_3827796\|2021-07-30 | 21A (Delta) |
| hCoV-19/South_Africa/CERI-KRISP-Tyg1923/2021\|EPI_ISL_3827797\|2021-07-30 | 21A (Delta) |
| hCoV-19/South_Africa/CERI-KRISP-Tyg1924/2021\|EPI_ISL_3827798\|2021-07-30 | 21A (Delta) |
| hCoV-19/South_Africa/CERI-KRISP-Tyg1925/2021\|EPI_ISL_3827799\|2021-07-30 | 21A (Delta) |
| hCoV-19/South_Africa/CERI-KRISP-Tyg1926/2021\|EPI_ISL_3827800\|2021-07-30 | 21A (Delta) |
| hCoV-19/South_Africa/CERI-KRISP-Tyg1928/2021\|EPI_ISL_3827592\|2021-07-29 | 21A (Delta) |
| hCoV-19/South_Africa/CERI-KRISP-Tyg1930/2021\|EPI_ISL_3827801\|2021-07-30 | 21A (Delta) |
| hCoV-19/South_Africa/CERI-KRISP-Tyg1931/2021\|EPI_ISL_3827802\|2021-07-30 | 21A (Delta) |
| hCoV-19/South_Africa/CERI-KRISP-Tyg1932/2021\|EPI_ISL_3827585\|2021-07-30 | 21A (Delta) |
| hCoV-19/South_Africa/CERI-KRISP-Tyg1933/2021\|EPI_ISL_3827605\|2021-07-30 | 21A (Delta) |
| hCoV-19/South_Africa/CERI-KRISP-Tyg1934/2021\|EPI_ISL_3827803\|2021-07-30 | 21A (Delta) |
| hCoV-19/South_Africa/CERI-KRISP-Tyg1935/2021\|EPI_ISL_3827804\|2021-07-30 | 21A (Delta) |
| hCoV-19/South_Africa/CERI-KRISP-Tyg1937/2021\|EPI_ISL_3827805\|2021-07-30 | 21A (Delta) |
| hCoV-19/South_Africa/CERI-KRISP-Tyg1938/2021\|EPI_ISL_3827806\|2021-07-30 | 21A (Delta) |
| hCoV-19/South_Africa/CERI-KRISP-Tyg1939/2021\|EPI_ISL_3827807\|2021-07-30 | 21A (Delta) |
| hCoV-19/South_Africa/CERI-KRISP-Tyg1940/2021\|EPI_ISL_3827808\|2021-07-30 | 21A (Delta) |
| hCoV-19/South_Africa/CERI-KRISP-Tyg1941/2021\|EPI_ISL_3827622\|2021-07-30 | 21A (Delta) |
| hCoV-19/South_Africa/CERI-KRISP-Tyg1944/2021\|EPI_ISL_3827809\|2021-07-29 | 21A (Delta) |
| hCoV-19/South_Africa/CERI-KRISP-Tyg1945/2021\|EPI_ISL_3827810\|2021-07-30 | 21A (Delta) |
| hCoV-19/South_Africa/CERI-KRISP-Tyg1946/2021\|EPI_ISL_3827811\|2021-07-30 | 21A (Delta) |
| hCoV-19/South_Africa/CERI-KRISP-Tyg1948/2021\|EPI_ISL_3827812\|2021-07-30 | 21A (Delta) |
| hCoV-19/South_Africa/CERI-KRISP-Tyg1949/2021\|EPI_ISL_3827614\|2021-07-29 | 21A (Delta) |
| hCoV-19/South_Africa/CERI-KRISP-Tyg1950/2021\|EPI_ISL_3827623\|2021-07-30 | 21A (Delta) |
| hCoV-19/South_Africa/CERI-KRISP-Tyg1951/2021\|EPI_ISL_3827813\|2021-07-31 | 21A (Delta) |
| hCoV-19/South_Africa/CERI-KRISP-Tyg1952/2021\|EPI_ISL_3827814\|2021-07-30 | 21A (Delta) |
| hCoV-19/South_Africa/CERI-KRISP-Tyg1953/2021\|EPI_ISL_3827641\|2021-07-30 | 21A (Delta) |
| hCoV-19/South_Africa/CERI-KRISP-Tyg1954/2021\|EPI_ISL_3827815\|2021-07-30 | 21A (Delta) |
| hCoV-19/South_Africa/CERI-KRISP-Tyg1956/2021\|EPI_ISL_3827816\|2021-07-30 | 21A (Delta) |
| hCoV-19/South_Africa/CERI-KRISP-Tyg1957/2021\|EPI_ISL_3827817\|2021-07-30 | 21A (Delta) |
| hCoV-19/South_Africa/CERI-KRISP-Tyg1958/2021\|EPI_ISL_3827818\|2021-07-30 | 21A (Delta) |
| hCoV-19/South_Africa/CERI-KRISP-Tyg1959/2021\|EPI_ISL_3827819\|2021-07-30 | 21A (Delta) |
| hCoV-19/South_Africa/CERI-KRISP-Tyg1960/2021\|EPI_ISL_3827820\|2021-07-30 | 21A (Delta) |
| hCoV-19/South_Africa/KRISP-K010124/2021\|EPI_ISL_1366834\|2021-01-21 | 20H (Beta, V2) |
| hCoV-19/South_Africa/KRISP-K018720/2021\|EPI_ISL_2710322\|2021-06-13 | 20H (Beta, V2) |
| hCoV-19/South_Africa/NHLS-UCT-AM-Z017/2021\|EPI_ISL_2802186\|2021-06-02 | 20I (Alpha, V1) |

| | |
|---|---|
| hCoV-19/South_Africa/NHLS-UCT-AM-Z018/2021\|EPI_ISL_2802112\|2021-05-12 | 21A (Delta) |
| hCoV-19/South_Africa/NHLS-UCT-AM-Z019/2021\|EPI_ISL_2802113\|2021-05 | 20H (Beta, V2) |
| hCoV-19/South_Africa/NHLS-UCT-AM-Z021/2021\|EPI_ISL_2802115\|2021-05-17 | 20H (Beta, V2) |
| hCoV-19/South_Africa/NHLS-UCT-AM-Z022/2021\|EPI_ISL_2802116\|2021-05-14 | 20H (Beta, V2) |
| hCoV-19/South_Africa/NHLS-UCT-AM-Z029/2021\|EPI_ISL_3207515\|2021-04-26 | 20I (Alpha, V1) |
| hCoV-19/South_Africa/NHLS-UCT-AM-Z031/2021\|EPI_ISL_3207516\|2021-04-27 | 20I (Alpha, V1) |
| hCoV-19/South_Africa/NHLS-UCT-AM-Z032/2021\|EPI_ISL_3207517\|2021-06-19 | 21A (Delta) |
| hCoV-19/South_Africa/NHLS-UCT-AM-Z033/2021\|EPI_ISL_3207518\|2021-06-20 | 21A (Delta) |
| hCoV-19/South_Africa/NHLS-UCT-AM-Z034/2021\|EPI_ISL_3207519\|2021-06-20 | 21A (Delta) |
| hCoV-19/South_Africa/NHLS-UCT-AM-Z036/2021\|EPI_ISL_3207520\|2021-04-16 | 21A (Delta) |
| hCoV-19/South_Africa/NHLS-UCT-AM-Z037/2021\|EPI_ISL_3207521\|2021-06-20 | 20I (Alpha, V1) |
| hCoV-19/South_Africa/NHLS-UCT-AM-Z038/2021\|EPI_ISL_3207522\|2021-06-20 | 21A (Delta) |
| hCoV-19/South_Africa/NHLS-UCT-AM-Z039/2021\|EPI_ISL_3207523\|2021-06-20 | 21A (Delta) |
| hCoV-19/South_Africa/NHLS-UCT-AM-Z045/2021\|EPI_ISL_3690621\|2021-06-30 | 21A (Delta) |
| hCoV-19/South_Africa/NHLS-UCT-AM-Z046/2021\|EPI_ISL_3690628\|2021-06-27 | 21A (Delta) |
| hCoV-19/South_Africa/NHLS-UCT-AM-Z047/2021\|EPI_ISL_3690622\|2021-06-30 | 21A (Delta) |
| hCoV-19/South_Africa/NHLS-UCT-AM-Z048/2021\|EPI_ISL_3690629\|2021-06-27 | 21A (Delta) |
| hCoV-19/South_Africa/NHLS-UCT-AM-Z049/2021\|EPI_ISL_3690623\|2021-06-30 | 21A (Delta) |
| hCoV-19/South_Africa/NHLS-UCT-AM-Z050/2021\|EPI_ISL_3690551\|2021-06-27 | 20H (Beta, V2) |
| hCoV-19/South_Africa/NHLS-UCT-AM-Z051/2021\|EPI_ISL_3690550\|2021-07-03 | 21A (Delta) |
| hCoV-19/South_Africa/NHLS-UCT-AM-Z052/2021\|EPI_ISL_3690624\|2021-06-29 | 21A (Delta) |
| hCoV-19/South_Africa/NHLS-UCT-AM-Z053/2021\|EPI_ISL_3690630\|2021-06-27 | 21A (Delta) |
| hCoV-19/South_Africa/NHLS-UCT-AM-Z054/2021\|EPI_ISL_3690625\|2021-06-29 | 21A (Delta) |
| hCoV-19/South_Africa/NHLS-UCT-AM-Z055/2021\|EPI_ISL_3690631\|2021-06-26 | 21A (Delta) |
| hCoV-19/South_Africa/NHLS-UCT-AM-Z056/2021\|EPI_ISL_3690555\|2021-06-30 | 21A (Delta) |
| hCoV-19/South_Africa/NHLS-UCT-AM-Z058/2021\|EPI_ISL_3690547\|2021-07-10 | 21A (Delta) |
| hCoV-19/South_Africa/NHLS-UCT-AM-Z060/2021\|EPI_ISL_3690639\|2021-06-30 | 21A (Delta) |
| hCoV-19/South_Africa/NHLS-UCT-AM-Z061/2021\|EPI_ISL_3690626\|2021-06-29 | 21A (Delta) |
| hCoV-19/South_Africa/NHLS-UCT-AM-Z064/2021\|EPI_ISL_3690627\|2021-06-28 | 21A (Delta) |
| hCoV-19/South_Africa/NHLS-UCT-GP-6147/2021\|EPI_ISL_1817717\|2021-03-08 | 20H (Beta, V2) |
| hCoV-19/South_Africa/NHLS-UCT-GP-6148/2021\|EPI_ISL_1817718\|2021-03-11 | 20H (Beta, V2) |
| hCoV-19/South_Africa/NHLS-UCT-GP-6149/2021\|EPI_ISL_1817719\|2021-03-02 | 20H (Beta, V2) |
| hCoV-19/South_Africa/NHLS-UCT-GP-6150/2021\|EPI_ISL_1817720\|2021-02-17 | 20H (Beta, V2) |
| hCoV-19/South_Africa/NHLS-UCT-GP-6151/2021\|EPI_ISL_1817672\|2021-03-08 | 20H (Beta, V2) |
| hCoV-19/South_Africa/NHLS-UCT-GP-6152/2021\|EPI_ISL_1817721\|2021-03-01 | 20H (Beta, V2) |
| hCoV-19/South_Africa/NHLS-UCT-GP-6153/2021\|EPI_ISL_1817722\|2021-03-04 | 20H (Beta, V2) |
| hCoV-19/South_Africa/NHLS-UCT-GP-6154/2021\|EPI_ISL_1817678\|2021-03-08 | 20H (Beta, V2) |
| hCoV-19/South_Africa/NHLS-UCT-GP-6155/2021\|EPI_ISL_1817723\|2021-02-19 | 20H (Beta, V2) |
| hCoV-19/South_Africa/NHLS-UCT-GP-6156/2021\|EPI_ISL_1817724\|2021-03-08 | 20H (Beta, V2) |
| hCoV-19/South_Africa/NHLS-UCT-GP-6157/2021\|EPI_ISL_1817725\|2021-02-17 | 20H (Beta, V2) |
| hCoV-19/South_Africa/NHLS-UCT-GP-6159/2021\|EPI_ISL_1817660\|2021-03-08 | 20H (Beta, V2) |
| hCoV-19/South_Africa/NHLS-UCT-GP-6160/2021\|EPI_ISL_1817661\|2021-03-08 | 20H (Beta, V2) |
| hCoV-19/South_Africa/NHLS-UCT-GP-6161/2021\|EPI_ISL_1817726\|2021-03-01 | 20H (Beta, V2) |
| hCoV-19/South_Africa/NHLS-UCT-GP-6162/2021\|EPI_ISL_1817727\|2021-03-10 | 20H (Beta, V2) |
| hCoV-19/South_Africa/NHLS-UCT-GP-6163/2021\|EPI_ISL_1817676\|2021-02-17 | 20H (Beta, V2) |
| hCoV-19/South_Africa/NHLS-UCT-GP-6164/2021\|EPI_ISL_1817728\|2021-02-25 | 20H (Beta, V2) |
| hCoV-19/South_Africa/NHLS-UCT-GP-6165/2021\|EPI_ISL_1817729\|2021-03-02 | 20H (Beta, V2) |
| hCoV-19/South_Africa/NHLS-UCT-GP-6166/2021\|EPI_ISL_1817730\|2021-03-05 | 20H (Beta, V2) |
| hCoV-19/South_Africa/NHLS-UCT-GP-6167/2021\|EPI_ISL_1817708\|2021-03-08 | 20H (Beta, V2) |
| hCoV-19/South_Africa/NHLS-UCT-GP-6168/2021\|EPI_ISL_1817705\|2021-03-08 | 20H (Beta, V2) |

| | |
|---|---|
| hCoV-19/South_Africa/NHLS-UCT-GP-6169/2021\|EPI_ISL_1817731\|2021-03-01 | 20H (Beta, V2) |
| hCoV-19/South_Africa/NHLS-UCT-GP-6170/2021\|EPI_ISL_1817695\|2021-03-08 | 20H (Beta, V2) |
| hCoV-19/South_Africa/NHLS-UCT-GP-6171/2021\|EPI_ISL_1817685\|2021-03-08 | 20H (Beta, V2) |
| hCoV-19/South_Africa/NHLS-UCT-GP-6173/2021\|EPI_ISL_1817732\|2021-02-16 | 20H (Beta, V2) |
| hCoV-19/South_Africa/NHLS-UCT-GP-6174/2021\|EPI_ISL_1817666\|2021-03-08 | 20H (Beta, V2) |
| hCoV-19/South_Africa/NHLS-UCT-GP-6175/2021\|EPI_ISL_1817677\|2021-03-09 | 20H (Beta, V2) |
| hCoV-19/South_Africa/NHLS-UCT-GP-6178/2021\|EPI_ISL_2375910\|2021-04-16 | 20H (Beta, V2) |
| hCoV-19/South_Africa/NHLS-UCT-GP-6180/2021\|EPI_ISL_2375911\|2021-04-16 | 20H (Beta, V2) |
| hCoV-19/South_Africa/NHLS-UCT-GP-6204/2021\|EPI_ISL_2375912\|2021-04-20 | 20H (Beta, V2) |
| hCoV-19/South_Africa/NHLS-UCT-GP-6215/2021\|EPI_ISL_2375913\|2021-04-20 | 20H (Beta, V2) |
| hCoV-19/South_Africa/NHLS-UCT-GP-6221/2021\|EPI_ISL_2375914\|2021-04-22 | 20H (Beta, V2) |
| hCoV-19/South_Africa/NHLS-UCT-GP-6223/2021\|EPI_ISL_2375915\|2021-04-22 | 20H (Beta, V2) |
| hCoV-19/South_Africa/NHLS-UCT-GP-6228/2021\|EPI_ISL_2375916\|2021-04-15 | 20H (Beta, V2) |
| hCoV-19/South_Africa/NHLS-UCT-GP-6229/2021\|EPI_ISL_2375917\|2021-04-15 | 20H (Beta, V2) |
| hCoV-19/South_Africa/NHLS-UCT-GP-6234/2021\|EPI_ISL_2375918\|2021-04-17 | 20H (Beta, V2) |
| hCoV-19/South_Africa/NHLS-UCT-GP-6235/2021\|EPI_ISL_2375919\|2021-04-17 | 20H (Beta, V2) |
| hCoV-19/South_Africa/NHLS-UCT-GP-6265/2021\|EPI_ISL_2375920\|2021-04-22 | 20H (Beta, V2) |
| hCoV-19/South_Africa/NHLS-UCT-GP-6275/2021\|EPI_ISL_2375921\|2021-04-23 | 20H (Beta, V2) |
| hCoV-19/South_Africa/NHLS-UCT-GP-6317/2021\|EPI_ISL_2375923\|2021-04-24 | 20H (Beta, V2) |
| hCoV-19/South_Africa/NHLS-UCT-GP-6336/2021\|EPI_ISL_2375924\|2021-04-26 | 20B |
| hCoV-19/South_Africa/NHLS-UCT-GP-6362/2021\|EPI_ISL_2547340\|2021-05-03 | 20H (Beta, V2) |
| hCoV-19/South_Africa/NHLS-UCT-GP-6363/2021\|EPI_ISL_2547341\|2021-05-03 | 20H (Beta, V2) |
| hCoV-19/South_Africa/NHLS-UCT-GP-6364/2021\|EPI_ISL_2547342\|2021-05-03 | 20H (Beta, V2) |
| hCoV-19/South_Africa/NHLS-UCT-GP-6365/2021\|EPI_ISL_2547343\|2021-05-03 | 20H (Beta, V2) |
| hCoV-19/South_Africa/NHLS-UCT-GP-6366/2021\|EPI_ISL_2547344\|2021-05-04 | 20H (Beta, V2) |
| hCoV-19/South_Africa/NHLS-UCT-GP-6367/2021\|EPI_ISL_2547345\|2021-05-04 | 20H (Beta, V2) |
| hCoV-19/South_Africa/NHLS-UCT-GP-6368/2021\|EPI_ISL_2547346\|2021-05-04 | 20H (Beta, V2) |
| hCoV-19/South_Africa/NHLS-UCT-GP-6369/2021\|EPI_ISL_2547347\|2021-05-04 | 20H (Beta, V2) |
| hCoV-19/South_Africa/NHLS-UCT-GP-6370/2021\|EPI_ISL_2547348\|2021-05-04 | 20H (Beta, V2) |
| hCoV-19/South_Africa/NHLS-UCT-GP-6371/2021\|EPI_ISL_2547349\|2021-05-04 | 20H (Beta, V2) |
| hCoV-19/South_Africa/NHLS-UCT-GP-6372/2021\|EPI_ISL_2547350\|2021-05-04 | 20H (Beta, V2) |
| hCoV-19/South_Africa/NHLS-UCT-GP-6373/2021\|EPI_ISL_2547351\|2021-05-05 | 20H (Beta, V2) |
| hCoV-19/South_Africa/NHLS-UCT-GP-6374/2021\|EPI_ISL_2547352\|2021-05-06 | 20H (Beta, V2) |
| hCoV-19/South_Africa/NHLS-UCT-GP-6375/2021\|EPI_ISL_2547353\|2021-05-06 | 20H (Beta, V2) |
| hCoV-19/South_Africa/NHLS-UCT-GP-6376/2021\|EPI_ISL_2547354\|2021-05-06 | 20H (Beta, V2) |
| hCoV-19/South_Africa/NHLS-UCT-GP-6379/2021\|EPI_ISL_2547355\|2021-05-03 | 20I (Alpha, V1) |
| hCoV-19/South_Africa/NHLS-UCT-GP-6380/2021\|EPI_ISL_2547356\|2021-05-03 | 20H (Beta, V2) |
| hCoV-19/South_Africa/NHLS-UCT-GP-6386/2021\|EPI_ISL_2375925\|2021-05-03 | 20H (Beta, V2) |
| hCoV-19/South_Africa/NHLS-UCT-GP-6395/2021\|EPI_ISL_2375926\|2021-05-04 | 20H (Beta, V2) |
| hCoV-19/South_Africa/NHLS-UCT-GP-6415/2021\|EPI_ISL_2375927\|2021-05-04 | 20H (Beta, V2) |
| hCoV-19/South_Africa/NHLS-UCT-GP-6420/2021\|EPI_ISL_2375928\|2021-05-05 | 20H (Beta, V2) |
| hCoV-19/South_Africa/NHLS-UCT-GP-6421/2021\|EPI_ISL_2375929\|2021-05-05 | 20H (Beta, V2) |
| hCoV-19/South_Africa/NHLS-UCT-GP-6429/2021\|EPI_ISL_2547357\|2021-05-05 | 20H (Beta, V2) |
| hCoV-19/South_Africa/NHLS-UCT-GP-6430/2021\|EPI_ISL_2547358\|2021-05-05 | 20H (Beta, V2) |
| hCoV-19/South_Africa/NHLS-UCT-GP-6433/2021\|EPI_ISL_2547359\|2021-05-05 | 20H (Beta, V2) |
| hCoV-19/South_Africa/NHLS-UCT-GP-6440/2021\|EPI_ISL_2375930\|2021-05-06 | 20H (Beta, V2) |
| hCoV-19/South_Africa/NHLS-UCT-GP-6444/2021\|EPI_ISL_2375931\|2021-05-06 | 20H (Beta, V2) |
| hCoV-19/South_Africa/NHLS-UCT-GP-6446/2021\|EPI_ISL_2375932\|2021-05-06 | 20H (Beta, V2) |
| hCoV-19/South_Africa/NHLS-UCT-GP-6459/2021\|EPI_ISL_2547360\|2021-05-07 | 20H (Beta, V2) |
| hCoV-19/South_Africa/NHLS-UCT-GP-6460/2021\|EPI_ISL_2547361\|2021-05-07 | 20H (Beta, V2) |

| | |
|---|---|
| hCoV-19/South_Africa/NHLS-UCT-GP-6461/2021\|EPI_ISL_2547362\|2021-05-07 | 20H (Beta, V2) |
| hCoV-19/South_Africa/NHLS-UCT-GP-6473/2021\|EPI_ISL_2547363\|2021-05-07 | 20H (Beta, V2) |
| hCoV-19/South_Africa/NHLS-UCT-GP-6483/2021\|EPI_ISL_2547364\|2021-05-10 | 20H (Beta, V2) |
| hCoV-19/South_Africa/NHLS-UCT-GP-6499/2021\|EPI_ISL_2547365\|2021-05-12 | 20H (Beta, V2) |
| hCoV-19/South_Africa/NHLS-UCT-GP-6503/2021\|EPI_ISL_2547366\|2021-05-12 | 20H (Beta, V2) |
| hCoV-19/South_Africa/NHLS-UCT-GP-6506/2021\|EPI_ISL_2547367\|2021-05-12 | 20H (Beta, V2) |
| hCoV-19/South_Africa/NHLS-UCT-GP-6524/2021\|EPI_ISL_2547368\|2021-05-12 | 20H (Beta, V2) |
| hCoV-19/South_Africa/NHLS-UCT-GP-6525/2021\|EPI_ISL_2547369\|2021-05-13 | 20I (Alpha, V1) |
| hCoV-19/South_Africa/NHLS-UCT-GP-6526/2021\|EPI_ISL_2547370\|2021-05-12 | 20H (Beta, V2) |
| hCoV-19/South_Africa/NHLS-UCT-GP-6527/2021\|EPI_ISL_2547371\|2021-05-12 | 20H (Beta, V2) |
| hCoV-19/South_Africa/NHLS-UCT-GP-6528/2021\|EPI_ISL_2547372\|2021-05-10 | 20H (Beta, V2) |
| hCoV-19/South_Africa/NHLS-UCT-GP-6529/2021\|EPI_ISL_2621062\|2021-05 | 20H (Beta, V2) |
| hCoV-19/South_Africa/NHLS-UCT-GP-6535/2021\|EPI_ISL_2802107\|2021-05-25 | 20B |
| hCoV-19/South_Africa/NHLS-UCT-GP-6536/2021\|EPI_ISL_2802119\|2021-05-25 | 20I (Alpha, V1) |
| hCoV-19/South_Africa/NHLS-UCT-GP-6537/2021\|EPI_ISL_2802120\|2021-05-24 | 20H (Beta, V2) |
| hCoV-19/South_Africa/NHLS-UCT-GP-6538/2021\|EPI_ISL_2802108\|2021-05-25 | 20B |
| hCoV-19/South_Africa/NHLS-UCT-GP-6540/2021\|EPI_ISL_2802121\|2021-05-18 | 20H (Beta, V2) |
| hCoV-19/South_Africa/NHLS-UCT-GP-6541/2021\|EPI_ISL_2802122\|2021-05-27 | 20H (Beta, V2) |
| hCoV-19/South_Africa/NHLS-UCT-GP-6542/2021\|EPI_ISL_2802123\|2021-05-25 | 20I (Alpha, V1) |
| hCoV-19/South_Africa/NHLS-UCT-GP-6543/2021\|EPI_ISL_2802124\|2021-05-29 | 20H (Beta, V2) |
| hCoV-19/South_Africa/NHLS-UCT-GP-6544/2021\|EPI_ISL_2802188\|2021-06-02 | 20I (Alpha, V1) |
| hCoV-19/South_Africa/NHLS-UCT-GP-6546/2021\|EPI_ISL_2802190\|2021-06-01 | 21A (Delta) |
| hCoV-19/South_Africa/NHLS-UCT-GP-6548/2021\|EPI_ISL_2802125\|2021-05-24 | 21A (Delta) |
| hCoV-19/South_Africa/NHLS-UCT-GP-6549/2021\|EPI_ISL_2802126\|2021-05-24 | 21A (Delta) |
| hCoV-19/South_Africa/NHLS-UCT-GP-6550/2021\|EPI_ISL_2802127\|2021-05-24 | 21A (Delta) |
| hCoV-19/South_Africa/NHLS-UCT-GP-6555/2021\|EPI_ISL_3506441\|2021-05-24 | 21A (Delta) |
| hCoV-19/South_Africa/NHLS-UCT-GP-6556/2021\|EPI_ISL_3506442\|2021-05-24 | 21A (Delta) |
| hCoV-19/South_Africa/NHLS-UCT-GP-6557/2021\|EPI_ISL_3207499\|2021-05-27 | 20H (Beta, V2) |
| hCoV-19/South_Africa/NHLS-UCT-GP-6558/2021\|EPI_ISL_3207524\|2021-06-01 | 21A (Delta) |
| hCoV-19/South_Africa/NHLS-UCT-GP-6559/2021\|EPI_ISL_3207525\|2021-06-02 | 21A (Delta) |
| hCoV-19/South_Africa/NHLS-UCT-GP-6561/2021\|EPI_ISL_3207501\|2021-05-17 | 20H (Beta, V2) |
| hCoV-19/South_Africa/NHLS-UCT-GS-9424/2021\|EPI_ISL_1040659\|2021-01-01 | 20H (Beta, V2) |
| hCoV-19/South_Africa/NHLS-UCT-GS-9428/2021\|EPI_ISL_1534362\|2021-01-02 | 20H (Beta, V2) |
| hCoV-19/South_Africa/NHLS-UCT-GS-9435/2021\|EPI_ISL_1040658\|2021-01-02 | 20H (Beta, V2) |
| hCoV-19/South_Africa/NHLS-UCT-GS-9436/2021\|EPI_ISL_2140686\|2021-01-02 | 20H (Beta, V2) |
| hCoV-19/South_Africa/NHLS-UCT-GS-9438/2021\|EPI_ISL_1040646\|2021-01-02 | 20H (Beta, V2) |
| hCoV-19/South_Africa/NHLS-UCT-GS-9439/2021\|EPI_ISL_1534411\|2021-01-02 | 20H (Beta, V2) |
| hCoV-19/South_Africa/NHLS-UCT-GS-9449/2021\|EPI_ISL_1534413\|2021-01-04 | 20H (Beta, V2) |
| hCoV-19/South_Africa/NHLS-UCT-GS-9470/2021\|EPI_ISL_1534414\|2021-01-04 | 20H (Beta, V2) |
| hCoV-19/South_Africa/NHLS-UCT-GS-9471/2021\|EPI_ISL_1534415\|2021-01-04 | 20H (Beta, V2) |
| hCoV-19/South_Africa/NHLS-UCT-GS-9474/2021\|EPI_ISL_1040656\|2021-01-04 | 20H (Beta, V2) |
| hCoV-19/South_Africa/NHLS-UCT-GS-9475/2021\|EPI_ISL_2375979\|2021-01-04 | 20H (Beta, V2) |
| hCoV-19/South_Africa/NHLS-UCT-GS-9477/2021\|EPI_ISL_1040657\|2021-01-04 | 20H (Beta, V2) |
| hCoV-19/South_Africa/NHLS-UCT-GS-9484/2021\|EPI_ISL_1040660\|2021-01-05 | 20H (Beta, V2) |
| hCoV-19/South_Africa/NHLS-UCT-GS-9497/2021\|EPI_ISL_2140687\|2021-01-05 | 20H (Beta, V2) |
| hCoV-19/South_Africa/NHLS-UCT-GS-9504/2021\|EPI_ISL_2140688\|2021-01-05 | 20H (Beta, V2) |
| hCoV-19/South_Africa/NHLS-UCT-GS-9509/2021\|EPI_ISL_1534412\|2021-01-05 | 20H (Beta, V2) |
| hCoV-19/South_Africa/NHLS-UCT-GS-9510/2021\|EPI_ISL_2375980\|2021-01-06 | 20H (Beta, V2) |
| hCoV-19/South_Africa/NHLS-UCT-GS-9523/2021\|EPI_ISL_2621122\|2021-01-06 | 20H (Beta, V2) |
| hCoV-19/South_Africa/NHLS-UCT-GS-9527/2021\|EPI_ISL_2621123\|2021-01-06 | 20H (Beta, V2) |

| | |
|---|---|
| hCoV-19/South_Africa/NHLS-UCT-GS-9531/2021\|EPI_ISL_1534416\|2021-01-06 | 20H (Beta, V2) |
| hCoV-19/South_Africa/NHLS-UCT-GS-9534/2021\|EPI_ISL_2621124\|2021-01-06 | 20H (Beta, V2) |
| hCoV-19/South_Africa/NHLS-UCT-GS-9540/2021\|EPI_ISL_2621125\|2021-01-06 | 20B |
| hCoV-19/South_Africa/NHLS-UCT-GS-9542/2021\|EPI_ISL_2621126\|2021-01-07 | 20H (Beta, V2) |
| hCoV-19/South_Africa/NHLS-UCT-GS-9551/2021\|EPI_ISL_2621127\|2021-01-07 | 20H (Beta, V2) |
| hCoV-19/South_Africa/NHLS-UCT-GS-9553/2021\|EPI_ISL_2621128\|2021-01-07 | 20H (Beta, V2) |
| hCoV-19/South_Africa/NHLS-UCT-GS-9571/2021\|EPI_ISL_2375981\|2021-01-02 | 20H (Beta, V2) |
| hCoV-19/South_Africa/NHLS-UCT-GS-9581/2021\|EPI_ISL_1534381\|2021-01-02 | 20H (Beta, V2) |
| hCoV-19/South_Africa/NHLS-UCT-GS-9596/2021\|EPI_ISL_2375982\|2021-01-03 | 20H (Beta, V2) |
| hCoV-19/South_Africa/NHLS-UCT-GS-9617/2021\|EPI_ISL_1534375\|2021-01-03 | 20H (Beta, V2) |
| hCoV-19/South_Africa/NHLS-UCT-GS-9645/2021\|EPI_ISL_3207535\|2021-01-04 | 20H (Beta, V2) |
| hCoV-19/South_Africa/NHLS-UCT-GS-9649/2021\|EPI_ISL_1534417\|2021-01-04 | 20H (Beta, V2) |
| hCoV-19/South_Africa/NHLS-UCT-GS-9655/2021\|EPI_ISL_1534418\|2021-01-04 | 20H (Beta, V2) |
| hCoV-19/South_Africa/NHLS-UCT-GS-9661/2021\|EPI_ISL_1534460\|2021-01-04 | 20H (Beta, V2) |
| hCoV-19/South_Africa/NHLS-UCT-GS-9720/2021\|EPI_ISL_1534419\|2021-01-05 | 20H (Beta, V2) |
| hCoV-19/South_Africa/NHLS-UCT-GS-9730/2021\|EPI_ISL_1534420\|2021-01-06 | 20H (Beta, V2) |
| hCoV-19/South_Africa/NHLS-UCT-GS-9732/2021\|EPI_ISL_1534421\|2021-01-06 | 20H (Beta, V2) |
| hCoV-19/South_Africa/NHLS-UCT-GS-9754/2021\|EPI_ISL_1534384\|2021-01-06 | 20H (Beta, V2) |
| hCoV-19/South_Africa/NHLS-UCT-GS-9762/2021\|EPI_ISL_1534422\|2021-01-06 | 20H (Beta, V2) |
| hCoV-19/South_Africa/NHLS-UCT-GS-9812/2021\|EPI_ISL_1534423\|2021-01-05 | 20H (Beta, V2) |
| hCoV-19/South_Africa/NHLS-UCT-GS-9813/2021\|EPI_ISL_1534424\|2021-01-05 | 20H (Beta, V2) |
| hCoV-19/South_Africa/NHLS-UCT-GS-9818/2021\|EPI_ISL_1534425\|2021-01-05 | 20H (Beta, V2) |
| hCoV-19/South_Africa/NHLS-UCT-GS-9821/2021\|EPI_ISL_1040655\|2021-01-06 | 20H (Beta, V2) |
| hCoV-19/South_Africa/NHLS-UCT-GS-9825/2021\|EPI_ISL_1040662\|2021-01-01 | 20H (Beta, V2) |
| hCoV-19/South_Africa/NHLS-UCT-GS-9842/2021\|EPI_ISL_1534426\|2021-01-04 | 20H (Beta, V2) |
| hCoV-19/South_Africa/NHLS-UCT-GS-9852/2021\|EPI_ISL_1534427\|2021-01-05 | 20H (Beta, V2) |
| hCoV-19/South_Africa/NHLS-UCT-GS-9855/2021\|EPI_ISL_2375983\|2021-01-06 | 20H (Beta, V2) |
| hCoV-19/South_Africa/NHLS-UCT-GS-9876/2021\|EPI_ISL_1534383\|2021-01-04 | 20H (Beta, V2) |
| hCoV-19/South_Africa/NHLS-UCT-GS-9882/2021\|EPI_ISL_1534387\|2021-01-05 | 20H (Beta, V2) |
| hCoV-19/South_Africa/NHLS-UCT-GS-9890/2021\|EPI_ISL_3957791\|2021-01-06 | 20B |
| hCoV-19/South_Africa/NHLS-UCT-GS-9907/2021\|EPI_ISL_1706570\|2021-01-05 | 20H (Beta, V2) |
| hCoV-19/South_Africa/NHLS-UCT-GS-9911/2021\|EPI_ISL_1534389\|2021-01-09 | 20H (Beta, V2) |
| hCoV-19/South_Africa/NHLS-UCT-GS-9919/2021\|EPI_ISL_1040654\|2021-01-09 | 20H (Beta, V2) |
| hCoV-19/South_Africa/NHLS-UCT-GS-9930/2021\|EPI_ISL_2802173\|2021-01-11 | 20H (Beta, V2) |
| hCoV-19/South_Africa/NHLS-UCT-GS-9932/2021\|EPI_ISL_1040652\|2021-01-11 | 20H (Beta, V2) |
| hCoV-19/South_Africa/NHLS-UCT-GS-9945/2021\|EPI_ISL_1534390\|2021-01-11 | 20H (Beta, V2) |
| hCoV-19/South_Africa/NHLS-UCT-GS-9952/2021\|EPI_ISL_1040649\|2021-01-11 | 20H (Beta, V2) |
| hCoV-19/South_Africa/NHLS-UCT-GS-9956/2021\|EPI_ISL_2375984\|2021-01-11 | 20H (Beta, V2) |
| hCoV-19/South_Africa/NHLS-UCT-GS-9957/2021\|EPI_ISL_1706571\|2021-01-11 | 20H (Beta, V2) |
| hCoV-19/South_Africa/NHLS-UCT-GS-9961/2021\|EPI_ISL_1534376\|2021-01-11 | 20H (Beta, V2) |
| hCoV-19/South_Africa/NHLS-UCT-GS-9966/2021\|EPI_ISL_1040653\|2021-01-11 | 20H (Beta, V2) |
| hCoV-19/South_Africa/NHLS-UCT-GS-9969/2021\|EPI_ISL_1040661\|2021-01-11 | 20H (Beta, V2) |
| hCoV-19/South_Africa/NHLS-UCT-GS-9970/2021\|EPI_ISL_1040648\|2021-01-11 | 20H (Beta, V2) |
| hCoV-19/South_Africa/NHLS-UCT-GS-9975/2021\|EPI_ISL_1040663\|2021-01-11 | 20H (Beta, V2) |
| hCoV-19/South_Africa/NHLS-UCT-GS-9979/2021\|EPI_ISL_1040644\|2021-01-12 | 20H (Beta, V2) |
| hCoV-19/South_Africa/NHLS-UCT-GS-9982/2021\|EPI_ISL_1040650\|2021-01-12 | 20H (Beta, V2) |
| hCoV-19/South_Africa/NHLS-UCT-GS-9988/2021\|EPI_ISL_1040651\|2021-01-12 | 20H (Beta, V2) |
| hCoV-19/South_Africa/NHLS-UCT-GS-9998/2021\|EPI_ISL_1534391\|2021-01-12 | 20H (Beta, V2) |
| hCoV-19/South_Africa/NHLS-UCT-GS-A002/2021\|EPI_ISL_2140689\|2021-01-12 | 20H (Beta, V2) |
| hCoV-19/South_Africa/NHLS-UCT-GS-A004/2021\|EPI_ISL_1040645\|2021-01-12 | 20H (Beta, V2) |

| | |
|---|---|
| hCoV-19/South_Africa/NHLS-UCT-GS-A006/2021\|EPI_ISL_2375985\|2021-01-12 | 20H (Beta, V2) |
| hCoV-19/South_Africa/NHLS-UCT-GS-A016/2021\|EPI_ISL_2802174\|2021-01-13 | 20H (Beta, V2) |
| hCoV-19/South_Africa/NHLS-UCT-GS-A030/2021\|EPI_ISL_1534392\|2021-01-13 | 20H (Beta, V2) |
| hCoV-19/South_Africa/NHLS-UCT-GS-A042/2021\|EPI_ISL_2140690\|2021-01-14 | 20H (Beta, V2) |
| hCoV-19/South_Africa/NHLS-UCT-GS-A043/2021\|EPI_ISL_1534457\|2021-01-13 | 20H (Beta, V2) |
| hCoV-19/South_Africa/NHLS-UCT-GS-A046/2021\|EPI_ISL_2621129\|2021-01-13 | 20H (Beta, V2) |
| hCoV-19/South_Africa/NHLS-UCT-GS-A049/2021\|EPI_ISL_1534455\|2021-01-14 | 20H (Beta, V2) |
| hCoV-19/South_Africa/NHLS-UCT-GS-A078/2021\|EPI_ISL_1706576\|2021-01-15 | 20H (Beta, V2) |
| hCoV-19/South_Africa/NHLS-UCT-GS-A079/2021\|EPI_ISL_1534393\|2021-01-15 | 20H (Beta, V2) |
| hCoV-19/South_Africa/NHLS-UCT-GS-A085/2021\|EPI_ISL_1534394\|2021-01-15 | 20H (Beta, V2) |
| hCoV-19/South_Africa/NHLS-UCT-GS-A091/2021\|EPI_ISL_2375986\|2021-01-15 | 20H (Beta, V2) |
| hCoV-19/South_Africa/NHLS-UCT-GS-A108/2021\|EPI_ISL_1534380\|2021-01-10 | 20H (Beta, V2) |
| hCoV-19/South_Africa/NHLS-UCT-GS-A145/2021\|EPI_ISL_1534395\|2021-01-11 | 20H (Beta, V2) |
| hCoV-19/South_Africa/NHLS-UCT-GS-A153/2021\|EPI_ISL_1534396\|2021-01-12 | 20H (Beta, V2) |
| hCoV-19/South_Africa/NHLS-UCT-GS-A159/2021\|EPI_ISL_1534397\|2021-01-12 | 20H (Beta, V2) |
| hCoV-19/South_Africa/NHLS-UCT-GS-A172/2021\|EPI_ISL_1534398\|2021-01-11 | 20H (Beta, V2) |
| hCoV-19/South_Africa/NHLS-UCT-GS-A190/2021\|EPI_ISL_1534374\|2021-01-13 | 20H (Beta, V2) |
| hCoV-19/South_Africa/NHLS-UCT-GS-A204/2021\|EPI_ISL_1534399\|2021-01-13 | 20H (Beta, V2) |
| hCoV-19/South_Africa/NHLS-UCT-GS-A207/2021\|EPI_ISL_1534400\|2021-01-14 | 20H (Beta, V2) |
| hCoV-19/South_Africa/NHLS-UCT-GS-A211/2021\|EPI_ISL_1534401\|2021-01-14 | 20H (Beta, V2) |
| hCoV-19/South_Africa/NHLS-UCT-GS-A250/2021\|EPI_ISL_1534402\|2021-01-11 | 20H (Beta, V2) |
| hCoV-19/South_Africa/NHLS-UCT-GS-A259/2021\|EPI_ISL_1534403\|2021-01-12 | 20H (Beta, V2) |
| hCoV-19/South_Africa/NHLS-UCT-GS-A261/2021\|EPI_ISL_1534388\|2021-01-13 | 20H (Beta, V2) |
| hCoV-19/South_Africa/NHLS-UCT-GS-A281/2021\|EPI_ISL_1534404\|2021-01-11 | 20H (Beta, V2) |
| hCoV-19/South_Africa/NHLS-UCT-GS-A287/2021\|EPI_ISL_1534405\|2021-01-12 | 20H (Beta, V2) |
| hCoV-19/South_Africa/NHLS-UCT-GS-A296/2021\|EPI_ISL_1534456\|2021-01-13 | 20H (Beta, V2) |
| hCoV-19/South_Africa/NHLS-UCT-GS-A303/2021\|EPI_ISL_1817684\|2021-01-14 | 20H (Beta, V2) |
| hCoV-19/South_Africa/NHLS-UCT-GS-A354/2021\|EPI_ISL_1534406\|2021-01-19 | 20H (Beta, V2) |
| hCoV-19/South_Africa/NHLS-UCT-GS-A356/2021\|EPI_ISL_1534407\|2021-01-19 | 20H (Beta, V2) |
| hCoV-19/South_Africa/NHLS-UCT-GS-A372/2021\|EPI_ISL_1534408\|2021-01-20 | 20H (Beta, V2) |
| hCoV-19/South_Africa/NHLS-UCT-GS-A395/2021\|EPI_ISL_1534377\|2021-01-22 | 20H (Beta, V2) |
| hCoV-19/South_Africa/NHLS-UCT-GS-A396/2021\|EPI_ISL_1534459\|2021-01-22 | 20H (Beta, V2) |
| hCoV-19/South_Africa/NHLS-UCT-GS-A440/2021\|EPI_ISL_1534409\|2021-01-18 | 20H (Beta, V2) |
| hCoV-19/South_Africa/NHLS-UCT-GS-A444/2021\|EPI_ISL_1534410\|2021-01-18 | 20H (Beta, V2) |
| hCoV-19/South_Africa/NHLS-UCT-GS-A454/2021\|EPI_ISL_3207536\|2021-01-18 | 20H (Beta, V2) |
| hCoV-19/South_Africa/NHLS-UCT-GS-A495/2021\|EPI_ISL_1534386\|2021-01-19 | 20H (Beta, V2) |
| hCoV-19/South_Africa/NHLS-UCT-GS-A502/2021\|EPI_ISL_1534436\|2021-01-19 | 20H (Beta, V2) |
| hCoV-19/South_Africa/NHLS-UCT-GS-A504/2021\|EPI_ISL_1534379\|2021-01-18 | 20H (Beta, V2) |
| hCoV-19/South_Africa/NHLS-UCT-GS-A505/2021\|EPI_ISL_1706572\|2021-01-19 | 20H (Beta, V2) |
| hCoV-19/South_Africa/NHLS-UCT-GS-A506/2021\|EPI_ISL_1534437\|2021-01-15 | 20H (Beta, V2) |
| hCoV-19/South_Africa/NHLS-UCT-GS-A510/2021\|EPI_ISL_1534438\|2021-01-19 | 20H (Beta, V2) |
| hCoV-19/South_Africa/NHLS-UCT-GS-A522/2021\|EPI_ISL_1534439\|2021-01-20 | 20H (Beta, V2) |
| hCoV-19/South_Africa/NHLS-UCT-GS-A523/2021\|EPI_ISL_1534440\|2021-01-20 | 20H (Beta, V2) |
| hCoV-19/South_Africa/NHLS-UCT-GS-A542/2021\|EPI_ISL_1534441\|2021-01-22 | 20H (Beta, V2) |
| hCoV-19/South_Africa/NHLS-UCT-GS-A543/2021\|EPI_ISL_1534442\|2021-01-22 | 20H (Beta, V2) |
| hCoV-19/South_Africa/NHLS-UCT-GS-A546/2021\|EPI_ISL_1706573\|2021-01-18 | 20H (Beta, V2) |
| hCoV-19/South_Africa/NHLS-UCT-GS-A548/2021\|EPI_ISL_1534443\|2021-01-19 | 20H (Beta, V2) |
| hCoV-19/South_Africa/NHLS-UCT-GS-A567/2021\|EPI_ISL_1534444\|2021-01-21 | 20H (Beta, V2) |
| hCoV-19/South_Africa/NHLS-UCT-GS-A568/2021\|EPI_ISL_1534445\|2021-01-22 | 20H (Beta, V2) |
| hCoV-19/South_Africa/NHLS-UCT-GS-A571/2021\|EPI_ISL_1534446\|2021-01-17 | 20H (Beta, V2) |

| | |
|---|---|
| hCoV-19/South_Africa/NHLS-UCT-GS-A572/2021\|EPI_ISL_1534447\|2021-01-17 | 20H (Beta, V2) |
| hCoV-19/South_Africa/NHLS-UCT-GS-A582/2021\|EPI_ISL_1534448\|2021-01-21 | 20H (Beta, V2) |
| hCoV-19/South_Africa/NHLS-UCT-GS-A591/2021\|EPI_ISL_1534385\|2021-01-18 | 20H (Beta, V2) |
| hCoV-19/South_Africa/NHLS-UCT-GS-A607/2021\|EPI_ISL_1534378\|2021-01-20 | 20H (Beta, V2) |
| hCoV-19/South_Africa/NHLS-UCT-GS-A614/2021\|EPI_ISL_1534449\|2021-01-22 | 20H (Beta, V2) |
| hCoV-19/South_Africa/NHLS-UCT-GS-A615/2021\|EPI_ISL_1706574\|2021-01-22 | 20H (Beta, V2) |
| hCoV-19/South_Africa/NHLS-UCT-GS-A638/2021\|EPI_ISL_1534450\|2021-01-25 | 20H (Beta, V2) |
| hCoV-19/South_Africa/NHLS-UCT-GS-A639/2021\|EPI_ISL_1534451\|2021-01-25 | 20H (Beta, V2) |
| hCoV-19/South_Africa/NHLS-UCT-GS-A647/2021\|EPI_ISL_1534452\|2021-01-26 | 20H (Beta, V2) |
| hCoV-19/South_Africa/NHLS-UCT-GS-A648/2021\|EPI_ISL_1534382\|2021-01-26 | 20H (Beta, V2) |
| hCoV-19/South_Africa/NHLS-UCT-GS-A656/2021\|EPI_ISL_1534453\|2021-01-27 | 20H (Beta, V2) |
| hCoV-19/South_Africa/NHLS-UCT-GS-A660/2021\|EPI_ISL_1706575\|2021-01-22 | 20H (Beta, V2) |
| hCoV-19/South_Africa/NHLS-UCT-GS-A664/2021\|EPI_ISL_1534454\|2021-01-22 | 20H (Beta, V2) |
| hCoV-19/South_Africa/NHLS-UCT-GS-A690/2021\|EPI_ISL_1534428\|2021-01-25 | 20H (Beta, V2) |
| hCoV-19/South_Africa/NHLS-UCT-GS-A717/2021\|EPI_ISL_1534429\|2021-01-27 | 20H (Beta, V2) |
| hCoV-19/South_Africa/NHLS-UCT-GS-A723/2021\|EPI_ISL_1534430\|2021-01-27 | 20H (Beta, V2) |
| hCoV-19/South_Africa/NHLS-UCT-GS-A730/2021\|EPI_ISL_1534461\|2021-01-28 | 20H (Beta, V2) |
| hCoV-19/South_Africa/NHLS-UCT-GS-A745/2021\|EPI_ISL_1534431\|2021-01-25 | 20H (Beta, V2) |
| hCoV-19/South_Africa/NHLS-UCT-GS-A752/2021\|EPI_ISL_1534432\|2021-01-25 | 20H (Beta, V2) |
| hCoV-19/South_Africa/NHLS-UCT-GS-A753/2021\|EPI_ISL_1534433\|2021-01-26 | 20H (Beta, V2) |
| hCoV-19/South_Africa/NHLS-UCT-GS-A755/2021\|EPI_ISL_1534434\|2021-01-26 | 20H (Beta, V2) |
| hCoV-19/South_Africa/NHLS-UCT-GS-A761/2021\|EPI_ISL_1534458\|2021-01-27 | 20H (Beta, V2) |
| hCoV-19/South_Africa/NHLS-UCT-GS-A769/2021\|EPI_ISL_1534435\|2021-01-23 | 20H (Beta, V2) |
| hCoV-19/South_Africa/NHLS-UCT-GS-A800/2021\|EPI_ISL_1706562\|2021-02-02 | 20H (Beta, V2) |
| hCoV-19/South_Africa/NHLS-UCT-GS-A801/2021\|EPI_ISL_1534364\|2021-02-03 | 20A |
| hCoV-19/South_Africa/NHLS-UCT-GS-A802/2021\|EPI_ISL_1706563\|2021-02-03 | 20H (Beta, V2) |
| hCoV-19/South_Africa/NHLS-UCT-GS-A804/2021\|EPI_ISL_1706564\|2021-02-03 | 20H (Beta, V2) |
| hCoV-19/South_Africa/NHLS-UCT-GS-A805/2021\|EPI_ISL_1706565\|2021-02-04 | 20H (Beta, V2) |
| hCoV-19/South_Africa/NHLS-UCT-GS-A806/2021\|EPI_ISL_1706566\|2021-02-01 | 20H (Beta, V2) |
| hCoV-19/South_Africa/NHLS-UCT-GS-A807/2021\|EPI_ISL_1706567\|2021-02-02 | 20H (Beta, V2) |
| hCoV-19/South_Africa/NHLS-UCT-GS-A808/2021\|EPI_ISL_1534365\|2021-02-01 | 20H (Beta, V2) |
| hCoV-19/South_Africa/NHLS-UCT-GS-A809/2021\|EPI_ISL_1534366\|2021-02-01 | 20H (Beta, V2) |
| hCoV-19/South_Africa/NHLS-UCT-GS-A811/2021\|EPI_ISL_1534367\|2021-02-01 | 20H (Beta, V2) |
| hCoV-19/South_Africa/NHLS-UCT-GS-A812/2021\|EPI_ISL_1534368\|2021-02-02 | 20H (Beta, V2) |
| hCoV-19/South_Africa/NHLS-UCT-GS-A813/2021\|EPI_ISL_1534369\|2021-02-02 | 20H (Beta, V2) |
| hCoV-19/South_Africa/NHLS-UCT-GS-A814/2021\|EPI_ISL_1534370\|2021-02-05 | 20H (Beta, V2) |
| hCoV-19/South_Africa/NHLS-UCT-GS-A815/2021\|EPI_ISL_1706568\|2021-02-03 | 20H (Beta, V2) |
| hCoV-19/South_Africa/NHLS-UCT-GS-A816/2021\|EPI_ISL_1534371\|2021-02-02 | 20H (Beta, V2) |
| hCoV-19/South_Africa/NHLS-UCT-GS-A817/2021\|EPI_ISL_1706569\|2021-02-04 | 20H (Beta, V2) |
| hCoV-19/South_Africa/NHLS-UCT-GS-A818/2021\|EPI_ISL_1534372\|2021-02-03 | 20H (Beta, V2) |
| hCoV-19/South_Africa/NHLS-UCT-GS-A820/2021\|EPI_ISL_1534373\|2021-01-21 | 20H (Beta, V2) |
| hCoV-19/South_Africa/NHLS-UCT-GS-A826/2021\|EPI_ISL_2802175\|2021-01-31 | 20H (Beta, V2) |
| hCoV-19/South_Africa/NHLS-UCT-GS-A839/2021\|EPI_ISL_1534310\|2021-02-03 | 20H (Beta, V2) |
| hCoV-19/South_Africa/NHLS-UCT-GS-A868/2021\|EPI_ISL_1534245\|2021-02-05 | 20H (Beta, V2) |
| hCoV-19/South_Africa/NHLS-UCT-GS-A887/2021\|EPI_ISL_1534246\|2021-02-01 | 20H (Beta, V2) |
| hCoV-19/South_Africa/NHLS-UCT-GS-A894/2021\|EPI_ISL_1817752\|2021-02-03 | 20B |
| hCoV-19/South_Africa/NHLS-UCT-GS-A895/2021\|EPI_ISL_1534247\|2021-02-08 | 20H (Beta, V2) |
| hCoV-19/South_Africa/NHLS-UCT-GS-A896/2021\|EPI_ISL_1534248\|2021-02-08 | 20H (Beta, V2) |
| hCoV-19/South_Africa/NHLS-UCT-GS-A897/2021\|EPI_ISL_1534249\|2021-02-09 | 20H (Beta, V2) |
| hCoV-19/South_Africa/NHLS-UCT-GS-A898/2021\|EPI_ISL_1534250\|2021-02-09 | 20H (Beta, V2) |

| | |
|---|---|
| hCoV-19/South_Africa/NHLS-UCT-GS-A899/2021\|EPI_ISL_1534251\|2021-02-09 | 20H (Beta, V2) |
| hCoV-19/South_Africa/NHLS-UCT-GS-A900/2021\|EPI_ISL_1534252\|2021-02-09 | 20H (Beta, V2) |
| hCoV-19/South_Africa/NHLS-UCT-GS-A904/2021\|EPI_ISL_1534253\|2021-02-10 | 20H (Beta, V2) |
| hCoV-19/South_Africa/NHLS-UCT-GS-A907/2021\|EPI_ISL_1534254\|2021-02-11 | 20H (Beta, V2) |
| hCoV-19/South_Africa/NHLS-UCT-GS-A908/2021\|EPI_ISL_1534255\|2021-02-11 | 20H (Beta, V2) |
| hCoV-19/South_Africa/NHLS-UCT-GS-A909/2021\|EPI_ISL_1534256\|2021-02-12 | 20H (Beta, V2) |
| hCoV-19/South_Africa/NHLS-UCT-GS-A910/2021\|EPI_ISL_1534257\|2021-02-12 | 20H (Beta, V2) |
| hCoV-19/South_Africa/NHLS-UCT-GS-A912/2021\|EPI_ISL_1534258\|2021-02-12 | 20H (Beta, V2) |
| hCoV-19/South_Africa/NHLS-UCT-GS-A913/2021\|EPI_ISL_1534259\|2021-02-12 | 20H (Beta, V2) |
| hCoV-19/South_Africa/NHLS-UCT-GS-A920/2021\|EPI_ISL_1534260\|2021-02-08 | 20H (Beta, V2) |
| hCoV-19/South_Africa/NHLS-UCT-GS-A921/2021\|EPI_ISL_1817692\|2021-02-08 | 20H (Beta, V2) |
| hCoV-19/South_Africa/NHLS-UCT-GS-A923/2021\|EPI_ISL_1534261\|2021-02-08 | 20H (Beta, V2) |
| hCoV-19/South_Africa/NHLS-UCT-GS-A924/2021\|EPI_ISL_1534262\|2021-02-08 | 20H (Beta, V2) |
| hCoV-19/South_Africa/NHLS-UCT-GS-A925/2021\|EPI_ISL_1534263\|2021-02-08 | 20H (Beta, V2) |
| hCoV-19/South_Africa/NHLS-UCT-GS-A926/2021\|EPI_ISL_1534264\|2021-02-08 | 20H (Beta, V2) |
| hCoV-19/South_Africa/NHLS-UCT-GS-A928/2021\|EPI_ISL_1534265\|2021-02-09 | 20H (Beta, V2) |
| hCoV-19/South_Africa/NHLS-UCT-GS-A929/2021\|EPI_ISL_1534266\|2021-02-09 | 20H (Beta, V2) |
| hCoV-19/South_Africa/NHLS-UCT-GS-A930/2021\|EPI_ISL_1534267\|2021-02-09 | 20H (Beta, V2) |
| hCoV-19/South_Africa/NHLS-UCT-GS-B010/2021\|EPI_ISL_2375987\|2021-01-13 | 20H (Beta, V2) |
| hCoV-19/South_Africa/NHLS-UCT-GS-B050/2021\|EPI_ISL_2802176\|2021-01-04 | 20H (Beta, V2) |
| hCoV-19/South_Africa/NHLS-UCT-GS-B053/2021\|EPI_ISL_2375988\|2021-01-03 | 20H (Beta, V2) |
| hCoV-19/South_Africa/NHLS-UCT-GS-B086/2021\|EPI_ISL_2621130\|2021-01-04 | 20H (Beta, V2) |
| hCoV-19/South_Africa/NHLS-UCT-GS-B147/2021\|EPI_ISL_1817694\|2021-02-15 | 20H (Beta, V2) |
| hCoV-19/South_Africa/NHLS-UCT-GS-B151/2021\|EPI_ISL_1534268\|2021-02-16 | 20H (Beta, V2) |
| hCoV-19/South_Africa/NHLS-UCT-GS-B158/2021\|EPI_ISL_1817665\|2021-02-19 | 20H (Beta, V2) |
| hCoV-19/South_Africa/NHLS-UCT-GS-B177/2021\|EPI_ISL_1534269\|2021-02-15 | 20H (Beta, V2) |
| hCoV-19/South_Africa/NHLS-UCT-GS-B179/2021\|EPI_ISL_1534270\|2021-02-15 | 20H (Beta, V2) |
| hCoV-19/South_Africa/NHLS-UCT-GS-B181/2021\|EPI_ISL_1817706\|2021-02-15 | 20H (Beta, V2) |
| hCoV-19/South_Africa/NHLS-UCT-GS-B182/2021\|EPI_ISL_1817688\|2021-02-15 | 20H (Beta, V2) |
| hCoV-19/South_Africa/NHLS-UCT-GS-B188/2021\|EPI_ISL_1534311\|2021-02-16 | 20H (Beta, V2) |
| hCoV-19/South_Africa/NHLS-UCT-GS-B190/2021\|EPI_ISL_1817674\|2021-02-17 | 20H (Beta, V2) |
| hCoV-19/South_Africa/NHLS-UCT-GS-B191/2021\|EPI_ISL_1534271\|2021-02-17 | 20H (Beta, V2) |
| hCoV-19/South_Africa/NHLS-UCT-GS-B192/2021\|EPI_ISL_1534272\|2021-02-17 | 20H (Beta, V2) |
| hCoV-19/South_Africa/NHLS-UCT-GS-B198/2021\|EPI_ISL_1534273\|2021-02-18 | 20H (Beta, V2) |
| hCoV-19/South_Africa/NHLS-UCT-GS-B199/2021\|EPI_ISL_1534274\|2021-02-18 | 20H (Beta, V2) |
| hCoV-19/South_Africa/NHLS-UCT-GS-B202/2021\|EPI_ISL_1534275\|2021-02-18 | 20H (Beta, V2) |
| hCoV-19/South_Africa/NHLS-UCT-GS-B207/2021\|EPI_ISL_1817680\|2021-02-16 | 20H (Beta, V2) |
| hCoV-19/South_Africa/NHLS-UCT-GS-B208/2021\|EPI_ISL_1534276\|2021-02-15 | 20H (Beta, V2) |
| hCoV-19/South_Africa/NHLS-UCT-GS-B210/2021\|EPI_ISL_1534277\|2021-02-16 | 20H (Beta, V2) |
| hCoV-19/South_Africa/NHLS-UCT-GS-B214/2021\|EPI_ISL_1534278\|2021-02-25 | 20H (Beta, V2) |
| hCoV-19/South_Africa/NHLS-UCT-GS-B215/2021\|EPI_ISL_1534279\|2021-03-02 | 20H (Beta, V2) |
| hCoV-19/South_Africa/NHLS-UCT-GS-B216/2021\|EPI_ISL_1534280\|2021-02-22 | 20H (Beta, V2) |
| hCoV-19/South_Africa/NHLS-UCT-GS-B217/2021\|EPI_ISL_1534281\|2021-03-03 | 20H (Beta, V2) |
| hCoV-19/South_Africa/NHLS-UCT-GS-B218/2021\|EPI_ISL_1534282\|2021-03-02 | 20H (Beta, V2) |
| hCoV-19/South_Africa/NHLS-UCT-GS-B219/2021\|EPI_ISL_1534283\|2021-02-23 | 20H (Beta, V2) |
| hCoV-19/South_Africa/NHLS-UCT-GS-B221/2021\|EPI_ISL_1534284\|2021-02-22 | 20H (Beta, V2) |
| hCoV-19/South_Africa/NHLS-UCT-GS-B222/2021\|EPI_ISL_1534285\|2021-02-19 | 20H (Beta, V2) |
| hCoV-19/South_Africa/NHLS-UCT-GS-B223/2021\|EPI_ISL_1534286\|2021-03-02 | 20H (Beta, V2) |
| hCoV-19/South_Africa/NHLS-UCT-GS-B224/2021\|EPI_ISL_1534287\|2021-03-05 | 20H (Beta, V2) |
| hCoV-19/South_Africa/NHLS-UCT-GS-B225/2021\|EPI_ISL_1534288\|2021-02-25 | 20H (Beta, V2) |

| | |
|---|---|
| hCoV-19/South_Africa/NHLS-UCT-GS-B226/2021\|EPI_ISL_1534289\|2021-03-04 | 20H (Beta, V2) |
| hCoV-19/South_Africa/NHLS-UCT-GS-B227/2021\|EPI_ISL_1534290\|2021-02-25 | 20H (Beta, V2) |
| hCoV-19/South_Africa/NHLS-UCT-GS-B228/2021\|EPI_ISL_1534291\|2021-02-24 | 20H (Beta, V2) |
| hCoV-19/South_Africa/NHLS-UCT-GS-B229/2021\|EPI_ISL_1534292\|2021-03-02 | 20H (Beta, V2) |
| hCoV-19/South_Africa/NHLS-UCT-GS-B231/2021\|EPI_ISL_1817736\|2021-02-24 | 20H (Beta, V2) |
| hCoV-19/South_Africa/NHLS-UCT-GS-B232/2021\|EPI_ISL_1534293\|2021-02-25 | 20H (Beta, V2) |
| hCoV-19/South_Africa/NHLS-UCT-GS-B234/2021\|EPI_ISL_1534294\|2021-02-22 | 20H (Beta, V2) |
| hCoV-19/South_Africa/NHLS-UCT-GS-B235/2021\|EPI_ISL_1534295\|2021-02-20 | 20H (Beta, V2) |
| hCoV-19/South_Africa/NHLS-UCT-GS-B236/2021\|EPI_ISL_1534296\|2021-02-23 | 20H (Beta, V2) |
| hCoV-19/South_Africa/NHLS-UCT-GS-B237/2021\|EPI_ISL_1534297\|2021-02-22 | 20H (Beta, V2) |
| hCoV-19/South_Africa/NHLS-UCT-GS-B238/2021\|EPI_ISL_1534298\|2021-02-23 | 20H (Beta, V2) |
| hCoV-19/South_Africa/NHLS-UCT-GS-B239/2021\|EPI_ISL_1817690\|2021-02-22 | 20H (Beta, V2) |
| hCoV-19/South_Africa/NHLS-UCT-GS-B241/2021\|EPI_ISL_1534299\|2021-03-04 | 20H (Beta, V2) |
| hCoV-19/South_Africa/NHLS-UCT-GS-B242/2021\|EPI_ISL_1534300\|2021-02-23 | 20H (Beta, V2) |
| hCoV-19/South_Africa/NHLS-UCT-GS-B243/2021\|EPI_ISL_1534301\|2021-02-19 | 20H (Beta, V2) |
| hCoV-19/South_Africa/NHLS-UCT-GS-B244/2021\|EPI_ISL_1534302\|2021-02-20 | 20H (Beta, V2) |
| hCoV-19/South_Africa/NHLS-UCT-GS-B246/2021\|EPI_ISL_1534303\|2021-03-01 | 20H (Beta, V2) |
| hCoV-19/South_Africa/NHLS-UCT-GS-B248/2021\|EPI_ISL_1534304\|2021-02-25 | 20H (Beta, V2) |
| hCoV-19/South_Africa/NHLS-UCT-GS-B249/2021\|EPI_ISL_1534305\|2021-02-22 | 20H (Beta, V2) |
| hCoV-19/South_Africa/NHLS-UCT-GS-B250/2021\|EPI_ISL_1817691\|2021-03-04 | 20H (Beta, V2) |
| hCoV-19/South_Africa/NHLS-UCT-GS-B251/2021\|EPI_ISL_1817687\|2021-02-23 | 20H (Beta, V2) |
| hCoV-19/South_Africa/NHLS-UCT-GS-B252/2021\|EPI_ISL_1534306\|2021-02-23 | 20H (Beta, V2) |
| hCoV-19/South_Africa/NHLS-UCT-GS-B253/2021\|EPI_ISL_1534312\|2021-03-01 | 20H (Beta, V2) |
| hCoV-19/South_Africa/NHLS-UCT-GS-B254/2021\|EPI_ISL_1817671\|2021-03-02 | 20H (Beta, V2) |
| hCoV-19/South_Africa/NHLS-UCT-GS-B255/2021\|EPI_ISL_1817663\|2021-03-03 | 20H (Beta, V2) |
| hCoV-19/South_Africa/NHLS-UCT-GS-B256/2021\|EPI_ISL_1534307\|2021-02-26 | 20H (Beta, V2) |
| hCoV-19/South_Africa/NHLS-UCT-GS-B257/2021\|EPI_ISL_1534308\|2021-03-01 | 20H (Beta, V2) |
| hCoV-19/South_Africa/NHLS-UCT-GS-B258/2021\|EPI_ISL_1534309\|2021-03-01 | 20H (Beta, V2) |
| hCoV-19/South_Africa/NHLS-UCT-GS-B260/2021\|EPI_ISL_1817673\|2021-03-05 | 20H (Beta, V2) |
| hCoV-19/South_Africa/NHLS-UCT-GS-B261/2021\|EPI_ISL_1817707\|2021-02-22 | 20H (Beta, V2) |
| hCoV-19/South_Africa/NHLS-UCT-GS-B300/2021\|EPI_ISL_1817744\|2021-03-06 | 20H (Beta, V2) |
| hCoV-19/South_Africa/NHLS-UCT-GS-B313/2021\|EPI_ISL_1817681\|2021-03-11 | 20H (Beta, V2) |
| hCoV-19/South_Africa/NHLS-UCT-GS-B319/2021\|EPI_ISL_1817737\|2021-03-08 | 20H (Beta, V2) |
| hCoV-19/South_Africa/NHLS-UCT-GS-B320/2021\|EPI_ISL_1817710\|2021-03-08 | 20H (Beta, V2) |
| hCoV-19/South_Africa/NHLS-UCT-GS-B322/2021\|EPI_ISL_1817711\|2021-03-08 | 20H (Beta, V2) |
| hCoV-19/South_Africa/NHLS-UCT-GS-B324/2021\|EPI_ISL_1817712\|2021-03-08 | 20H (Beta, V2) |
| hCoV-19/South_Africa/NHLS-UCT-GS-B325/2021\|EPI_ISL_1817715\|2021-03-09 | 20H (Beta, V2) |
| hCoV-19/South_Africa/NHLS-UCT-GS-B326/2021\|EPI_ISL_1817733\|2021-03-10 | 20H (Beta, V2) |
| hCoV-19/South_Africa/NHLS-UCT-GS-B328/2021\|EPI_ISL_1817735\|2021-03-06 | 20H (Beta, V2) |
| hCoV-19/South_Africa/NHLS-UCT-GS-B335/2021\|EPI_ISL_1817667\|2021-03-15 | 20H (Beta, V2) |
| hCoV-19/South_Africa/NHLS-UCT-GS-B338/2021\|EPI_ISL_1817738\|2021-03-14 | 20H (Beta, V2) |
| hCoV-19/South_Africa/NHLS-UCT-GS-B341/2021\|EPI_ISL_1817739\|2021-03-15 | 20H (Beta, V2) |
| hCoV-19/South_Africa/NHLS-UCT-GS-B343/2021\|EPI_ISL_1817740\|2021-03-15 | 20H (Beta, V2) |
| hCoV-19/South_Africa/NHLS-UCT-GS-B346/2021\|EPI_ISL_1817741\|2021-03-17 | 20H (Beta, V2) |
| hCoV-19/South_Africa/NHLS-UCT-GS-B351/2021\|EPI_ISL_1817734\|2021-03-17 | 20H (Beta, V2) |
| hCoV-19/South_Africa/NHLS-UCT-GS-B352/2021\|EPI_ISL_1817670\|2021-03-17 | 20H (Beta, V2) |
| hCoV-19/South_Africa/NHLS-UCT-GS-B356/2021\|EPI_ISL_1817669\|2021-03-15 | 20H (Beta, V2) |
| hCoV-19/South_Africa/NHLS-UCT-GS-B360/2021\|EPI_ISL_1817745\|2021-03-23 | 20H (Beta, V2) |
| hCoV-19/South_Africa/NHLS-UCT-GS-B361/2021\|EPI_ISL_1817746\|2021-03-23 | 20H (Beta, V2) |
| hCoV-19/South_Africa/NHLS-UCT-GS-B363/2021\|EPI_ISL_1817702\|2021-03-23 | 20H (Beta, V2) |

| | |
|---|---|
| hCoV-19/South_Africa/NHLS-UCT-GS-B367/2021\|EPI_ISL_1817747\|2021-03-24 | 20H (Beta, V2) |
| hCoV-19/South_Africa/NHLS-UCT-GS-B368/2021\|EPI_ISL_1817748\|2021-03-25 | 20H (Beta, V2) |
| hCoV-19/South_Africa/NHLS-UCT-GS-B369/2021\|EPI_ISL_1817749\|2021-03-25 | 20H (Beta, V2) |
| hCoV-19/South_Africa/NHLS-UCT-GS-B371/2021\|EPI_ISL_1817703\|2021-03-25 | 20H (Beta, V2) |
| hCoV-19/South_Africa/NHLS-UCT-GS-B372/2021\|EPI_ISL_1817704\|2021-03-25 | 20H (Beta, V2) |
| hCoV-19/South_Africa/NHLS-UCT-GS-B376/2021\|EPI_ISL_1817675\|2021-03-19 | 20H (Beta, V2) |
| hCoV-19/South_Africa/NHLS-UCT-GS-B379/2021\|EPI_ISL_1817742\|2021-03-20 | 20H (Beta, V2) |
| hCoV-19/South_Africa/NHLS-UCT-GS-B380/2021\|EPI_ISL_1817743\|2021-03-20 | 20H (Beta, V2) |
| hCoV-19/South_Africa/NHLS-UCT-GS-B384/2021\|EPI_ISL_1817701\|2021-03-22 | 20H (Beta, V2) |
| hCoV-19/South_Africa/NHLS-UCT-GS-B403/2021\|EPI_ISL_1817679\|2021-03-23 | 20H (Beta, V2) |
| hCoV-19/South_Africa/NHLS-UCT-GS-B409/2021\|EPI_ISL_1817689\|2021-03-26 | 20H (Beta, V2) |
| hCoV-19/South_Africa/NHLS-UCT-GS-B415/2021\|EPI_ISL_1817664\|2021-03-17 | 20H (Beta, V2) |
| hCoV-19/South_Africa/NHLS-UCT-GS-B416/2021\|EPI_ISL_1817662\|2021-03-17 | 20H (Beta, V2) |
| hCoV-19/South_Africa/NHLS-UCT-GS-B417/2021\|EPI_ISL_1817682\|2021-03-17 | 20H (Beta, V2) |
| hCoV-19/South_Africa/NHLS-UCT-GS-B421/2021\|EPI_ISL_1817709\|2021-03-17 | 20H (Beta, V2) |
| hCoV-19/South_Africa/NHLS-UCT-GS-B430/2021\|EPI_ISL_2140691\|2021-04-12 | 20H (Beta, V2) |
| hCoV-19/South_Africa/NHLS-UCT-GS-B431/2021\|EPI_ISL_3207493\|2021-04-13 | 20H (Beta, V2) |
| hCoV-19/South_Africa/NHLS-UCT-GS-B438/2021\|EPI_ISL_2140692\|2021-04-11 | 20H (Beta, V2) |
| hCoV-19/South_Africa/NHLS-UCT-GS-B441/2021\|EPI_ISL_2140693\|2021-04-11 | 20H (Beta, V2) |
| hCoV-19/South_Africa/NHLS-UCT-GS-B445/2021\|EPI_ISL_2140694\|2021-04-13 | 20H (Beta, V2) |
| hCoV-19/South_Africa/NHLS-UCT-GS-B448/2021\|EPI_ISL_2140695\|2021-04-14 | 20H (Beta, V2) |
| hCoV-19/South_Africa/NHLS-UCT-GS-B449/2021\|EPI_ISL_2140696\|2021-04-14 | 20H (Beta, V2) |
| hCoV-19/South_Africa/NHLS-UCT-GS-B461/2021\|EPI_ISL_2140697\|2021-04-12 | 20H (Beta, V2) |
| hCoV-19/South_Africa/NHLS-UCT-GS-B467/2021\|EPI_ISL_2140698\|2021-04-14 | 20H (Beta, V2) |
| hCoV-19/South_Africa/NHLS-UCT-GS-B473/2021\|EPI_ISL_2140699\|2021-04-14 | 20H (Beta, V2) |
| hCoV-19/South_Africa/NHLS-UCT-GS-B475/2021\|EPI_ISL_2140700\|2021-03-27 | 20H (Beta, V2) |
| hCoV-19/South_Africa/NHLS-UCT-GS-B476/2021\|EPI_ISL_2140701\|2021-03-29 | 20H (Beta, V2) |
| hCoV-19/South_Africa/NHLS-UCT-GS-B478/2021\|EPI_ISL_2140702\|2021-03-31 | 20H (Beta, V2) |
| hCoV-19/South_Africa/NHLS-UCT-GS-B479/2021\|EPI_ISL_2140703\|2021-04-01 | 20H (Beta, V2) |
| hCoV-19/South_Africa/NHLS-UCT-GS-B481/2021\|EPI_ISL_2140704\|2021-04-02 | 20H (Beta, V2) |
| hCoV-19/South_Africa/NHLS-UCT-GS-B484/2021\|EPI_ISL_2140705\|2021-03-27 | 20H (Beta, V2) |
| hCoV-19/South_Africa/NHLS-UCT-GS-B487/2021\|EPI_ISL_2140706\|2021-03-29 | 20H (Beta, V2) |
| hCoV-19/South_Africa/NHLS-UCT-GS-B488/2021\|EPI_ISL_2140707\|2021-03-29 | 20H (Beta, V2) |
| hCoV-19/South_Africa/NHLS-UCT-GS-B501/2021\|EPI_ISL_2140708\|2021-04-01 | 20H (Beta, V2) |
| hCoV-19/South_Africa/NHLS-UCT-GS-B505/2021\|EPI_ISL_2140709\|2021-04-01 | 20H (Beta, V2) |
| hCoV-19/South_Africa/NHLS-UCT-GS-B510/2021\|EPI_ISL_2140710\|2021-03-29 | 20H (Beta, V2) |
| hCoV-19/South_Africa/NHLS-UCT-GS-B515/2021\|EPI_ISL_2140711\|2021-03-31 | 20H (Beta, V2) |
| hCoV-19/South_Africa/NHLS-UCT-GS-B530/2021\|EPI_ISL_2140712\|2021-04-09 | 20H (Beta, V2) |
| hCoV-19/South_Africa/NHLS-UCT-GS-B531/2021\|EPI_ISL_2140713\|2021-04-04 | 20H (Beta, V2) |
| hCoV-19/South_Africa/NHLS-UCT-GS-B535/2021\|EPI_ISL_2140714\|2021-04-05 | 20H (Beta, V2) |
| hCoV-19/South_Africa/NHLS-UCT-GS-B537/2021\|EPI_ISL_2140715\|2021-04-06 | 20H (Beta, V2) |
| hCoV-19/South_Africa/NHLS-UCT-GS-B540/2021\|EPI_ISL_2140716\|2021-04-06 | 20H (Beta, V2) |
| hCoV-19/South_Africa/NHLS-UCT-GS-B542/2021\|EPI_ISL_2140717\|2021-04-07 | 20H (Beta, V2) |
| hCoV-19/South_Africa/NHLS-UCT-GS-B543/2021\|EPI_ISL_2140718\|2021-04-07 | 20H (Beta, V2) |
| hCoV-19/South_Africa/NHLS-UCT-GS-B544/2021\|EPI_ISL_2140719\|2021-04-08 | 20H (Beta, V2) |
| hCoV-19/South_Africa/NHLS-UCT-GS-B547/2021\|EPI_ISL_2140720\|2021-04-09 | 20H (Beta, V2) |
| hCoV-19/South_Africa/NHLS-UCT-GS-B551/2021\|EPI_ISL_2140721\|2021-04-09 | 20H (Beta, V2) |
| hCoV-19/South_Africa/NHLS-UCT-GS-B555/2021\|EPI_ISL_2140722\|2021-04-06 | 20H (Beta, V2) |
| hCoV-19/South_Africa/NHLS-UCT-GS-B556/2021\|EPI_ISL_2140723\|2021-04-05 | 20H (Beta, V2) |
| hCoV-19/South_Africa/NHLS-UCT-GS-B557/2021\|EPI_ISL_2140724\|2021-04-06 | 20H (Beta, V2) |

| | |
|---|---|
| hCoV-19/South_Africa/NHLS-UCT-GS-B559/2021\|EPI_ISL_2140725\|2021-04-07 | 20H (Beta, V2) |
| hCoV-19/South_Africa/NHLS-UCT-GS-B561/2021\|EPI_ISL_2140726\|2021-04-07 | 20H (Beta, V2) |
| hCoV-19/South_Africa/NHLS-UCT-GS-B563/2021\|EPI_ISL_2140727\|2021-04-08 | 20H (Beta, V2) |
| hCoV-19/South_Africa/NHLS-UCT-GS-B564/2021\|EPI_ISL_2140728\|2021-04-08 | 20H (Beta, V2) |
| hCoV-19/South_Africa/NHLS-UCT-GS-B580/2021\|EPI_ISL_3506436\|2021-04-19 | 20H (Beta, V2) |
| hCoV-19/South_Africa/NHLS-UCT-GS-B594/2021\|EPI_ISL_3506363\|2021-04-19 | 20H (Beta, V2) |
| hCoV-19/South_Africa/NHLS-UCT-GS-B602/2021\|EPI_ISL_2375989\|2021-04-20 | 20H (Beta, V2) |
| hCoV-19/South_Africa/NHLS-UCT-GS-B617/2021\|EPI_ISL_2375990\|2021-04-23 | 20H (Beta, V2) |
| hCoV-19/South_Africa/NHLS-UCT-GS-B665/2021\|EPI_ISL_2375991\|2021-04-28 | 20H (Beta, V2) |
| hCoV-19/South_Africa/NHLS-UCT-GS-B691/2021\|EPI_ISL_2375992\|2021-04-30 | 20H (Beta, V2) |
| hCoV-19/South_Africa/NHLS-UCT-GS-B695/2021\|EPI_ISL_2375993\|2021-05-03 | 20H (Beta, V2) |
| hCoV-19/South_Africa/NHLS-UCT-GS-B708/2021\|EPI_ISL_2375994\|2021-05-04 | 20H (Beta, V2) |
| hCoV-19/South_Africa/NHLS-UCT-GS-B717/2021\|EPI_ISL_2375995\|2021-05-05 | 20H (Beta, V2) |
| hCoV-19/South_Africa/NHLS-UCT-GS-B741/2021\|EPI_ISL_3506437\|2021-05-05 | 20H (Beta, V2) |
| hCoV-19/South_Africa/NHLS-UCT-GS-B766/2021\|EPI_ISL_2547409\|2021-05-07 | 21D (Eta) |
| hCoV-19/South_Africa/NHLS-UCT-GS-B781/2021\|EPI_ISL_2547410\|2021-05-10 | 21D (Eta) |
| hCoV-19/South_Africa/NHLS-UCT-GS-B787/2021\|EPI_ISL_2547411\|2021-05-12 | 20H (Beta, V2) |
| hCoV-19/South_Africa/NHLS-UCT-GS-B793/2021\|EPI_ISL_2547412\|2021-05-13 | 20I (Alpha, V1) |
| hCoV-19/South_Africa/NHLS-UCT-GS-B799/2021\|EPI_ISL_2547413\|2021-05-14 | 20H (Beta, V2) |
| hCoV-19/South_Africa/NHLS-UCT-GS-B801/2021\|EPI_ISL_2547414\|2021-05-08 | 20B |
| hCoV-19/South_Africa/NHLS-UCT-GS-B806/2021\|EPI_ISL_2547415\|2021-05-12 | 20H (Beta, V2) |
| hCoV-19/South_Africa/NHLS-UCT-GS-B810/2021\|EPI_ISL_3506435\|2021-05-12 | 20H (Beta, V2) |
| hCoV-19/South_Africa/NHLS-UCT-GS-B812/2021\|EPI_ISL_2547416\|2021-05-14 | 21D (Eta) |
| hCoV-19/South_Africa/NHLS-UCT-GS-B817/2021\|EPI_ISL_2547417\|2021-05-15 | 20I (Alpha, V1) |
| hCoV-19/South_Africa/NHLS-UCT-GS-B822/2021\|EPI_ISL_2547418\|2021-05-16 | 20H (Beta, V2) |
| hCoV-19/South_Africa/NHLS-UCT-GS-B823/2021\|EPI_ISL_2547419\|2021-05-17 | 20H (Beta, V2) |
| hCoV-19/South_Africa/NHLS-UCT-GS-B832/2021\|EPI_ISL_2547420\|2021-05-17 | 20H (Beta, V2) |
| hCoV-19/South_Africa/NHLS-UCT-GS-B837/2021\|EPI_ISL_2547421\|2021-05-17 | 20H (Beta, V2) |
| hCoV-19/South_Africa/NHLS-UCT-GS-B838/2021\|EPI_ISL_2547422\|2021-05-17 | 20H (Beta, V2) |
| hCoV-19/South_Africa/NHLS-UCT-GS-B854/2021\|EPI_ISL_2547423\|2021-05-18 | 20I (Alpha, V1) |
| hCoV-19/South_Africa/NHLS-UCT-GS-B865/2021\|EPI_ISL_2547424\|2021-05-20 | 20H (Beta, V2) |
| hCoV-19/South_Africa/NHLS-UCT-GS-B866/2021\|EPI_ISL_2547425\|2021-05-20 | 20H (Beta, V2) |
| hCoV-19/South_Africa/NHLS-UCT-GS-B872/2021\|EPI_ISL_2547426\|2021-05-20 | 20H (Beta, V2) |
| hCoV-19/South_Africa/NHLS-UCT-GS-B877/2021\|EPI_ISL_2547427\|2021-05-21 | 20H (Beta, V2) |
| hCoV-19/South_Africa/NHLS-UCT-GS-B880/2021\|EPI_ISL_2621054\|2021-05-22 | 20H (Beta, V2) |
| hCoV-19/South_Africa/NHLS-UCT-GS-B888/2021\|EPI_ISL_2621059\|2021-05-25 | 20B |
| hCoV-19/South_Africa/NHLS-UCT-GS-B895/2021\|EPI_ISL_2621132\|2021-05-27 | 20H (Beta, V2) |
| hCoV-19/South_Africa/NHLS-UCT-GS-B898/2021\|EPI_ISL_2621133\|2021-05-28 | 20H (Beta, V2) |
| hCoV-19/South_Africa/NHLS-UCT-GS-B909/2021\|EPI_ISL_2621134\|2021-05-24 | 20H (Beta, V2) |
| hCoV-19/South_Africa/NHLS-UCT-GS-B912/2021\|EPI_ISL_2621135\|2021-05-24 | 21A (Delta) |
| hCoV-19/South_Africa/NHLS-UCT-GS-B918/2021\|EPI_ISL_2621136\|2021-05-25 | 20H (Beta, V2) |
| hCoV-19/South_Africa/NHLS-UCT-GS-B928/2021\|EPI_ISL_2621060\|2021-05-25 | 20H (Beta, V2) |
| hCoV-19/South_Africa/NHLS-UCT-GS-B933/2021\|EPI_ISL_2621137\|2021-05-26 | 20H (Beta, V2) |
| hCoV-19/South_Africa/NHLS-UCT-GS-B937/2021\|EPI_ISL_2621138\|2021-05-26 | 21A (Delta) |
| hCoV-19/South_Africa/NHLS-UCT-GS-B938/2021\|EPI_ISL_2621139\|2021-05-26 | 21A (Delta) |
| hCoV-19/South_Africa/NHLS-UCT-GS-B940/2021\|EPI_ISL_2621140\|2021-05-26 | 21D (Eta) |
| hCoV-19/South_Africa/NHLS-UCT-GS-B944/2021\|EPI_ISL_2621141\|2021-05-26 | 21D (Eta) |
| hCoV-19/South_Africa/NHLS-UCT-GS-B951/2021\|EPI_ISL_2621142\|2021-05-26 | 21D (Eta) |
| hCoV-19/South_Africa/NHLS-UCT-GS-B952/2021\|EPI_ISL_2621061\|2021-05-27 | 20H (Beta, V2) |
| hCoV-19/South_Africa/NHLS-UCT-GS-B953/2021\|EPI_ISL_2621143\|2021-05-27 | 20H (Beta, V2) |

| | |
|---|---|
| hCoV-19/South_Africa/NHLS-UCT-GS-B964/2021\|EPI_ISL_3506438\|2021-05-28 | 20H (Beta, V2) |
| hCoV-19/South_Africa/NHLS-UCT-GS-B966/2021\|EPI_ISL_2621144\|2021-05-28 | 20H (Beta, V2) |
| hCoV-19/South_Africa/NHLS-UCT-GS-B967/2021\|EPI_ISL_3506396\|2021-05-28 | 20I (Alpha, V1) |
| hCoV-19/South_Africa/NHLS-UCT-GS-B982/2021\|EPI_ISL_2621145\|2021-05-27 | 20H (Beta, V2) |
| hCoV-19/South_Africa/NHLS-UCT-GS-B989/2021\|EPI_ISL_2621146\|2021-05-27 | 20H (Beta, V2) |
| hCoV-19/South_Africa/NHLS-UCT-GS-B990/2021\|EPI_ISL_2621147\|2021-05-28 | 20H (Beta, V2) |
| hCoV-19/South_Africa/NHLS-UCT-GS-B992/2021\|EPI_ISL_2802199\|2021-05-29 | 21A (Delta) |
| hCoV-19/South_Africa/NHLS-UCT-GS-B998/2021\|EPI_ISL_2802178\|2021-05-31 | 20H (Beta, V2) |
| hCoV-19/South_Africa/NHLS-UCT-GS-C004/2021\|EPI_ISL_2802179\|2021-05-31 | 20H (Beta, V2) |
| hCoV-19/South_Africa/NHLS-UCT-GS-C018/2021\|EPI_ISL_2802180\|2021-05-31 | 21A (Delta) |
| hCoV-19/South_Africa/NHLS-UCT-GS-C025/2021\|EPI_ISL_2802181\|2021-05-31 | 21A (Delta) |
| hCoV-19/South_Africa/NHLS-UCT-GS-C027/2021\|EPI_ISL_2802192\|2021-06-01 | 20H (Beta, V2) |
| hCoV-19/South_Africa/NHLS-UCT-GS-C029/2021\|EPI_ISL_2802193\|2021-06-01 | 20H (Beta, V2) |
| hCoV-19/South_Africa/NHLS-UCT-GS-C031/2021\|EPI_ISL_2802194\|2021-06-01 | 21A (Delta) |
| hCoV-19/South_Africa/NHLS-UCT-GS-C033/2021\|EPI_ISL_2802182\|2021-06-01 | 20H (Beta, V2) |
| hCoV-19/South_Africa/NHLS-UCT-GS-C037/2021\|EPI_ISL_3957792\|2021-06-02 | 20H (Beta, V2) |
| hCoV-19/South_Africa/NHLS-UCT-GS-C045/2021\|EPI_ISL_3506360\|2021-06-02 | 21A (Delta) |
| hCoV-19/South_Africa/NHLS-UCT-GS-C080/2021\|EPI_ISL_3957793\|2021-06-04 | 20H (Beta, V2) |
| hCoV-19/South_Africa/NHLS-UCT-GS-C087/2021\|EPI_ISL_3957775\|2021-06-02 | 20H (Beta, V2) |
| hCoV-19/South_Africa/NHLS-UCT-GS-C101/2021\|EPI_ISL_2802195\|2021-06-07 | 21A (Delta) |
| hCoV-19/South_Africa/NHLS-UCT-GS-C102/2021\|EPI_ISL_2876940\|2021-06-11 | 21A (Delta) |
| hCoV-19/South_Africa/NHLS-UCT-GS-C104/2021\|EPI_ISL_2876941\|2021-06-11 | 21A (Delta) |
| hCoV-19/South_Africa/NHLS-UCT-GS-C105/2021\|EPI_ISL_2876942\|2021-06-11 | 21A (Delta) |
| hCoV-19/South_Africa/NHLS-UCT-GS-C108/2021\|EPI_ISL_2876943\|2021-06-14 | 20A |
| hCoV-19/South_Africa/NHLS-UCT-GS-C109/2021\|EPI_ISL_2876944\|2021-06-14 | 21A (Delta) |
| hCoV-19/South_Africa/NHLS-UCT-GS-C110/2021\|EPI_ISL_2876945\|2021-06-14 | 21A (Delta) |
| hCoV-19/South_Africa/NHLS-UCT-GS-C111/2021\|EPI_ISL_2876946\|2021-06-14 | 21A (Delta) |
| hCoV-19/South_Africa/NHLS-UCT-GS-C112/2021\|EPI_ISL_2876947\|2021-06-14 | 20A |
| hCoV-19/South_Africa/NHLS-UCT-GS-C113/2021\|EPI_ISL_2876948\|2021-06-14 | 21A (Delta) |
| hCoV-19/South_Africa/NHLS-UCT-GS-C114/2021\|EPI_ISL_2876935\|2021-06-14 | 20H (Beta, V2) |
| hCoV-19/South_Africa/NHLS-UCT-GS-C115/2021\|EPI_ISL_2876949\|2021-06-14 | 21A (Delta) |
| hCoV-19/South_Africa/NHLS-UCT-GS-C116/2021\|EPI_ISL_2876950\|2021-06-14 | 21A (Delta) |
| hCoV-19/South_Africa/NHLS-UCT-GS-C117/2021\|EPI_ISL_2876951\|2021-06-14 | 20I (Alpha, V1) |
| hCoV-19/South_Africa/NHLS-UCT-GS-C118/2021\|EPI_ISL_2876952\|2021-06-14 | 21A (Delta) |
| hCoV-19/South_Africa/NHLS-UCT-GS-C119/2021\|EPI_ISL_2876953\|2021-06-15 | 21A (Delta) |
| hCoV-19/South_Africa/NHLS-UCT-GS-C120/2021\|EPI_ISL_2876954\|2021-06-14 | 21A (Delta) |
| hCoV-19/South_Africa/NHLS-UCT-GS-C121/2021\|EPI_ISL_2876955\|2021-06-15 | 21A (Delta) |
| hCoV-19/South_Africa/NHLS-UCT-GS-C122/2021\|EPI_ISL_2876936\|2021-06-15 | 20H (Beta, V2) |
| hCoV-19/South_Africa/NHLS-UCT-GS-C123/2021\|EPI_ISL_2876956\|2021-06-14 | 20I (Alpha, V1) |
| hCoV-19/South_Africa/NHLS-UCT-GS-C124/2021\|EPI_ISL_2876957\|2021-06-15 | 21A (Delta) |
| hCoV-19/South_Africa/NHLS-UCT-GS-C125/2021\|EPI_ISL_2876958\|2021-06-15 | 20H (Beta, V2) |
| hCoV-19/South_Africa/NHLS-UCT-GS-C126/2021\|EPI_ISL_2876959\|2021-06-15 | 21A (Delta) |
| hCoV-19/South_Africa/NHLS-UCT-GS-C127/2021\|EPI_ISL_2876960\|2021-06-15 | 21A (Delta) |
| hCoV-19/South_Africa/NHLS-UCT-GS-C128/2021\|EPI_ISL_2876961\|2021-06-15 | 21A (Delta) |
| hCoV-19/South_Africa/NHLS-UCT-GS-C129/2021\|EPI_ISL_2876962\|2021-06-15 | 21A (Delta) |
| hCoV-19/South_Africa/NHLS-UCT-GS-C130/2021\|EPI_ISL_2876963\|2021-06-15 | 21A (Delta) |
| hCoV-19/South_Africa/NHLS-UCT-GS-C131/2021\|EPI_ISL_2876964\|2021-06-15 | 21A (Delta) |
| hCoV-19/South_Africa/NHLS-UCT-GS-C132/2021\|EPI_ISL_2876965\|2021-06-15 | 21A (Delta) |
| hCoV-19/South_Africa/NHLS-UCT-GS-C133/2021\|EPI_ISL_2876966\|2021-06-15 | 21A (Delta) |
| hCoV-19/South_Africa/NHLS-UCT-GS-C134/2021\|EPI_ISL_2876967\|2021-06-15 | 21A (Delta) |

| | |
|---|---|
| hCoV-19/South_Africa/NHLS-UCT-GS-C135/2021\|EPI_ISL_2876968\|2021-06-15 | 21A (Delta) |
| hCoV-19/South_Africa/NHLS-UCT-GS-C136/2021\|EPI_ISL_2876969\|2021-06-15 | 21A (Delta) |
| hCoV-19/South_Africa/NHLS-UCT-GS-C137/2021\|EPI_ISL_2876970\|2021-06-16 | 21A (Delta) |
| hCoV-19/South_Africa/NHLS-UCT-GS-C138/2021\|EPI_ISL_2876971\|2021-06-17 | 21A (Delta) |
| hCoV-19/South_Africa/NHLS-UCT-GS-C139/2021\|EPI_ISL_2876972\|2021-06-17 | 20H (Beta, V2) |
| hCoV-19/South_Africa/NHLS-UCT-GS-C140/2021\|EPI_ISL_2876973\|2021-06-17 | 21A (Delta) |
| hCoV-19/South_Africa/NHLS-UCT-GS-C141/2021\|EPI_ISL_2876974\|2021-06-17 | 20B |
| hCoV-19/South_Africa/NHLS-UCT-GS-C143/2021\|EPI_ISL_2876975\|2021-06-17 | 21A (Delta) |
| hCoV-19/South_Africa/NHLS-UCT-GS-C145/2021\|EPI_ISL_2876976\|2021-06-17 | 21A (Delta) |
| hCoV-19/South_Africa/NHLS-UCT-GS-C146/2021\|EPI_ISL_2876977\|2021-06-17 | 21A (Delta) |
| hCoV-19/South_Africa/NHLS-UCT-GS-C148/2021\|EPI_ISL_2876978\|2021-06-17 | 20H (Beta, V2) |
| hCoV-19/South_Africa/NHLS-UCT-GS-C149/2021\|EPI_ISL_2876979\|2021-06-17 | 21A (Delta) |
| hCoV-19/South_Africa/NHLS-UCT-GS-C150/2021\|EPI_ISL_2876980\|2021-06-17 | 21A (Delta) |
| hCoV-19/South_Africa/NHLS-UCT-GS-C151/2021\|EPI_ISL_2876981\|2021-06-17 | 21A (Delta) |
| hCoV-19/South_Africa/NHLS-UCT-GS-C152/2021\|EPI_ISL_2876982\|2021-06-17 | 20H (Beta, V2) |
| hCoV-19/South_Africa/NHLS-UCT-GS-C155/2021\|EPI_ISL_2876983\|2021-06-17 | 21A (Delta) |
| hCoV-19/South_Africa/NHLS-UCT-GS-C156/2021\|EPI_ISL_2876984\|2021-06-17 | 21A (Delta) |
| hCoV-19/South_Africa/NHLS-UCT-GS-C157/2021\|EPI_ISL_2876985\|2021-06-17 | 21A (Delta) |
| hCoV-19/South_Africa/NHLS-UCT-GS-C158/2021\|EPI_ISL_2876986\|2021-06-17 | 21A (Delta) |
| hCoV-19/South_Africa/NHLS-UCT-GS-C160/2021\|EPI_ISL_2876987\|2021-06-17 | 21A (Delta) |
| hCoV-19/South_Africa/NHLS-UCT-GS-C161/2021\|EPI_ISL_2876988\|2021-06-17 | 21A (Delta) |
| hCoV-19/South_Africa/NHLS-UCT-GS-C162/2021\|EPI_ISL_2876989\|2021-06-18 | 20H (Beta, V2) |
| hCoV-19/South_Africa/NHLS-UCT-GS-C163/2021\|EPI_ISL_2876990\|2021-06-17 | 21A (Delta) |
| hCoV-19/South_Africa/NHLS-UCT-GS-C165/2021\|EPI_ISL_2876991\|2021-06-18 | 21A (Delta) |
| hCoV-19/South_Africa/NHLS-UCT-GS-C166/2021\|EPI_ISL_2876992\|2021-06-17 | 21A (Delta) |
| hCoV-19/South_Africa/NHLS-UCT-GS-C167/2021\|EPI_ISL_2876993\|2021-06-18 | 20A |
| hCoV-19/South_Africa/NHLS-UCT-GS-C168/2021\|EPI_ISL_2876994\|2021-06-17 | 21A (Delta) |
| hCoV-19/South_Africa/NHLS-UCT-GS-C169/2021\|EPI_ISL_2876995\|2021-06-17 | 21A (Delta) |
| hCoV-19/South_Africa/NHLS-UCT-GS-C170/2021\|EPI_ISL_2876996\|2021-06-17 | 21A (Delta) |
| hCoV-19/South_Africa/NHLS-UCT-GS-C171/2021\|EPI_ISL_2876997\|2021-06-18 | 21A (Delta) |
| hCoV-19/South_Africa/NHLS-UCT-GS-C172/2021\|EPI_ISL_2876998\|2021-06-18 | 21A (Delta) |
| hCoV-19/South_Africa/NHLS-UCT-GS-C173/2021\|EPI_ISL_2876999\|2021-06-12 | 21A (Delta) |
| hCoV-19/South_Africa/NHLS-UCT-GS-C174/2021\|EPI_ISL_2877000\|2021-06-15 | 21A (Delta) |
| hCoV-19/South_Africa/NHLS-UCT-GS-C176/2021\|EPI_ISL_2876937\|2021-06-17 | 21A (Delta) |
| hCoV-19/South_Africa/NHLS-UCT-GS-C177/2021\|EPI_ISL_2877001\|2021-06-17 | 21A (Delta) |
| hCoV-19/South_Africa/NHLS-UCT-GS-C178/2021\|EPI_ISL_2877002\|2021-06-14 | 21A (Delta) |
| hCoV-19/South_Africa/NHLS-UCT-GS-C180/2021\|EPI_ISL_2877003\|2021-06-14 | 21A (Delta) |
| hCoV-19/South_Africa/NHLS-UCT-GS-C181/2021\|EPI_ISL_2877004\|2021-06-17 | 21A (Delta) |
| hCoV-19/South_Africa/NHLS-UCT-GS-C182/2021\|EPI_ISL_2877005\|2021-06-18 | 21A (Delta) |
| hCoV-19/South_Africa/NHLS-UCT-GS-C183/2021\|EPI_ISL_2877006\|2021-06-18 | 21A (Delta) |
| hCoV-19/South_Africa/NHLS-UCT-GS-C187/2021\|EPI_ISL_3957794\|2021-06-06 | 21A (Delta) |
| hCoV-19/South_Africa/NHLS-UCT-GS-C191/2021\|EPI_ISL_3957795\|2021-06-07 | 21A (Delta) |
| hCoV-19/South_Africa/NHLS-UCT-GS-C192/2021\|EPI_ISL_3957796\|2021-06-08 | 20H (Beta, V2) |
| hCoV-19/South_Africa/NHLS-UCT-GS-C193/2021\|EPI_ISL_3957776\|2021-06-08 | 20B |
| hCoV-19/South_Africa/NHLS-UCT-GS-C198/2021\|EPI_ISL_3957797\|2021-06-08 | 21A (Delta) |
| hCoV-19/South_Africa/NHLS-UCT-GS-C210/2021\|EPI_ISL_3957777\|2021-06-09 | 20H (Beta, V2) |
| hCoV-19/South_Africa/NHLS-UCT-GS-C216/2021\|EPI_ISL_3957798\|2021-06-10 | 21A (Delta) |
| hCoV-19/South_Africa/NHLS-UCT-GS-C217/2021\|EPI_ISL_3957799\|2021-06-10 | 21A (Delta) |
| hCoV-19/South_Africa/NHLS-UCT-GS-C224/2021\|EPI_ISL_3957778\|2021-06-11 | 20B |
| hCoV-19/South_Africa/NHLS-UCT-GS-C233/2021\|EPI_ISL_3957800\|2021-06-05 | 20H (Beta, V2) |

| | |
|---|---|
| hCoV-19/South_Africa/NHLS-UCT-GS-C237/2021\|EPI_ISL_3957779\|2021-06-06 | 20H (Beta, V2) |
| hCoV-19/South_Africa/NHLS-UCT-GS-C242/2021\|EPI_ISL_3957780\|2021-06-07 | 20H (Beta, V2) |
| hCoV-19/South_Africa/NHLS-UCT-GS-C292/2021\|EPI_ISL_3957801\|2021-06-08 | 21A (Delta) |
| hCoV-19/South_Africa/NHLS-UCT-GS-C377/2021\|EPI_ISL_3957802\|2021-06-06 | 21A (Delta) |
| hCoV-19/South_Africa/NHLS-UCT-GS-C382/2021\|EPI_ISL_3957803\|2021-06-08 | 20H (Beta, V2) |
| hCoV-19/South_Africa/NHLS-UCT-GS-C383/2021\|EPI_ISL_3957781\|2021-06-08 | 20H (Beta, V2) |
| hCoV-19/South_Africa/NHLS-UCT-GS-C384/2021\|EPI_ISL_3957804\|2021-06-08 | 20H (Beta, V2) |
| hCoV-19/South_Africa/NHLS-UCT-GS-C385/2021\|EPI_ISL_3957805\|2021-06-09 | 20H (Beta, V2) |
| hCoV-19/South_Africa/NHLS-UCT-GS-C388/2021\|EPI_ISL_3957782\|2021-06-09 | 20B |
| hCoV-19/South_Africa/NHLS-UCT-GS-C394/2021\|EPI_ISL_3957806\|2021-06-11 | 20H (Beta, V2) |
| hCoV-19/South_Africa/NHLS-UCT-GS-C418/2021\|EPI_ISL_3957807\|2021-06-12 | 21A (Delta) |
| hCoV-19/South_Africa/NHLS-UCT-GS-C419/2021\|EPI_ISL_3957808\|2021-06-12 | 20H (Beta, V2) |
| hCoV-19/South_Africa/NHLS-UCT-GS-C420/2021\|EPI_ISL_3690553\|2021-06-13 | 20H (Beta, V2) |
| hCoV-19/South_Africa/NHLS-UCT-GS-C421/2021\|EPI_ISL_3690638\|2021-06-13 | 21A (Delta) |
| hCoV-19/South_Africa/NHLS-UCT-GS-C423/2021\|EPI_ISL_3957809\|2021-06-13 | 21A (Delta) |
| hCoV-19/South_Africa/NHLS-UCT-GS-C426/2021\|EPI_ISL_3690637\|2021-06-14 | 21A (Delta) |
| hCoV-19/South_Africa/NHLS-UCT-GS-C439/2021\|EPI_ISL_3690576\|2021-06-15 | 20H (Beta, V2) |
| hCoV-19/South_Africa/NHLS-UCT-GS-C440/2021\|EPI_ISL_3690634\|2021-06-15 | 21A (Delta) |
| hCoV-19/South_Africa/NHLS-UCT-GS-C446/2021\|EPI_ISL_3957783\|2021-06-15 | 20B |
| hCoV-19/South_Africa/NHLS-UCT-GS-C482/2021\|EPI_ISL_3506496\|2021-06-17 | 21A (Delta) |
| hCoV-19/South_Africa/NHLS-UCT-GS-C483/2021\|EPI_ISL_3506372\|2021-06-17 | 20H (Beta, V2) |
| hCoV-19/South_Africa/NHLS-UCT-GS-C491/2021\|EPI_ISL_3690556\|2021-06-17 | 21A (Delta) |
| hCoV-19/South_Africa/NHLS-UCT-GS-C499/2021\|EPI_ISL_3506467\|2021-06-18 | 21A (Delta) |
| hCoV-19/South_Africa/NHLS-UCT-GS-C501/2021\|EPI_ISL_3506497\|2021-06-18 | 21A (Delta) |
| hCoV-19/South_Africa/NHLS-UCT-GS-C505/2021\|EPI_ISL_3957810\|2021-06-11 | 21A (Delta) |
| hCoV-19/South_Africa/NHLS-UCT-GS-C521/2021\|EPI_ISL_3957784\|2021-06-11 | 20B |
| hCoV-19/South_Africa/NHLS-UCT-GS-C625/2021\|EPI_ISL_3690633\|2021-06-17 | 21A (Delta) |
| hCoV-19/South_Africa/NHLS-UCT-GS-C641/2021\|EPI_ISL_3957811\|2021-06-12 | 20H (Beta, V2) |
| hCoV-19/South_Africa/NHLS-UCT-GS-C646/2021\|EPI_ISL_3690577\|2021-06-14 | 20H (Beta, V2) |
| hCoV-19/South_Africa/NHLS-UCT-GS-C647/2021\|EPI_ISL_3690574\|2021-06-14 | 21A (Delta) |
| hCoV-19/South_Africa/NHLS-UCT-GS-C648/2021\|EPI_ISL_3690635\|2021-06-15 | 21A (Delta) |
| hCoV-19/South_Africa/NHLS-UCT-GS-C650/2021\|EPI_ISL_3690636\|2021-06-15 | 21A (Delta) |
| hCoV-19/South_Africa/NHLS-UCT-GS-C652/2021\|EPI_ISL_3690552\|2021-06-17 | 20H (Beta, V2) |
| hCoV-19/South_Africa/NHLS-UCT-GS-C655/2021\|EPI_ISL_3506428\|2021-06-18 | 20H (Beta, V2) |
| hCoV-19/South_Africa/NHLS-UCT-GS-C681/2021\|EPI_ISL_3506369\|2021-06-19 | 21A (Delta) |
| hCoV-19/South_Africa/NHLS-UCT-GS-C682/2021\|EPI_ISL_3506482\|2021-06-19 | 21A (Delta) |
| hCoV-19/South_Africa/NHLS-UCT-GS-C686/2021\|EPI_ISL_3506389\|2021-06-19 | 21A (Delta) |
| hCoV-19/South_Africa/NHLS-UCT-GS-C687/2021\|EPI_ISL_3506422\|2021-06-19 | 21A (Delta) |
| hCoV-19/South_Africa/NHLS-UCT-GS-C689/2021\|EPI_ISL_3506498\|2021-06-19 | 21A (Delta) |
| hCoV-19/South_Africa/NHLS-UCT-GS-C693/2021\|EPI_ISL_3506370\|2021-06-20 | 21A (Delta) |
| hCoV-19/South_Africa/NHLS-UCT-GS-C694/2021\|EPI_ISL_3506499\|2021-06-20 | 21A (Delta) |
| hCoV-19/South_Africa/NHLS-UCT-GS-C707/2021\|EPI_ISL_3506431\|2021-06-21 | 20H (Beta, V2) |
| hCoV-19/South_Africa/NHLS-UCT-GS-C709/2021\|EPI_ISL_3506500\|2021-06-21 | 21A (Delta) |
| hCoV-19/South_Africa/NHLS-UCT-GS-C710/2021\|EPI_ISL_3506432\|2021-06-21 | 20H (Beta, V2) |
| hCoV-19/South_Africa/NHLS-UCT-GS-C713/2021\|EPI_ISL_3506394\|2021-06-21 | 20I (Alpha, V1) |
| hCoV-19/South_Africa/NHLS-UCT-GS-C714/2021\|EPI_ISL_3506430\|2021-06-21 | 20H (Beta, V2) |
| hCoV-19/South_Africa/NHLS-UCT-GS-C732/2021\|EPI_ISL_3506483\|2021-06-22 | 21A (Delta) |
| hCoV-19/South_Africa/NHLS-UCT-GS-C736/2021\|EPI_ISL_3506443\|2021-06-23 | 21A (Delta) |
| hCoV-19/South_Africa/NHLS-UCT-GS-C737/2021\|EPI_ISL_3506421\|2021-06-23 | 21A (Delta) |
| hCoV-19/South_Africa/NHLS-UCT-GS-C739/2021\|EPI_ISL_3506475\|2021-06-23 | 21A (Delta) |

| | |
|---|---|
| hCoV-19/South_Africa/NHLS-UCT-GS-C740/2021\|EPI_ISL_3957812\|2021-06-23 | 21A (Delta) |
| hCoV-19/South_Africa/NHLS-UCT-GS-C745/2021\|EPI_ISL_3506468\|2021-06-24 | 21A (Delta) |
| hCoV-19/South_Africa/NHLS-UCT-GS-C750/2021\|EPI_ISL_3506501\|2021-06-24 | 21A (Delta) |
| hCoV-19/South_Africa/NHLS-UCT-GS-C751/2021\|EPI_ISL_3506402\|2021-06-24 | 21A (Delta) |
| hCoV-19/South_Africa/NHLS-UCT-GS-C753/2021\|EPI_ISL_3506391\|2021-06-24 | 20I (Alpha, V1) |
| hCoV-19/South_Africa/NHLS-UCT-GS-C754/2021\|EPI_ISL_3506376\|2021-06-25 | 21A (Delta) |
| hCoV-19/South_Africa/NHLS-UCT-GS-C755/2021\|EPI_ISL_3506444\|2021-06-25 | 21A (Delta) |
| hCoV-19/South_Africa/NHLS-UCT-GS-C756/2021\|EPI_ISL_3506445\|2021-06-25 | 21A (Delta) |
| hCoV-19/South_Africa/NHLS-UCT-GS-C758/2021\|EPI_ISL_3506476\|2021-06-25 | 21A (Delta) |
| hCoV-19/South_Africa/NHLS-UCT-GS-C760/2021\|EPI_ISL_3506426\|2021-06-25 | 20H (Beta, V2) |
| hCoV-19/South_Africa/NHLS-UCT-GS-C765/2021\|EPI_ISL_3506400\|2021-06-25 | 21A (Delta) |
| hCoV-19/South_Africa/NHLS-UCT-GS-C771/2021\|EPI_ISL_3506469\|2021-06-18 | 21A (Delta) |
| hCoV-19/South_Africa/NHLS-UCT-GS-C775/2021\|EPI_ISL_3506384\|2021-06-21 | 21A (Delta) |
| hCoV-19/South_Africa/NHLS-UCT-GS-C776/2021\|EPI_ISL_3506382\|2021-06-18 | 21A (Delta) |
| hCoV-19/South_Africa/NHLS-UCT-GS-C780/2021\|EPI_ISL_3506446\|2021-06-24 | 21A (Delta) |
| hCoV-19/South_Africa/NHLS-UCT-GS-C782/2021\|EPI_ISL_3506361\|2021-06-24 | 21A (Delta) |
| hCoV-19/South_Africa/NHLS-UCT-GS-C790/2021\|EPI_ISL_3506447\|2021-06-24 | 21A (Delta) |
| hCoV-19/South_Africa/NHLS-UCT-GS-C791/2021\|EPI_ISL_3506401\|2021-06-22 | 21A (Delta) |
| hCoV-19/South_Africa/NHLS-UCT-GS-C803/2021\|EPI_ISL_3506427\|2021-06-24 | 20H (Beta, V2) |
| hCoV-19/South_Africa/NHLS-UCT-GS-C813/2021\|EPI_ISL_3506448\|2021-06-22 | 21A (Delta) |
| hCoV-19/South_Africa/NHLS-UCT-GS-C821/2021\|EPI_ISL_3506502\|2021-06-18 | 21A (Delta) |
| hCoV-19/South_Africa/NHLS-UCT-GS-C830/2021\|EPI_ISL_3506449\|2021-06-24 | 21A (Delta) |
| hCoV-19/South_Africa/NHLS-UCT-GS-C838/2021\|EPI_ISL_3506477\|2021-06-22 | 21A (Delta) |
| hCoV-19/South_Africa/NHLS-UCT-GS-C839/2021\|EPI_ISL_3506450\|2021-06-22 | 21A (Delta) |
| hCoV-19/South_Africa/NHLS-UCT-GS-C844/2021\|EPI_ISL_3506451\|2021-06-24 | 21A (Delta) |
| hCoV-19/South_Africa/NHLS-UCT-GS-C885/2021\|EPI_ISL_3506403\|2021-06-24 | 21A (Delta) |
| hCoV-19/South_Africa/NHLS-UCT-GS-C896/2021\|EPI_ISL_3506433\|2021-06-21 | 20H (Beta, V2) |
| hCoV-19/South_Africa/NHLS-UCT-GS-C897/2021\|EPI_ISL_3506478\|2021-06-21 | 21A (Delta) |
| hCoV-19/South_Africa/NHLS-UCT-GS-C908/2021\|EPI_ISL_3506452\|2021-06-22 | 21A (Delta) |
| hCoV-19/South_Africa/NHLS-UCT-GS-C909/2021\|EPI_ISL_3506453\|2021-06-22 | 21A (Delta) |
| hCoV-19/South_Africa/NHLS-UCT-GS-C911/2021\|EPI_ISL_3506454\|2021-06-23 | 21A (Delta) |
| hCoV-19/South_Africa/NHLS-UCT-GS-C915/2021\|EPI_ISL_3506455\|2021-06-22 | 21A (Delta) |
| hCoV-19/South_Africa/NHLS-UCT-GS-C916/2021\|EPI_ISL_3506404\|2021-06-23 | 21A (Delta) |
| hCoV-19/South_Africa/NHLS-UCT-GS-C919/2021\|EPI_ISL_3506456\|2021-06-23 | 21A (Delta) |
| hCoV-19/South_Africa/NHLS-UCT-GS-C920/2021\|EPI_ISL_3506457\|2021-06-22 | 21A (Delta) |
| hCoV-19/South_Africa/NHLS-UCT-GS-C921/2021\|EPI_ISL_3506390\|2021-06-24 | 21A (Delta) |
| hCoV-19/South_Africa/NHLS-UCT-GS-C922/2021\|EPI_ISL_3506458\|2021-06-24 | 21A (Delta) |
| hCoV-19/South_Africa/NHLS-UCT-GS-C929/2021\|EPI_ISL_3506459\|2021-06-25 | 21A (Delta) |
| hCoV-19/South_Africa/NHLS-UCT-GS-C935/2021\|EPI_ISL_3506460\|2021-06-22 | 21A (Delta) |
| hCoV-19/South_Africa/NHLS-UCT-GS-C943/2021\|EPI_ISL_3506386\|2021-06-25 | 21A (Delta) |
| hCoV-19/South_Africa/NHLS-UCT-GS-C948/2021\|EPI_ISL_3506358\|2021-06-24 | 21A (Delta) |
| hCoV-19/South_Africa/NHLS-UCT-GS-C950/2021\|EPI_ISL_3506503\|2021-06-22 | 21A (Delta) |
| hCoV-19/South_Africa/NHLS-UCT-GS-C952/2021\|EPI_ISL_3506461\|2021-06-22 | 21A (Delta) |
| hCoV-19/South_Africa/NHLS-UCT-GS-C955/2021\|EPI_ISL_3506378\|2021-06-18 | 20H (Beta, V2) |
| hCoV-19/South_Africa/NHLS-UCT-GS-C956/2021\|EPI_ISL_3506470\|2021-06-24 | 21A (Delta) |
| hCoV-19/South_Africa/NHLS-UCT-GS-C957/2021\|EPI_ISL_3506366\|2021-06-19 | 21A (Delta) |
| hCoV-19/South_Africa/NHLS-UCT-GS-C958/2021\|EPI_ISL_3506429\|2021-06-22 | 20H (Beta, V2) |
| hCoV-19/South_Africa/NHLS-UCT-GS-C959/2021\|EPI_ISL_3506491\|2021-06-19 | 21A (Delta) |
| hCoV-19/South_Africa/NHLS-UCT-GS-C963/2021\|EPI_ISL_3690632\|2021-06-22 | 21A (Delta) |
| hCoV-19/South_Africa/NHLS-UCT-GS-C967/2021\|EPI_ISL_3506359\|2021-06-22 | 21A (Delta) |

| | |
|---|---|
| hCoV-19/South_Africa/NHLS-UCT-GS-C982/2021\|EPI_ISL_3506374\|2021-06-24 | 21A (Delta) |
| hCoV-19/South_Africa/NHLS-UCT-GS-C984/2021\|EPI_ISL_3506395\|2021-06-23 | 20I (Alpha, V1) |
| hCoV-19/South_Africa/NHLS-UCT-GS-C987/2021\|EPI_ISL_3506405\|2021-06-24 | 21A (Delta) |
| hCoV-19/South_Africa/NHLS-UCT-GS-C998/2021\|EPI_ISL_3506423\|2021-06-24 | 20H (Beta, V2) |
| hCoV-19/South_Africa/NHLS-UCT-GS-D004/2021\|EPI_ISL_3506555\|2021-06-09 | 21A (Delta) |
| hCoV-19/South_Africa/NHLS-UCT-GS-D005/2021\|EPI_ISL_3506556\|2021-06-18 | 21A (Delta) |
| hCoV-19/South_Africa/NHLS-UCT-GS-D006/2021\|EPI_ISL_3506439\|2021-06-14 | 20H (Beta, V2) |
| hCoV-19/South_Africa/NHLS-UCT-GS-D008/2021\|EPI_ISL_3506419\|2021-06-15 | 21A (Delta) |
| hCoV-19/South_Africa/NHLS-UCT-GS-D009/2021\|EPI_ISL_3506557\|2021-06-22 | 21A (Delta) |
| hCoV-19/South_Africa/NHLS-UCT-GS-D010/2021\|EPI_ISL_3506420\|2021-06-22 | 21A (Delta) |
| hCoV-19/South_Africa/NHLS-UCT-GS-D011/2021\|EPI_ISL_3506558\|2021-06-21 | 21A (Delta) |
| hCoV-19/South_Africa/NHLS-UCT-GS-D013/2021\|EPI_ISL_3207537\|2021-06-28 | 21A (Delta) |
| hCoV-19/South_Africa/NHLS-UCT-GS-D014/2021\|EPI_ISL_3207538\|2021-06-29 | 21A (Delta) |
| hCoV-19/South_Africa/NHLS-UCT-GS-D015/2021\|EPI_ISL_3207539\|2021-06-30 | 21A (Delta) |
| hCoV-19/South_Africa/NHLS-UCT-GS-D016/2021\|EPI_ISL_3207503\|2021-06-30 | 21A (Delta) |
| hCoV-19/South_Africa/NHLS-UCT-GS-D018/2021\|EPI_ISL_3207540\|2021-07-01 | 21A (Delta) |
| hCoV-19/South_Africa/NHLS-UCT-GS-D019/2021\|EPI_ISL_3207541\|2021-07-01 | 21A (Delta) |
| hCoV-19/South_Africa/NHLS-UCT-GS-D020/2021\|EPI_ISL_3207542\|2021-07-01 | 21A (Delta) |
| hCoV-19/South_Africa/NHLS-UCT-GS-D021/2021\|EPI_ISL_3207543\|2021-07-01 | 21A (Delta) |
| hCoV-19/South_Africa/NHLS-UCT-GS-D022/2021\|EPI_ISL_3207544\|2021-07-01 | 21A (Delta) |
| hCoV-19/South_Africa/NHLS-UCT-GS-D023/2021\|EPI_ISL_3207545\|2021-07-01 | 21A (Delta) |
| hCoV-19/South_Africa/NHLS-UCT-GS-D024/2021\|EPI_ISL_3207546\|2021-07-01 | 21A (Delta) |
| hCoV-19/South_Africa/NHLS-UCT-GS-D025/2021\|EPI_ISL_3207547\|2021-07-02 | 21A (Delta) |
| hCoV-19/South_Africa/NHLS-UCT-GS-D026/2021\|EPI_ISL_3207548\|2021-07-02 | 21A (Delta) |
| hCoV-19/South_Africa/NHLS-UCT-GS-D027/2021\|EPI_ISL_3207549\|2021-07-02 | 21A (Delta) |
| hCoV-19/South_Africa/NHLS-UCT-GS-D028/2021\|EPI_ISL_3207511\|2021-07-02 | 21A (Delta) |
| hCoV-19/South_Africa/NHLS-UCT-GS-D030/2021\|EPI_ISL_3207550\|2021-07-02 | 21A (Delta) |
| hCoV-19/South_Africa/NHLS-UCT-GS-D031/2021\|EPI_ISL_3207551\|2021-07-02 | 21A (Delta) |
| hCoV-19/South_Africa/NHLS-UCT-GS-D032/2021\|EPI_ISL_3207552\|2021-07-02 | 21A (Delta) |
| hCoV-19/South_Africa/NHLS-UCT-GS-D033/2021\|EPI_ISL_3207553\|2021-07-02 | 21A (Delta) |
| hCoV-19/South_Africa/NHLS-UCT-GS-D034/2021\|EPI_ISL_3207554\|2021-07-05 | 21A (Delta) |
| hCoV-19/South_Africa/NHLS-UCT-GS-D035/2021\|EPI_ISL_3207555\|2021-07-05 | 21A (Delta) |
| hCoV-19/South_Africa/NHLS-UCT-GS-D036/2021\|EPI_ISL_3207556\|2021-07-06 | 21A (Delta) |
| hCoV-19/South_Africa/NHLS-UCT-GS-D037/2021\|EPI_ISL_3207557\|2021-07-05 | 21A (Delta) |
| hCoV-19/South_Africa/NHLS-UCT-GS-D038/2021\|EPI_ISL_3207558\|2021-07-05 | 21A (Delta) |
| hCoV-19/South_Africa/NHLS-UCT-GS-D040/2021\|EPI_ISL_3207559\|2021-07-05 | 21A (Delta) |
| hCoV-19/South_Africa/NHLS-UCT-GS-D041/2021\|EPI_ISL_3207560\|2021-07-06 | 21A (Delta) |
| hCoV-19/South_Africa/NHLS-UCT-GS-D042/2021\|EPI_ISL_3207512\|2021-07-06 | 21A (Delta) |
| hCoV-19/South_Africa/NHLS-UCT-GS-D043/2021\|EPI_ISL_3207561\|2021-07-06 | 21A (Delta) |
| hCoV-19/South_Africa/NHLS-UCT-GS-D044/2021\|EPI_ISL_3207562\|2021-07-06 | 21A (Delta) |
| hCoV-19/South_Africa/NHLS-UCT-GS-D046/2021\|EPI_ISL_3207563\|2021-07-07 | 21A (Delta) |
| hCoV-19/South_Africa/NHLS-UCT-GS-D047/2021\|EPI_ISL_3207564\|2021-07-07 | 21A (Delta) |
| hCoV-19/South_Africa/NHLS-UCT-GS-D048/2021\|EPI_ISL_3207565\|2021-07-08 | 21A (Delta) |
| hCoV-19/South_Africa/NHLS-UCT-GS-D049/2021\|EPI_ISL_3207566\|2021-07-08 | 21A (Delta) |
| hCoV-19/South_Africa/NHLS-UCT-GS-D050/2021\|EPI_ISL_3207567\|2021-07-08 | 21A (Delta) |
| hCoV-19/South_Africa/NHLS-UCT-GS-D051/2021\|EPI_ISL_3207568\|2021-07-08 | 21A (Delta) |
| hCoV-19/South_Africa/NHLS-UCT-GS-D052/2021\|EPI_ISL_3207569\|2021-07-08 | 21A (Delta) |
| hCoV-19/South_Africa/NHLS-UCT-GS-D053/2021\|EPI_ISL_3207513\|2021-07-09 | 20H (Beta, V2) |
| hCoV-19/South_Africa/NHLS-UCT-GS-D054/2021\|EPI_ISL_3207570\|2021-07-06 | 21A (Delta) |
| hCoV-19/South_Africa/NHLS-UCT-GS-D055/2021\|EPI_ISL_3207571\|2021-06-29 | 21A (Delta) |

| | |
|---|---|
| hCoV-19/South_Africa/NHLS-UCT-GS-D056/2021\|EPI_ISL_3207572\|2021-07-03 | 21A (Delta) |
| hCoV-19/South_Africa/NHLS-UCT-GS-D057/2021\|EPI_ISL_3207573\|2021-07-05 | 21A (Delta) |
| hCoV-19/South_Africa/NHLS-UCT-GS-D058/2021\|EPI_ISL_3207504\|2021-06-28 | 21A (Delta) |
| hCoV-19/South_Africa/NHLS-UCT-GS-D402/2021\|EPI_ISL_3506492\|2021-07-07 | 21A (Delta) |
| hCoV-19/South_Africa/NHLS-UCT-GS-D506/2021\|EPI_ISL_3957813\|2021-07-09 | 21A (Delta) |
| hCoV-19/South_Africa/NHLS-UCT-GS-D598/2021\|EPI_ISL_3506504\|2021-07-09 | 21A (Delta) |
| hCoV-19/South_Africa/NHLS-UCT-GS-D626/2021\|EPI_ISL_3506505\|2021-07-09 | 21A (Delta) |
| hCoV-19/South_Africa/NHLS-UCT-GS-D629/2021\|EPI_ISL_3506471\|2021-07-09 | 21A (Delta) |
| hCoV-19/South_Africa/NHLS-UCT-GS-D648/2021\|EPI_ISL_3506434\|2021-07-10 | 20H (Beta, V2) |
| hCoV-19/South_Africa/NHLS-UCT-GS-D664/2021\|EPI_ISL_3506506\|2021-07-10 | 21A (Delta) |
| hCoV-19/South_Africa/NHLS-UCT-GS-D705/2021\|EPI_ISL_3506493\|2021-07-12 | 21A (Delta) |
| hCoV-19/South_Africa/NHLS-UCT-GS-D730/2021\|EPI_ISL_3506472\|2021-07-12 | 21A (Delta) |
| hCoV-19/South_Africa/NHLS-UCT-GS-D737/2021\|EPI_ISL_3506507\|2021-07-12 | 21A (Delta) |
| hCoV-19/South_Africa/NHLS-UCT-GS-D755/2021\|EPI_ISL_3506509\|2021-07-12 | 21A (Delta) |
| hCoV-19/South_Africa/NHLS-UCT-GS-D767/2021\|EPI_ISL_3506510\|2021-07-12 | 21A (Delta) |
| hCoV-19/South_Africa/NHLS-UCT-GS-D779/2021\|EPI_ISL_3506511\|2021-07-12 | 21A (Delta) |
| hCoV-19/South_Africa/NHLS-UCT-GS-D781/2021\|EPI_ISL_3506479\|2021-07-13 | 21A (Delta) |
| hCoV-19/South_Africa/NHLS-UCT-GS-D793/2021\|EPI_ISL_3506387\|2021-07-12 | 21A (Delta) |
| hCoV-19/South_Africa/NHLS-UCT-GS-D803/2021\|EPI_ISL_3506381\|2021-07-12 | 21A (Delta) |
| hCoV-19/South_Africa/NHLS-UCT-GS-D811/2021\|EPI_ISL_3506368\|2021-07-12 | 21A (Delta) |
| hCoV-19/South_Africa/NHLS-UCT-GS-D858/2021\|EPI_ISL_3506512\|2021-07-13 | 21A (Delta) |
| hCoV-19/South_Africa/NHLS-UCT-GS-D866/2021\|EPI_ISL_3506513\|2021-07-13 | 21A (Delta) |
| hCoV-19/South_Africa/NHLS-UCT-GS-D876/2021\|EPI_ISL_3506514\|2021-07-13 | 21A (Delta) |
| hCoV-19/South_Africa/NHLS-UCT-GS-D892/2021\|EPI_ISL_3506515\|2021-07-14 | 21A (Delta) |
| hCoV-19/South_Africa/NHLS-UCT-GS-D897/2021\|EPI_ISL_3506516\|2021-07-14 | 21A (Delta) |
| hCoV-19/South_Africa/NHLS-UCT-GS-D909/2021\|EPI_ISL_3506484\|2021-07-14 | 21A (Delta) |
| hCoV-19/South_Africa/NHLS-UCT-GS-D911/2021\|EPI_ISL_3506517\|2021-07-14 | 21A (Delta) |
| hCoV-19/South_Africa/NHLS-UCT-GS-D922/2021\|EPI_ISL_3506364\|2021-07-14 | 21A (Delta) |
| hCoV-19/South_Africa/NHLS-UCT-GS-D923/2021\|EPI_ISL_3506485\|2021-07-14 | 21A (Delta) |
| hCoV-19/South_Africa/NHLS-UCT-GS-D948/2021\|EPI_ISL_3506473\|2021-07-14 | 21A (Delta) |
| hCoV-19/South_Africa/NHLS-UCT-GS-D958/2021\|EPI_ISL_3506412\|2021-07-14 | 21A (Delta) |
| hCoV-19/South_Africa/NHLS-UCT-GS-D999/2021\|EPI_ISL_3506518\|2021-07-15 | 21A (Delta) |
| hCoV-19/South_Africa/NHLS-UCT-GS-E010/2021\|EPI_ISL_3506486\|2021-07-15 | 21A (Delta) |
| hCoV-19/South_Africa/NHLS-UCT-GS-E012/2021\|EPI_ISL_3506474\|2021-07-15 | 21A (Delta) |
| hCoV-19/South_Africa/NHLS-UCT-GS-E045/2021\|EPI_ISL_3506409\|2021-07-15 | 21A (Delta) |
| hCoV-19/South_Africa/NHLS-UCT-GS-E080/2021\|EPI_ISL_3506379\|2021-07-16 | 21A (Delta) |
| hCoV-19/South_Africa/NHLS-UCT-GS-E089/2021\|EPI_ISL_3506519\|2021-07-16 | 21A (Delta) |
| hCoV-19/South_Africa/NHLS-UCT-GS-E090/2021\|EPI_ISL_3506494\|2021-07-16 | 21A (Delta) |
| hCoV-19/South_Africa/NHLS-UCT-GS-E106/2021\|EPI_ISL_3506487\|2021-07-13 | 21A (Delta) |
| hCoV-19/South_Africa/NHLS-UCT-GS-E116/2021\|EPI_ISL_3506488\|2021-07-12 | 21A (Delta) |
| hCoV-19/South_Africa/NHLS-UCT-GS-E129/2021\|EPI_ISL_3506480\|2021-07-15 | 21A (Delta) |
| hCoV-19/South_Africa/NHLS-UCT-GS-E140/2021\|EPI_ISL_3506385\|2021-07-12 | 21A (Delta) |
| hCoV-19/South_Africa/NHLS-UCT-GS-E150/2021\|EPI_ISL_3506495\|2021-07-09 | 21A (Delta) |
| hCoV-19/South_Africa/NHLS-UCT-GS-E168/2021\|EPI_ISL_3506520\|2021-07-16 | 21A (Delta) |
| hCoV-19/South_Africa/NHLS-UCT-GS-E169/2021\|EPI_ISL_3506521\|2021-07-19 | 21A (Delta) |
| hCoV-19/South_Africa/NHLS-UCT-GS-E170/2021\|EPI_ISL_3506413\|2021-07-19 | 21A (Delta) |
| hCoV-19/South_Africa/NHLS-UCT-GS-E171/2021\|EPI_ISL_3506522\|2021-07-19 | 21A (Delta) |
| hCoV-19/South_Africa/NHLS-UCT-GS-E173/2021\|EPI_ISL_3506561\|2021-07-19 | 21A (Delta) |
| hCoV-19/South_Africa/NHLS-UCT-GS-E174/2021\|EPI_ISL_3506523\|2021-07-19 | 21A (Delta) |
| hCoV-19/South_Africa/NHLS-UCT-GS-E175/2021\|EPI_ISL_3506524\|2021-07-19 | 21A (Delta) |

| | |
|---|---|
| hCoV-19/South_Africa/NHLS-UCT-GS-E176/2021\|EPI_ISL_3506525\|2021-07-20 | 21A (Delta) |
| hCoV-19/South_Africa/NHLS-UCT-GS-E177/2021\|EPI_ISL_3506526\|2021-07-20 | 21A (Delta) |
| hCoV-19/South_Africa/NHLS-UCT-GS-E178/2021\|EPI_ISL_3506375\|2021-07-20 | 21A (Delta) |
| hCoV-19/South_Africa/NHLS-UCT-GS-E179/2021\|EPI_ISL_3506425\|2021-07-19 | 21A (Delta) |
| hCoV-19/South_Africa/NHLS-UCT-GS-E180/2021\|EPI_ISL_3506527\|2021-07-20 | 21A (Delta) |
| hCoV-19/South_Africa/NHLS-UCT-GS-E181/2021\|EPI_ISL_3506528\|2021-07-19 | 21A (Delta) |
| hCoV-19/South_Africa/NHLS-UCT-GS-E182/2021\|EPI_ISL_3506388\|2021-07-19 | 21A (Delta) |
| hCoV-19/South_Africa/NHLS-UCT-GS-E183/2021\|EPI_ISL_3506529\|2021-07-20 | 21A (Delta) |
| hCoV-19/South_Africa/NHLS-UCT-GS-E184/2021\|EPI_ISL_3506530\|2021-07-20 | 21A (Delta) |
| hCoV-19/South_Africa/NHLS-UCT-GS-E185/2021\|EPI_ISL_3506531\|2021-07-21 | 21A (Delta) |
| hCoV-19/South_Africa/NHLS-UCT-GS-E186/2021\|EPI_ISL_3506532\|2021-07-20 | 21A (Delta) |
| hCoV-19/South_Africa/NHLS-UCT-GS-E187/2021\|EPI_ISL_3506533\|2021-07-21 | 21A (Delta) |
| hCoV-19/South_Africa/NHLS-UCT-GS-E188/2021\|EPI_ISL_3506534\|2021-07-21 | 21A (Delta) |
| hCoV-19/South_Africa/NHLS-UCT-GS-E189/2021\|EPI_ISL_3506535\|2021-07-22 | 21A (Delta) |
| hCoV-19/South_Africa/NHLS-UCT-GS-E190/2021\|EPI_ISL_3506536\|2021-07-22 | 21A (Delta) |
| hCoV-19/South_Africa/NHLS-UCT-GS-E191/2021\|EPI_ISL_3506537\|2021-07-22 | 21A (Delta) |
| hCoV-19/South_Africa/NHLS-UCT-GS-E192/2021\|EPI_ISL_3506538\|2021-07-21 | 21A (Delta) |
| hCoV-19/South_Africa/NHLS-UCT-GS-E193/2021\|EPI_ISL_3506414\|2021-07-22 | 21A (Delta) |
| hCoV-19/South_Africa/NHLS-UCT-GS-E206/2021\|EPI_ISL_3690619\|2021-07-16 | 21A (Delta) |
| hCoV-19/South_Africa/NHLS-UCT-GS-E217/2021\|EPI_ISL_3957814\|2021-07-17 | 21A (Delta) |
| hCoV-19/South_Africa/NHLS-UCT-GS-E219/2021\|EPI_ISL_3690573\|2021-07-16 | 21A (Delta) |
| hCoV-19/South_Africa/NHLS-UCT-GS-E263/2021\|EPI_ISL_3957815\|2021-07-19 | 21A (Delta) |
| hCoV-19/South_Africa/NHLS-UCT-GS-E264/2021\|EPI_ISL_3957816\|2021-07-19 | 21A (Delta) |
| hCoV-19/South_Africa/NHLS-UCT-GS-E320/2021\|EPI_ISL_3957817\|2021-07-20 | 21A (Delta) |
| hCoV-19/South_Africa/NHLS-UCT-GS-E335/2021\|EPI_ISL_3690579\|2021-07-19 | 21A (Delta) |
| hCoV-19/South_Africa/NHLS-UCT-GS-E370/2021\|EPI_ISL_3690617\|2021-07-20 | 21A (Delta) |
| hCoV-19/South_Africa/NHLS-UCT-GS-E494/2021\|EPI_ISL_3690572\|2021-07-20 | 21A (Delta) |
| hCoV-19/South_Africa/NHLS-UCT-GS-E638/2021\|EPI_ISL_3690618\|2021-07-19 | 21A (Delta) |
| hCoV-19/South_Africa/NHLS-UCT-GS-E773/2021\|EPI_ISL_3690613\|2021-07-23 | 21A (Delta) |
| hCoV-19/South_Africa/NHLS-UCT-GS-E799/2021\|EPI_ISL_3690614\|2021-07-22 | 21A (Delta) |
| hCoV-19/South_Africa/NHLS-UCT-GS-E809/2021\|EPI_ISL_3690571\|2021-07-24 | 21A (Delta) |
| hCoV-19/South_Africa/NHLS-UCT-GS-E853/2021\|EPI_ISL_3690612\|2021-07-25 | 21A (Delta) |
| hCoV-19/South_Africa/NHLS-UCT-GS-E905/2021\|EPI_ISL_3690609\|2021-07-26 | 21A (Delta) |
| hCoV-19/South_Africa/NHLS-UCT-GS-E923/2021\|EPI_ISL_3957818\|2021-07-26 | 20H (Beta, V2) |
| hCoV-19/South_Africa/NHLS-UCT-GS-E925/2021\|EPI_ISL_3690610\|2021-07-26 | 21A (Delta) |
| hCoV-19/South_Africa/NHLS-UCT-GS-E938/2021\|EPI_ISL_3957819\|2021-07-26 | 21A (Delta) |
| hCoV-19/South_Africa/NHLS-UCT-GS-E976/2021\|EPI_ISL_3690575\|2021-07-26 | 21A (Delta) |
| hCoV-19/South_Africa/NHLS-UCT-GS-E984/2021\|EPI_ISL_3690611\|2021-07-26 | 21A (Delta) |
| hCoV-19/South_Africa/NHLS-UCT-GS-F061/2021\|EPI_ISL_3690604\|2021-07-28 | 21A (Delta) |
| hCoV-19/South_Africa/NHLS-UCT-GS-F076/2021\|EPI_ISL_3690605\|2021-07-28 | 21A (Delta) |
| hCoV-19/South_Africa/NHLS-UCT-GS-F085/2021\|EPI_ISL_3690608\|2021-07-27 | 21A (Delta) |
| hCoV-19/South_Africa/NHLS-UCT-GS-F089/2021\|EPI_ISL_3690570\|2021-07-27 | 21A (Delta) |
| hCoV-19/South_Africa/NHLS-UCT-GS-F132/2021\|EPI_ISL_3690606\|2021-07-28 | 21A (Delta) |
| hCoV-19/South_Africa/NHLS-UCT-GS-F147/2021\|EPI_ISL_3957820\|2021-07-29 | 21A (Delta) |
| hCoV-19/South_Africa/NHLS-UCT-GS-F429/2021\|EPI_ISL_3690545\|2021-08-01 | 21A (Delta) |
| hCoV-19/South_Africa/NHLS-UCT-GS-F461/2021\|EPI_ISL_3690603\|2021-08-01 | 21A (Delta) |
| hCoV-19/South_Africa/NHLS-UCT-GS-F494/2021\|EPI_ISL_3690554\|2021-08-02 | 21A (Delta) |
| hCoV-19/South_Africa/NHLS-UCT-GS-F504/2021\|EPI_ISL_3690599\|2021-08-02 | 21A (Delta) |
| hCoV-19/South_Africa/NHLS-UCT-GS-F522/2021\|EPI_ISL_3690600\|2021-08-02 | 21A (Delta) |
| hCoV-19/South_Africa/NHLS-UCT-GS-F523/2021\|EPI_ISL_3690601\|2021-08-02 | 21A (Delta) |

| | |
|---|---|
| hCoV-19/South_Africa/NHLS-UCT-GS-F557/2021\|EPI_ISL_3690582\|2021-08-04 | 21A (Delta) |
| hCoV-19/South_Africa/NHLS-UCT-GS-F572/2021\|EPI_ISL_3690566\|2021-08-03 | 21A (Delta) |
| hCoV-19/South_Africa/NHLS-UCT-GS-F574/2021\|EPI_ISL_3690583\|2021-08-04 | 21A (Delta) |
| hCoV-19/South_Africa/NHLS-UCT-GS-F579/2021\|EPI_ISL_3690584\|2021-08-04 | 21A (Delta) |
| hCoV-19/South_Africa/NHLS-UCT-GS-F581/2021\|EPI_ISL_3690585\|2021-08-04 | 21A (Delta) |
| hCoV-19/South_Africa/NHLS-UCT-GS-F591/2021\|EPI_ISL_3690591\|2021-08-03 | 21A (Delta) |
| hCoV-19/South_Africa/NHLS-UCT-GS-F603/2021\|EPI_ISL_3690586\|2021-08-04 | 21A (Delta) |
| hCoV-19/South_Africa/NHLS-UCT-GS-F605/2021\|EPI_ISL_3690587\|2021-08-04 | 21A (Delta) |
| hCoV-19/South_Africa/NHLS-UCT-GS-F628/2021\|EPI_ISL_3690565\|2021-08-04 | 21A (Delta) |
| hCoV-19/South_Africa/NHLS-UCT-GS-F647/2021\|EPI_ISL_3690548\|2021-08-03 | 21A (Delta) |
| hCoV-19/South_Africa/NHLS-UCT-GS-F682/2021\|EPI_ISL_3690580\|2021-08-05 | 21A (Delta) |
| hCoV-19/South_Africa/NHLS-UCT-GS-F705/2021\|EPI_ISL_3690581\|2021-08-05 | 21A (Delta) |
| hCoV-19/South_Africa/NHLS-UCT-GS-F763/2021\|EPI_ISL_3690549\|2021-08-04 | 21A (Delta) |
| hCoV-19/South_Africa/NHLS-UCT-GS-F781/2021\|EPI_ISL_3690588\|2021-08-04 | 21A (Delta) |
| hCoV-19/South_Africa/NHLS-UCT-GS-F790/2021\|EPI_ISL_3957821\|2021-08-04 | 20B |
| hCoV-19/South_Africa/NHLS-UCT-GS-F793/2021\|EPI_ISL_3957822\|2021-08-04 | 21A (Delta) |
| hCoV-19/South_Africa/NHLS-UCT-GS-F795/2021\|EPI_ISL_3690592\|2021-08-03 | 21A (Delta) |
| hCoV-19/South_Africa/NHLS-UCT-GS-F799/2021\|EPI_ISL_3690589\|2021-08-04 | 21A (Delta) |
| hCoV-19/South_Africa/NHLS-UCT-GS-F849/2021\|EPI_ISL_3957823\|2021-08-09 | 21A (Delta) |
| hCoV-19/South_Africa/NHLS-UCT-GS-F853/2021\|EPI_ISL_3957824\|2021-08-09 | 21A (Delta) |
| hCoV-19/South_Africa/NHLS-UCT-GS-F859/2021\|EPI_ISL_3957825\|2021-08-09 | 21A (Delta) |
| hCoV-19/South_Africa/NHLS-UCT-GS-F861/2021\|EPI_ISL_3957826\|2021-08-09 | 21A (Delta) |
| hCoV-19/South_Africa/NHLS-UCT-GS-F918/2021\|EPI_ISL_3957827\|2021-08-10 | 21A (Delta) |
| hCoV-19/South_Africa/NHLS-UCT-GS-F924/2021\|EPI_ISL_3957828\|2021-08-10 | 21A (Delta) |
| hCoV-19/South_Africa/NHLS-UCT-GS-F941/2021\|EPI_ISL_3957829\|2021-08-10 | 21A (Delta) |
| hCoV-19/South_Africa/NHLS-UCT-GS-F962/2021\|EPI_ISL_3957830\|2021-08-11 | 21A (Delta) |
| hCoV-19/South_Africa/NHLS-UCT-GS-F971/2021\|EPI_ISL_3957831\|2021-08-10 | 21A (Delta) |
| hCoV-19/South_Africa/NHLS-UCT-GS-F973/2021\|EPI_ISL_3957832\|2021-08-10 | 21A (Delta) |
| hCoV-19/South_Africa/NHLS-UCT-GS-F980/2021\|EPI_ISL_3957833\|2021-08-11 | 21A (Delta) |
| hCoV-19/South_Africa/NHLS-UCT-GS-F986/2021\|EPI_ISL_3957834\|2021-08-11 | 21A (Delta) |
| hCoV-19/South_Africa/NHLS-UCT-GS-F991/2021\|EPI_ISL_3957835\|2021-08-10 | 21A (Delta) |
| hCoV-19/South_Africa/NHLS-UCT-GS-F997/2021\|EPI_ISL_3957836\|2021-08-11 | 21A (Delta) |
| hCoV-19/South_Africa/NHLS-UCT-GS-G013/2021\|EPI_ISL_3957837\|2021-08-11 | 21A (Delta) |
| hCoV-19/South_Africa/NHLS-UCT-GS-G015/2021\|EPI_ISL_3957838\|2021-08-11 | 21A (Delta) |
| hCoV-19/South_Africa/NHLS-UCT-GS-G021/2021\|EPI_ISL_3957839\|2021-08-11 | 21A (Delta) |
| hCoV-19/South_Africa/NHLS-UCT-GS-G023/2021\|EPI_ISL_3957840\|2021-08-11 | 21A (Delta) |
| hCoV-19/South_Africa/NHLS-UCT-GS-G036/2021\|EPI_ISL_3957841\|2021-08-12 | 21A (Delta) |
| hCoV-19/South_Africa/NHLS-UCT-GS-G059/2021\|EPI_ISL_3957842\|2021-08-11 | 21A (Delta) |
| hCoV-19/South_Africa/NHLS-UCT-GS-G060/2021\|EPI_ISL_3957843\|2021-08-11 | 21A (Delta) |
| hCoV-19/South_Africa/NHLS-UCT-GS-G062/2021\|EPI_ISL_3957844\|2021-08-12 | 21A (Delta) |
| hCoV-19/South_Africa/NHLS-UCT-GS-G083/2021\|EPI_ISL_3957845\|2021-08-13 | 21A (Delta) |
| hCoV-19/South_Africa/NHLS-UCT-GS-G084/2021\|EPI_ISL_3957846\|2021-08-13 | 21A (Delta) |
| hCoV-19/South_Africa/NHLS-UCT-GS-G097/2021\|EPI_ISL_3957847\|2021-08-13 | 21A (Delta) |
| hCoV-19/South_Africa/NHLS-UCT-GS-G103/2021\|EPI_ISL_3957848\|2021-08-13 | 21A (Delta) |
| hCoV-19/South_Africa/NHLS-UCT-GS-G107/2021\|EPI_ISL_3957849\|2021-08-12 | 21A (Delta) |
| hCoV-19/South_Africa/NHLS-UCT-GS-G120/2021\|EPI_ISL_3957850\|2021-08-09 | 21A (Delta) |
| hCoV-19/South_Africa/NHLS-UCT-GS-G145/2021\|EPI_ISL_3957851\|2021-08-07 | 21A (Delta) |
| hCoV-19/South_Africa/NHLS-UCT-GS-G146/2021\|EPI_ISL_3957852\|2021-08-06 | 21A (Delta) |
| hCoV-19/South_Africa/NHLS-UCT-GS-G157/2021\|EPI_ISL_3957853\|2021-08-10 | 21A (Delta) |
| hCoV-19/South_Africa/NHLS-UCT-LA-Z501/2021\|EPI_ISL_2802196\|2021-06-01 | 21A (Delta) |

| | |
|---|---|
| hCoV-19/South_Africa/NHLS-UCT-LA-Z502/2021\|EPI_ISL_2802197\|2021-06-01 | 20H (Beta, V2) |
| hCoV-19/South_Africa/NHLS-UCT-LA-Z503/2021\|EPI_ISL_2802198\|2021-06-01 | 20H (Beta, V2) |
| hCoV-19/South_Africa/NHLS-UCT-LA-Z504/2021\|EPI_ISL_2802184\|2021-06-01 | 20B |
| hCoV-19/South_Africa/NHLS-UCT-LA-Z505/2021\|EPI_ISL_2802185\|2021-06-01 | 20B |
| hCoV-19/South_Africa/NHLS-UCT-LA-Z506/2021\|EPI_ISL_2802183\|2021-06-01 | 20H (Beta, V2) |
| hCoV-19/South_Africa/NHLS-UCT-LA-Z512/2021\|EPI_ISL_3506357\|2021-06-22 | 21A (Delta) |
| hCoV-19/South_Africa/NHLS-UCT-LA-Z513/2021\|EPI_ISL_3506406\|2021-06-22 | 21A (Delta) |
| hCoV-19/South_Africa/NHLS-UCT-LA-Z514/2021\|EPI_ISL_3506462\|2021-06-22 | 21A (Delta) |
| hCoV-19/South_Africa/NHLS-UCT-LA-Z515/2021\|EPI_ISL_3506407\|2021-06-22 | 21A (Delta) |
| hCoV-19/South_Africa/NHLS-UCT-LA-Z516/2021\|EPI_ISL_3506440\|2021-06-22 | 20H (Beta, V2) |
| hCoV-19/South_Africa/NHLS-UCT-LA-Z517/2021\|EPI_ISL_3506463\|2021-06-22 | 21A (Delta) |
| hCoV-19/South_Africa/NHLS-UCT-LA-Z518/2021\|EPI_ISL_3506464\|2021-06-22 | 21A (Delta) |
| hCoV-19/South_Africa/NHLS-UCT-LA-Z519/2021\|EPI_ISL_3506559\|2021-06-22 | 21A (Delta) |
| hCoV-19/South_Africa/NHLS-UCT-LA-Z520/2021\|EPI_ISL_3506398\|2021-06-22 | 21A (Delta) |
| hCoV-19/South_Africa/NHLS-UCT-LA-Z522/2021\|EPI_ISL_3506465\|2021-06-22 | 21A (Delta) |
| hCoV-19/South_Africa/NHLS-UCT-LA-Z524/2021\|EPI_ISL_3506362\|2021-06-29 | 21A (Delta) |
| hCoV-19/South_Africa/NHLS-UCT-LA-Z525/2021\|EPI_ISL_3506408\|2021-06-29 | 21A (Delta) |
| hCoV-19/South_Africa/NHLS-UCT-LA-Z526/2021\|EPI_ISL_3506399\|2021-06-29 | 21A (Delta) |
| hCoV-19/South_Africa/NHLS-UCT-LA-Z529/2021\|EPI_ISL_3506466\|2021-06-29 | 21A (Delta) |
| hCoV-19/South_Africa/NHLS-UCT-LA-Z531/2021\|EPI_ISL_3506380\|2021-06-29 | 21A (Delta) |
| hCoV-19/South_Africa/NHLS-UCT-LA-Z532/2021\|EPI_ISL_3207574\|2021-07-05 | 21A (Delta) |
| hCoV-19/South_Africa/NHLS-UCT-LA-Z533/2021\|EPI_ISL_3207575\|2021-07-05 | 21A (Delta) |
| hCoV-19/South_Africa/NHLS-UCT-LA-Z534/2021\|EPI_ISL_3207576\|2021-06-29 | 20H (Beta, V2) |
| hCoV-19/South_Africa/NHLS-UCT-LA-Z535/2021\|EPI_ISL_3207577\|2021-06-29 | 21A (Delta) |
| hCoV-19/South_Africa/NHLS-UCT-LA-Z536/2021\|EPI_ISL_3207505\|2021-07-03 | 21A (Delta) |
| hCoV-19/South_Africa/NHLS-UCT-LA-Z537/2021\|EPI_ISL_3207506\|2021-07-03 | 21A (Delta) |
| hCoV-19/South_Africa/NHLS-UCT-LA-Z538/2021\|EPI_ISL_3207507\|2021-07-03 | 21A (Delta) |
| hCoV-19/South_Africa/NHLS-UCT-LA-Z539/2021\|EPI_ISL_3207578\|2021-07-05 | 21A (Delta) |
| hCoV-19/South_Africa/NHLS-UCT-LA-Z540/2021\|EPI_ISL_3207579\|2021-07-05 | 21A (Delta) |
| hCoV-19/South_Africa/NHLS-UCT-LA-Z541/2021\|EPI_ISL_3207580\|2021-07-05 | 21A (Delta) |
| hCoV-19/South_Africa/NHLS-UCT-LA-Z542/2021\|EPI_ISL_3207581\|2021-06-08 | 20H (Beta, V2) |
| hCoV-19/South_Africa/NHLS-UCT-LA-Z543/2021\|EPI_ISL_3207514\|2021-07-05 | 20B |
| hCoV-19/South_Africa/NHLS-UCT-LA-Z544/2021\|EPI_ISL_3207508\|2021-07-05 | 21A (Delta) |
| hCoV-19/South_Africa/NHLS-UCT-LA-Z546/2021\|EPI_ISL_3207582\|2021-07-05 | 21A (Delta) |
| hCoV-19/South_Africa/NHLS-UCT-LA-Z547/2021\|EPI_ISL_3207583\|2021-07-05 | 21A (Delta) |
| hCoV-19/South_Africa/NHLS-UCT-LA-Z548/2021\|EPI_ISL_3207584\|2021-07-05 | 21A (Delta) |
| hCoV-19/South_Africa/NHLS-UCT-LA-Z549/2021\|EPI_ISL_3207585\|2021-06-08 | 21A (Delta) |
| hCoV-19/South_Africa/NHLS-UCT-LA-Z550/2021\|EPI_ISL_3207586\|2021-07-05 | 21A (Delta) |
| hCoV-19/South_Africa/NHLS-UCT-LA-Z551/2021\|EPI_ISL_3207587\|2021-07-05 | 21A (Delta) |
| hCoV-19/South_Africa/NHLS-UCT-LA-Z553/2021\|EPI_ISL_3506489\|2021-07-14 | 21A (Delta) |
| hCoV-19/South_Africa/NHLS-UCT-LA-Z554/2021\|EPI_ISL_3506377\|2021-07-14 | 21A (Delta) |
| hCoV-19/South_Africa/NHLS-UCT-LA-Z555/2021\|EPI_ISL_3506560\|2021-07-14 | 21A (Delta) |
| hCoV-19/South_Africa/NHLS-UCT-LA-Z556/2021\|EPI_ISL_3506410\|2021-07-14 | 21A (Delta) |
| hCoV-19/South_Africa/NHLS-UCT-LA-Z557/2021\|EPI_ISL_3506415\|2021-07-14 | 21A (Delta) |
| hCoV-19/South_Africa/NHLS-UCT-LA-Z558/2021\|EPI_ISL_3506411\|2021-07-14 | 21A (Delta) |
| hCoV-19/South_Africa/NHLS-UCT-LA-Z559/2021\|EPI_ISL_3506371\|2021-07-14 | 21A (Delta) |
| hCoV-19/South_Africa/NHLS-UCT-LA-Z560/2021\|EPI_ISL_3506490\|2021-07-14 | 21A (Delta) |
| hCoV-19/South_Africa/NHLS-UCT-LA-Z561/2021\|EPI_ISL_3506540\|2021-07-14 | 21A (Delta) |
| hCoV-19/South_Africa/NHLS-UCT-LA-Z562/2021\|EPI_ISL_3506481\|2021-07-14 | 21A (Delta) |
| hCoV-19/South_Africa/NHLS-UCT-LA-Z563/2021\|EPI_ISL_3506541\|2021-07-14 | 21A (Delta) |

| | |
|---|---|
| hCoV-19/South_Africa/NHLS-UCT-LA-Z564/2021\|EPI_ISL_3506416\|2021-07-21 | 21A (Delta) |
| hCoV-19/South_Africa/NHLS-UCT-LA-Z565/2021\|EPI_ISL_3506542\|2021-07-21 | 21A (Delta) |
| hCoV-19/South_Africa/NHLS-UCT-LA-Z566/2021\|EPI_ISL_3506543\|2021-07-28 | 21A (Delta) |
| hCoV-19/South_Africa/NHLS-UCT-LA-Z567/2021\|EPI_ISL_3506544\|2021-07-28 | 21A (Delta) |
| hCoV-19/South_Africa/NHLS-UCT-LA-Z570/2021\|EPI_ISL_3506545\|2021-07-27 | 21A (Delta) |
| hCoV-19/South_Africa/NHLS-UCT-LA-Z571/2021\|EPI_ISL_3506546\|2021-07-21 | 21A (Delta) |
| hCoV-19/South_Africa/NHLS-UCT-LA-Z572/2021\|EPI_ISL_3506547\|2021-07-28 | 21A (Delta) |
| hCoV-19/South_Africa/NHLS-UCT-LA-Z574/2021\|EPI_ISL_3506548\|2021-07-28 | 21A (Delta) |
| hCoV-19/South_Africa/NHLS-UCT-LA-Z576/2021\|EPI_ISL_3506549\|2021-07-28 | 21A (Delta) |
| hCoV-19/South_Africa/NHLS-UCT-LA-Z577/2021\|EPI_ISL_3506550\|2021-07-28 | 21A (Delta) |
| hCoV-19/South_Africa/NHLS-UCT-LA-Z578/2021\|EPI_ISL_3506551\|2021-07-28 | 21A (Delta) |
| hCoV-19/South_Africa/NHLS-UCT-LA-Z579/2021\|EPI_ISL_3506552\|2021-07-21 | 21A (Delta) |
| hCoV-19/South_Africa/NHLS-UCT-LA-Z580/2021\|EPI_ISL_3506553\|2021-07-28 | 21A (Delta) |
| hCoV-19/South_Africa/NHLS-UCT-LA-Z581/2021\|EPI_ISL_3506554\|2021-07-21 | 21A (Delta) |
| hCoV-19/South_Africa/NHLS-UCT-LA-Z583/2021\|EPI_ISL_3506417\|2021-07-28 | 21A (Delta) |
| hCoV-19/South_Africa/NHLS-UCT-LA-Z584/2021\|EPI_ISL_3506418\|2021-07-21 | 21A (Delta) |
| hCoV-19/South_Africa/NHLS-UCT-LA-Z589/2021\|EPI_ISL_3690567\|2021-08-03 | 21A (Delta) |
| hCoV-19/South_Africa/NHLS-UCT-LA-Z590/2021\|EPI_ISL_3690568\|2021-08-03 | 21A (Delta) |
| hCoV-19/South_Africa/NHLS-UCT-LA-Z591/2021\|EPI_ISL_3690593\|2021-08-03 | 21A (Delta) |
| hCoV-19/South_Africa/NHLS-UCT-LA-Z592/2021\|EPI_ISL_3690569\|2021-08-03 | 21A (Delta) |
| hCoV-19/South_Africa/NHLS-UCT-LA-Z594/2021\|EPI_ISL_3690590\|2021-08-03 | 21A (Delta) |
| hCoV-19/South_Africa/NHLS-UCT-LA-Z595/2021\|EPI_ISL_3690594\|2021-08-03 | 21A (Delta) |
| hCoV-19/South_Africa/NHLS-UCT-LA-Z596/2021\|EPI_ISL_3690595\|2021-08-03 | 21A (Delta) |
| hCoV-19/South_Africa/NHLS-UCT-LA-Z597/2021\|EPI_ISL_3690596\|2021-08-03 | 21A (Delta) |
| hCoV-19/South_Africa/NHLS-UCT-LA-Z598/2021\|EPI_ISL_3690597\|2021-08-03 | 21A (Delta) |
| hCoV-19/South_Africa/NHLS-UCT-LA-Z599/2021\|EPI_ISL_3690598\|2021-08-03 | 21A (Delta) |
| hCoV-19/South_Africa/NHLS-UCT-LA-Z601/2021\|EPI_ISL_3690560\|2021-07-21 | 21A (Delta) |
| hCoV-19/South_Africa/NHLS-UCT-LA-Z602/2021\|EPI_ISL_3690615\|2021-07-21 | 21A (Delta) |
| hCoV-19/South_Africa/NHLS-UCT-LA-Z603/2021\|EPI_ISL_3690616\|2021-07-21 | 21A (Delta) |
| hCoV-19/South_Africa/NHLS-UCT-LA-Z604/2021\|EPI_ISL_3690578\|2021-07-28 | 21A (Delta) |
| hCoV-19/South_Africa/NHLS-UCT-LA-Z605/2021\|EPI_ISL_3690607\|2021-07-28 | 21A (Delta) |
| hCoV-19/South_Africa/NHLS-UCT-LA-Z606/2021\|EPI_ISL_3957854\|2021-08-11 | 21A (Delta) |
| hCoV-19/South_Africa/NHLS-UCT-LA-Z607/2021\|EPI_ISL_3957855\|2021-08-11 | 21A (Delta) |
| hCoV-19/South_Africa/NHLS-UCT-LA-Z608/2021\|EPI_ISL_3957856\|2021-08-11 | 21A (Delta) |
| hCoV-19/South_Africa/NHLS-UCT-LA-Z609/2021\|EPI_ISL_3957857\|2021-08-11 | 21A (Delta) |
| hCoV-19/South_Africa/NHLS-UCT-LA-Z610/2021\|EPI_ISL_3957858\|2021-08-11 | 21A (Delta) |
| hCoV-19/South_Africa/NHLS-UCT-LA-Z612/2021\|EPI_ISL_3957859\|2021-08-11 | 21A (Delta) |
| hCoV-19/South_Africa/NHLS-UCT-LA-Z613/2021\|EPI_ISL_3957860\|2021-08-11 | 21A (Delta) |
| hCoV-19/South_Africa/NHLS-UCT-LA-Z614/2021\|EPI_ISL_3957861\|2021-08-11 | 21A (Delta) |
| hCoV-19/South_Africa/NHLS-UCT-LA-Z615/2021\|EPI_ISL_3957862\|2021-08-11 | 21A (Delta) |
| hCoV-19/South_Africa/NHLS-UCT-LA-Z616/2021\|EPI_ISL_3957863\|2021-08-11 | 21A (Delta) |
| hCoV-19/South_Africa/NHLS-UCT-LA-Z617/2021\|EPI_ISL_3957864\|2021-08-11 | 21A (Delta) |
| hCoV-19/South_Africa/NHLS-UCT-PA-J001/2021\|EPI_ISL_3207489\|2021-03-19 | 20H (Beta, V2) |
| hCoV-19/South_Africa/NHLS-UCT-PA-J003/2021\|EPI_ISL_3207494\|2021-03-29 | 20H (Beta, V2) |
| hCoV-19/South_Africa/NHLS-UCT-PA-J004/2021\|EPI_ISL_3207490\|2021-03-20 | 20H (Beta, V2) |
| hCoV-19/South_Africa/NHLS-UCT-PA-J005/2021\|EPI_ISL_3207491\|2021-03-29 | 20H (Beta, V2) |
| hCoV-19/South_Africa/NHLS-UCT-PA-J006/2021\|EPI_ISL_3207492\|2021-03-15 | 20H (Beta, V2) |
| hCoV-19/South_Africa/NHLS-UCT-PA-J007/2021\|EPI_ISL_3207495\|2021-03-25 | 20I (Alpha, V1) |
| hCoV-19/South_Africa/NHLS-UCT-PA-J008/2021\|EPI_ISL_3207496\|2021-04-16 | 20H (Beta, V2) |
| hCoV-19/South_Africa/NHLS-UCT-PA-J009/2021\|EPI_ISL_3207497\|2021-04-27 | 20H (Beta, V2) |

| | |
|---|---|
| hCoV-19/South_Africa/NHLS-UCT-PA-J010/2021\|EPI_ISL_3207498\|2021-04-21 | 20H (Beta, V2) |
| hCoV-19/South_Africa/NICD-CRDM11977/2021\|EPI_ISL_3718067\|2021-08-10 | 21A (Delta) |
| hCoV-19/South_Africa/NICD-CRDM12009/2021\|EPI_ISL_3718068\|2021-08-11 | 21A (Delta) |
| hCoV-19/South_Africa/NICD-CRDM12153/2021\|EPI_ISL_3718069\|2021-08-14 | 21A (Delta) |
| hCoV-19/South_Africa/NICD-N7458/2021\|EPI_ISL_3030526\|2021-05-12 | 21A (Delta) |
| hCoV-19/South_Africa/NICD-N7467/2021\|EPI_ISL_3030529\|2021-05 | 20C |
| hCoV-19/South_Africa/NICD-R10375/2021\|EPI_ISL_3101646\|2021-07-12 | 21A (Delta) |
| hCoV-19/South_Africa/NICD-R10437/2021\|EPI_ISL_3281587\|2021-07-12 | 21A (Delta) |
| hCoV-19/South_Africa/NICD-R12181/2021\|EPI_ISL_3717923\|2021-08-13 | 21A (Delta) |
| hCoV-19/South_Africa/NICD-R12183/2021\|EPI_ISL_3717914\|2021-08-13 | 21A (Delta) |
| hCoV-19/South_Africa/NICD-R12191/2021\|EPI_ISL_3717956\|2021-08-13 | 21A (Delta) |
| hCoV-19/South_Africa/Tygerberg_1064/2021\|EPI_ISL_2695811\|2021-06-02 | 20H (Beta, V2) |
| hCoV-19/South_Africa/Tygerberg_1065/2021\|EPI_ISL_2695812\|2021-06-02 | 20H (Beta, V2) |
| hCoV-19/South_Africa/Tygerberg_1066/2021\|EPI_ISL_2695813\|2021-06-02 | 20H (Beta, V2) |
| hCoV-19/South_Africa/Tygerberg_1067/2021\|EPI_ISL_2695814\|2021-06-02 | 20H (Beta, V2) |
| hCoV-19/South_Africa/Tygerberg_1068/2021\|EPI_ISL_2695815\|2021-06-02 | 20H (Beta, V2) |
| hCoV-19/South_Africa/Tygerberg_1069/2021\|EPI_ISL_2695816\|2021-06-02 | 20H (Beta, V2) |
| hCoV-19/South_Africa/Tygerberg_1070/2021\|EPI_ISL_2695817\|2021-06-02 | 21A (Delta) |
| hCoV-19/South_Africa/Tygerberg_1071/2021\|EPI_ISL_2695818\|2021-06-02 | 21A (Delta) |
| hCoV-19/South_Africa/Tygerberg_1072/2021\|EPI_ISL_2695819\|2021-06-02 | 20I (Alpha, V1) |
| hCoV-19/South_Africa/Tygerberg_1073/2021\|EPI_ISL_2695820\|2021-06-02 | 21A (Delta) |
| hCoV-19/South_Africa/Tygerberg_1074/2021\|EPI_ISL_2695821\|2021-05-24 | 21A (Delta) |
| hCoV-19/South_Africa/Tygerberg_1075/2021\|EPI_ISL_2695822\|2021-05-27 | 21A (Delta) |
| hCoV-19/South_Africa/Tygerberg_1076/2021\|EPI_ISL_2695823\|2021-05-27 | 20H (Beta, V2) |
| hCoV-19/South_Africa/Tygerberg_1077/2021\|EPI_ISL_2695824\|2021-05-27 | 20H (Beta, V2) |
| hCoV-19/South_Africa/Tygerberg_1078/2021\|EPI_ISL_2695825\|2021-05-27 | 20B |
| hCoV-19/South_Africa/Tygerberg_1079/2021\|EPI_ISL_2695826\|2021-05-27 | 20H (Beta, V2) |
| hCoV-19/South_Africa/Tygerberg_1080/2021\|EPI_ISL_2695827\|2021-05-27 | 20H (Beta, V2) |
| hCoV-19/South_Africa/Tygerberg_1081/2021\|EPI_ISL_2695828\|2021-05-27 | 20H (Beta, V2) |
| hCoV-19/South_Africa/Tygerberg_1082/2021\|EPI_ISL_2695829\|2021-05-27 | 21A (Delta) |
| hCoV-19/South_Africa/Tygerberg_1083/2021\|EPI_ISL_2695830\|2021-05-27 | 20H (Beta, V2) |
| hCoV-19/South_Africa/Tygerberg_1084/2021\|EPI_ISL_2695831\|2021-05-27 | 20H (Beta, V2) |
| hCoV-19/South_Africa/Tygerberg_1085/2021\|EPI_ISL_2695832\|2021-05-27 | 20H (Beta, V2) |
| hCoV-19/South_Africa/Tygerberg_1086/2021\|EPI_ISL_2695833\|2021-05-13 | 20H (Beta, V2) |
| hCoV-19/South_Africa/Tygerberg_1087/2021\|EPI_ISL_2695834\|2021-05-18 | 20B |
| hCoV-19/South_Africa/Tygerberg_1088/2021\|EPI_ISL_2695835\|2021-05-20 | 20H (Beta, V2) |
| hCoV-19/South_Africa/Tygerberg_1090/2021\|EPI_ISL_2695837\|2021-06-08 | 20H (Beta, V2) |
| hCoV-19/South_Africa/Tygerberg_1091/2021\|EPI_ISL_2695838\|2021-06-09 | 20H (Beta, V2) |
| hCoV-19/South_Africa/Tygerberg_1092/2021\|EPI_ISL_2695839\|2021-06-09 | 20H (Beta, V2) |
| hCoV-19/South_Africa/Tygerberg_1093/2021\|EPI_ISL_2695840\|2021-06-09 | 20H (Beta, V2) |
| hCoV-19/South_Africa/Tygerberg_1094/2021\|EPI_ISL_2695841\|2021-06-09 | 21A (Delta) |
| hCoV-19/South_Africa/Tygerberg_1095/2021\|EPI_ISL_2695842\|2021-06-09 | 21A (Delta) |
| hCoV-19/South_Africa/Tygerberg_1096/2021\|EPI_ISL_2695843\|2021-06-09 | 21A (Delta) |
| hCoV-19/South_Africa/Tygerberg_1097/2021\|EPI_ISL_2695844\|2021-06-09 | 21A (Delta) |
| hCoV-19/South_Africa/Tygerberg_1098/2021\|EPI_ISL_2695845\|2021-06-09 | 21A (Delta) |
| hCoV-19/South_Africa/Tygerberg_1099/2021\|EPI_ISL_2695846\|2021-06-09 | 20I (Alpha, V1) |
| hCoV-19/South_Africa/Tygerberg_1100/2021\|EPI_ISL_2695847\|2021-06-09 | 21A (Delta) |
| hCoV-19/South_Africa/Tygerberg_1101/2021\|EPI_ISL_2695848\|2021-06-09 | 20H (Beta, V2) |
| hCoV-19/South_Africa/Tygerberg_1105/2021\|EPI_ISL_2695849\|2021-05-19 | 20H (Beta, V2) |
| hCoV-19/South_Africa/Tygerberg_1107/2021\|EPI_ISL_2695850\|2021-05-18 | 20H (Beta, V2) |

| | |
|---|---|
| hCoV-19/South_Africa/Tygerberg_1108/2021\|EPI_ISL_2695851\|2021-05-19 | 20H (Beta, V2) |
| hCoV-19/South_Africa/Tygerberg_1110/2021\|EPI_ISL_2695852\|2021-05-08 | 20H (Beta, V2) |
| hCoV-19/South_Africa/Tygerberg_1112/2021\|EPI_ISL_2695853\|2021-05-02 | 20I (Alpha, V1) |
| hCoV-19/South_Africa/Tygerberg_1113/2021\|EPI_ISL_2695854\|2021-05-02 | 20H (Beta, V2) |
| hCoV-19/South_Africa/Tygerberg_1114/2021\|EPI_ISL_2695855\|2021-05-14 | 20B |
| hCoV-19/South_Africa/Tygerberg_1115/2021\|EPI_ISL_2695856\|2021-05-13 | 20H (Beta, V2) |
| hCoV-19/South_Africa/Tygerberg_1116/2021\|EPI_ISL_2695857\|2021-05-12 | 20H (Beta, V2) |
| hCoV-19/South_Africa/Tygerberg_1117/2021\|EPI_ISL_2695858\|2021-05-12 | 20H (Beta, V2) |
| hCoV-19/South_Africa/Tygerberg_1118/2021\|EPI_ISL_2695859\|2021-05-18 | 20H (Beta, V2) |
| hCoV-19/South_Africa/Tygerberg_1120/2021\|EPI_ISL_2695860\|2021-05-14 | 21D (Eta) |
| hCoV-19/South_Africa/Tygerberg_1121/2021\|EPI_ISL_2695861\|2021-05-14 | 20H (Beta, V2) |
| hCoV-19/South_Africa/Tygerberg_1122/2021\|EPI_ISL_2695862\|2021-05-12 | 20H (Beta, V2) |
| hCoV-19/South_Africa/Tygerberg_1123/2021\|EPI_ISL_2695863\|2021-05-15 | 20H (Beta, V2) |
| hCoV-19/South_Africa/Tygerberg_1124/2021\|EPI_ISL_2695864\|2021-05-07 | 20H (Beta, V2) |
| hCoV-19/South_Africa/Tygerberg_1126/2021\|EPI_ISL_2695865\|2021-05-10 | 20H (Beta, V2) |
| hCoV-19/South_Africa/Tygerberg_1127/2021\|EPI_ISL_2695866\|2021-05-18 | 20H (Beta, V2) |
| hCoV-19/South_Africa/Tygerberg_1128/2021\|EPI_ISL_2695867\|2021-05-12 | 20H (Beta, V2) |
| hCoV-19/South_Africa/Tygerberg_1129/2021\|EPI_ISL_2695868\|2021-05-12 | 20H (Beta, V2) |
| hCoV-19/South_Africa/Tygerberg_1131/2021\|EPI_ISL_2695869\|2021-05-11 | 20H (Beta, V2) |
| hCoV-19/South_Africa/Tygerberg_1132/2021\|EPI_ISL_2695870\|2021-04-21 | 20H (Beta, V2) |
| hCoV-19/South_Africa/Tygerberg_1134/2021\|EPI_ISL_2695871\|2021-04-22 | 20H (Beta, V2) |
| hCoV-19/South_Africa/Tygerberg_1135/2021\|EPI_ISL_2695872\|2021-04-09 | 20H (Beta, V2) |
| hCoV-19/South_Africa/Tygerberg_1137/2021\|EPI_ISL_2695873\|2021-05-01 | 20H (Beta, V2) |
| hCoV-19/South_Africa/Tygerberg_1138/2021\|EPI_ISL_2695874\|2021-04-29 | 20I (Alpha, V1) |
| hCoV-19/South_Africa/Tygerberg_1141/2021\|EPI_ISL_2694594\|2021-06-04 | 21A (Delta) |
| hCoV-19/South_Africa/Tygerberg_1142/2021\|EPI_ISL_2694595\|2021-06-15 | 21A (Delta) |
| hCoV-19/South_Africa/Tygerberg_1143/2021\|EPI_ISL_2694596\|2021-06-16 | 21A (Delta) |
| hCoV-19/South_Africa/Tygerberg_1144/2021\|EPI_ISL_2694597\|2021-06-16 | 21A (Delta) |
| hCoV-19/South_Africa/Tygerberg_1145/2021\|EPI_ISL_2694598\|2021-06-16 | 20I (Alpha, V1) |
| hCoV-19/South_Africa/Tygerberg_1146/2021\|EPI_ISL_2694599\|2021-06-15 | 21A (Delta) |
| hCoV-19/South_Africa/Tygerberg_1147/2021\|EPI_ISL_2694600\|2021-06-16 | 21A (Delta) |
| hCoV-19/South_Africa/Tygerberg_1148/2021\|EPI_ISL_2694601\|2021-06-16 | 20H (Beta, V2) |
| hCoV-19/South_Africa/Tygerberg_1149/2021\|EPI_ISL_2694602\|2021-06-16 | 21A (Delta) |
| hCoV-19/South_Africa/Tygerberg_1150/2021\|EPI_ISL_2694603\|2021-06-09 | 20H (Beta, V2) |
| hCoV-19/South_Africa/Tygerberg_1151/2021\|EPI_ISL_2694604\|2021-06-08 | 20H (Beta, V2) |
| hCoV-19/South_Africa/Tygerberg_1153/2021\|EPI_ISL_2694605\|2021-06-17 | 21A (Delta) |
| hCoV-19/South_Africa/Tygerberg_1154/2021\|EPI_ISL_2694606\|2021-06-15 | 20I (Alpha, V1) |
| hCoV-19/South_Africa/Tygerberg_1155/2021\|EPI_ISL_2694607\|2021-06-16 | 20B |
| hCoV-19/South_Africa/Tygerberg_1156/2021\|EPI_ISL_2694608\|2021-06-16 | 20H (Beta, V2) |
| hCoV-19/South_Africa/Tygerberg_1157/2021\|EPI_ISL_2694609\|2021-06-15 | 21A (Delta) |
| hCoV-19/South_Africa/Tygerberg_1158/2021\|EPI_ISL_2694610\|2021-06-16 | 21A (Delta) |
| hCoV-19/South_Africa/Tygerberg_1159/2021\|EPI_ISL_2876306\|2021-06-23 | 21A (Delta) |
| hCoV-19/South_Africa/Tygerberg_1160/2021\|EPI_ISL_2876317\|2021-06-23 | 21A (Delta) |
| hCoV-19/South_Africa/Tygerberg_1161/2021\|EPI_ISL_2876329\|2021-06-23 | 21A (Delta) |
| hCoV-19/South_Africa/Tygerberg_1162/2021\|EPI_ISL_2876340\|2021-06-23 | 21A (Delta) |
| hCoV-19/South_Africa/Tygerberg_1163/2021\|EPI_ISL_2876354\|2021-06-23 | 21A (Delta) |
| hCoV-19/South_Africa/Tygerberg_1164/2021\|EPI_ISL_2876365\|2021-06-23 | 21A (Delta) |
| hCoV-19/South_Africa/Tygerberg_1165/2021\|EPI_ISL_2876376\|2021-06-23 | 21A (Delta) |
| hCoV-19/South_Africa/Tygerberg_1166/2021\|EPI_ISL_2876388\|2021-06-23 | 21A (Delta) |
| hCoV-19/South_Africa/Tygerberg_1167/2021\|EPI_ISL_2876307\|2021-06-23 | 21A (Delta) |

| | |
|---|---|
| hCoV-19/South_Africa/Tygerberg_1168/2021\|EPI_ISL_2876318\|2021-06-23 | 21A (Delta) |
| hCoV-19/South_Africa/Tygerberg_1169/2021\|EPI_ISL_2876330\|2021-06-23 | 21A (Delta) |
| hCoV-19/South_Africa/Tygerberg_1170/2021\|EPI_ISL_2876341\|2021-06-23 | 21A (Delta) |
| hCoV-19/South_Africa/Tygerberg_1171/2021\|EPI_ISL_2876355\|2021-06-23 | 21A (Delta) |
| hCoV-19/South_Africa/Tygerberg_1172/2021\|EPI_ISL_2876366\|2021-06-23 | 20H (Beta, V2) |
| hCoV-19/South_Africa/Tygerberg_1173/2021\|EPI_ISL_2876377\|2021-06-23 | 20A |
| hCoV-19/South_Africa/Tygerberg_1174/2021\|EPI_ISL_2876389\|2021-06-23 | 21A (Delta) |
| hCoV-19/South_Africa/Tygerberg_1175/2021\|EPI_ISL_2876308\|2021-06-24 | 21A (Delta) |
| hCoV-19/South_Africa/Tygerberg_1176/2021\|EPI_ISL_2876319\|2021-06-23 | 21A (Delta) |
| hCoV-19/South_Africa/Tygerberg_1177/2021\|EPI_ISL_2876331\|2021-06-24 | 21A (Delta) |
| hCoV-19/South_Africa/Tygerberg_1178/2021\|EPI_ISL_2876342\|2021-06-23 | 21A (Delta) |
| hCoV-19/South_Africa/Tygerberg_1179/2021\|EPI_ISL_2876356\|2021-06-24 | 21A (Delta) |
| hCoV-19/South_Africa/Tygerberg_1180/2021\|EPI_ISL_2876367\|2021-06-24 | 21A (Delta) |
| hCoV-19/South_Africa/Tygerberg_1181/2021\|EPI_ISL_2876378\|2021-06-24 | 21A (Delta) |
| hCoV-19/South_Africa/Tygerberg_1182/2021\|EPI_ISL_2876390\|2021-06-24 | 21A (Delta) |
| hCoV-19/South_Africa/Tygerberg_1183/2021\|EPI_ISL_2876309\|2021-06-24 | 21A (Delta) |
| hCoV-19/South_Africa/Tygerberg_1184/2021\|EPI_ISL_2876320\|2021-06-24 | 20A |
| hCoV-19/South_Africa/Tygerberg_1185/2021\|EPI_ISL_2876332\|2021-06-24 | 21A (Delta) |
| hCoV-19/South_Africa/Tygerberg_1186/2021\|EPI_ISL_2876343\|2021-06-24 | 21A (Delta) |
| hCoV-19/South_Africa/Tygerberg_1187/2021\|EPI_ISL_2876357\|2021-06-24 | 21A (Delta) |
| hCoV-19/South_Africa/Tygerberg_1188/2021\|EPI_ISL_2876368\|2021-06-24 | 21A (Delta) |
| hCoV-19/South_Africa/Tygerberg_1189/2021\|EPI_ISL_2876379\|2021-06-24 | 21A (Delta) |
| hCoV-19/South_Africa/Tygerberg_1190/2021\|EPI_ISL_2876391\|2021-06-24 | 21A (Delta) |
| hCoV-19/South_Africa/Tygerberg_1191/2021\|EPI_ISL_2876310\|2021-06-24 | 21A (Delta) |
| hCoV-19/South_Africa/Tygerberg_1192/2021\|EPI_ISL_2876321\|2021-06-24 | 21A (Delta) |
| hCoV-19/South_Africa/Tygerberg_1193/2021\|EPI_ISL_2876333\|2021-06-24 | 20H (Beta, V2) |
| hCoV-19/South_Africa/Tygerberg_1194/2021\|EPI_ISL_2876344\|2021-06-24 | 21A (Delta) |
| hCoV-19/South_Africa/Tygerberg_1195/2021\|EPI_ISL_2876358\|2021-06-24 | 21A (Delta) |
| hCoV-19/South_Africa/Tygerberg_1196/2021\|EPI_ISL_2876369\|2021-06-24 | 21A (Delta) |
| hCoV-19/South_Africa/Tygerberg_1197/2021\|EPI_ISL_2876380\|2021-06-24 | 21A (Delta) |
| hCoV-19/South_Africa/Tygerberg_1198/2021\|EPI_ISL_2876392\|2021-06-24 | 21A (Delta) |
| hCoV-19/South_Africa/Tygerberg_1199/2021\|EPI_ISL_2876311\|2021-06-24 | 21A (Delta) |
| hCoV-19/South_Africa/Tygerberg_1200/2021\|EPI_ISL_2876322\|2021-06-24 | 21A (Delta) |
| hCoV-19/South_Africa/Tygerberg_1201/2021\|EPI_ISL_2876334\|2021-06-24 | 20H (Beta, V2) |
| hCoV-19/South_Africa/Tygerberg_1202/2021\|EPI_ISL_2876345\|2021-06-24 | 21A (Delta) |
| hCoV-19/South_Africa/Tygerberg_1203/2021\|EPI_ISL_2876359\|2021-06-24 | 21A (Delta) |
| hCoV-19/South_Africa/Tygerberg_1204/2021\|EPI_ISL_2876370\|2021-06-24 | 21A (Delta) |
| hCoV-19/South_Africa/Tygerberg_1205/2021\|EPI_ISL_2876381\|2021-06-24 | 21A (Delta) |
| hCoV-19/South_Africa/Tygerberg_1206/2021\|EPI_ISL_2876312\|2021-06-24 | 21A (Delta) |
| hCoV-19/South_Africa/Tygerberg_1207/2021\|EPI_ISL_2876323\|2021-06-25 | 21A (Delta) |
| hCoV-19/South_Africa/Tygerberg_1208/2021\|EPI_ISL_2876335\|2021-06-25 | 21A (Delta) |
| hCoV-19/South_Africa/Tygerberg_1209/2021\|EPI_ISL_2876346\|2021-06-25 | 21A (Delta) |
| hCoV-19/South_Africa/Tygerberg_1210/2021\|EPI_ISL_2876360\|2021-06-24 | 21A (Delta) |
| hCoV-19/South_Africa/Tygerberg_1211/2021\|EPI_ISL_2876371\|2021-06-25 | 20H (Beta, V2) |
| hCoV-19/South_Africa/Tygerberg_1212/2021\|EPI_ISL_2876382\|2021-06-25 | 20H (Beta, V2) |
| hCoV-19/South_Africa/Tygerberg_1213/2021\|EPI_ISL_2876393\|2021-06-25 | 21A (Delta) |
| hCoV-19/South_Africa/Tygerberg_1215/2021\|EPI_ISL_2876324\|2021-06-25 | 21A (Delta) |
| hCoV-19/South_Africa/Tygerberg_1216/2021\|EPI_ISL_2876336\|2021-06-26 | 21A (Delta) |
| hCoV-19/South_Africa/Tygerberg_1217/2021\|EPI_ISL_2876347\|2021-06-25 | 21A (Delta) |
| hCoV-19/South_Africa/Tygerberg_1218/2021\|EPI_ISL_2876361\|2021-06-25 | 21A (Delta) |

| | |
|---|---|
| hCoV-19/South_Africa/Tygerberg_1219/2021\|EPI_ISL_2876372\|2021-06-25 | 21A (Delta) |
| hCoV-19/South_Africa/Tygerberg_1220/2021\|EPI_ISL_2876383\|2021-06-25 | 21A (Delta) |
| hCoV-19/South_Africa/Tygerberg_1221/2021\|EPI_ISL_2876394\|2021-06-25 | 21A (Delta) |
| hCoV-19/South_Africa/Tygerberg_1222/2021\|EPI_ISL_2876313\|2021-06-25 | 21A (Delta) |
| hCoV-19/South_Africa/Tygerberg_1223/2021\|EPI_ISL_2876325\|2021-06-25 | 21A (Delta) |
| hCoV-19/South_Africa/Tygerberg_1225/2021\|EPI_ISL_2876348\|2021-06-25 | 20H (Beta, V2) |
| hCoV-19/South_Africa/Tygerberg_1226/2021\|EPI_ISL_2876362\|2021-06-25 | 21A (Delta) |
| hCoV-19/South_Africa/Tygerberg_1227/2021\|EPI_ISL_2876373\|2021-06-25 | 21A (Delta) |
| hCoV-19/South_Africa/Tygerberg_1228/2021\|EPI_ISL_2876384\|2021-06-25 | 21A (Delta) |
| hCoV-19/South_Africa/Tygerberg_1229/2021\|EPI_ISL_2876395\|2021-06-25 | 21A (Delta) |
| hCoV-19/South_Africa/Tygerberg_1230/2021\|EPI_ISL_2876314\|2021-06-25 | 21A (Delta) |
| hCoV-19/South_Africa/Tygerberg_1231/2021\|EPI_ISL_2876326\|2021-06-25 | 21A (Delta) |
| hCoV-19/South_Africa/Tygerberg_1232/2021\|EPI_ISL_2876337\|2021-06-26 | 21A (Delta) |
| hCoV-19/South_Africa/Tygerberg_1234/2021\|EPI_ISL_2876385\|2021-06-21 | 20H (Beta, V2) |
| hCoV-19/South_Africa/Tygerberg_1235/2021\|EPI_ISL_2876396\|2021-06-27 | 21A (Delta) |
| hCoV-19/South_Africa/Tygerberg_1236/2021\|EPI_ISL_2876315\|2021-06-23 | 20H (Beta, V2) |
| hCoV-19/South_Africa/Tygerberg_1237/2021\|EPI_ISL_2876327\|2021-06-23 | 20H (Beta, V2) |
| hCoV-19/South_Africa/Tygerberg_1238/2021\|EPI_ISL_2876338\|2021-06-23 | 20H (Beta, V2) |
| hCoV-19/South_Africa/Tygerberg_1239/2021\|EPI_ISL_2876352\|2021-06-23 | 21A (Delta) |
| hCoV-19/South_Africa/Tygerberg_1240/2021\|EPI_ISL_2876363\|2021-06-23 | 21A (Delta) |
| hCoV-19/South_Africa/Tygerberg_1241/2021\|EPI_ISL_2876374\|2021-06-23 | 21A (Delta) |
| hCoV-19/South_Africa/Tygerberg_1242/2021\|EPI_ISL_2876386\|2021-06-23 | 20B |
| hCoV-19/South_Africa/Tygerberg_1243/2021\|EPI_ISL_2876397\|2021-06-23 | 21A (Delta) |
| hCoV-19/South_Africa/Tygerberg_1244/2021\|EPI_ISL_2876316\|2021-06-23 | 21A (Delta) |
| hCoV-19/South_Africa/Tygerberg_1245/2021\|EPI_ISL_2876328\|2021-06-23 | 21A (Delta) |
| hCoV-19/South_Africa/Tygerberg_1246/2021\|EPI_ISL_2876339\|2021-06-23 | 21A (Delta) |
| hCoV-19/South_Africa/Tygerberg_1247/2021\|EPI_ISL_2876353\|2021-06-23 | 21A (Delta) |
| hCoV-19/South_Africa/Tygerberg_1248/2021\|EPI_ISL_2876364\|2021-06-23 | 21A (Delta) |
| hCoV-19/South_Africa/Tygerberg_1249/2021\|EPI_ISL_2876375\|2021-06-23 | 21A (Delta) |
| hCoV-19/South_Africa/Tygerberg_1250/2021\|EPI_ISL_2876387\|2021-06-23 | 21A (Delta) |
| hCoV-19/South_Africa/Tygerberg_1251/2021\|EPI_ISL_2876398\|2021-06-23 | 21A (Delta) |
| hCoV-19/South_Africa/Tygerberg_1252/2021\|EPI_ISL_3032461\|2021-06-30 | 21A (Delta) |
| hCoV-19/South_Africa/Tygerberg_1253/2021\|EPI_ISL_3032462\|2021-06-28 | 21A (Delta) |
| hCoV-19/South_Africa/Tygerberg_1254/2021\|EPI_ISL_3032463\|2021-06-30 | 21A (Delta) |
| hCoV-19/South_Africa/Tygerberg_1255/2021\|EPI_ISL_3032464\|2021-06-28 | 21A (Delta) |
| hCoV-19/South_Africa/Tygerberg_1256/2021\|EPI_ISL_3032465\|2021-06-29 | 21A (Delta) |
| hCoV-19/South_Africa/Tygerberg_1257/2021\|EPI_ISL_3032466\|2021-06-28 | 20H (Beta, V2) |
| hCoV-19/South_Africa/Tygerberg_1258/2021\|EPI_ISL_3032467\|2021-06-29 | 21A (Delta) |
| hCoV-19/South_Africa/Tygerberg_1259/2021\|EPI_ISL_3032468\|2021-06-28 | 21A (Delta) |
| hCoV-19/South_Africa/Tygerberg_1260/2021\|EPI_ISL_3032469\|2021-06-28 | 21A (Delta) |
| hCoV-19/South_Africa/Tygerberg_1261/2021\|EPI_ISL_3032470\|2021-06-30 | 21A (Delta) |
| hCoV-19/South_Africa/Tygerberg_1262/2021\|EPI_ISL_3032471\|2021-06-27 | 21A (Delta) |
| hCoV-19/South_Africa/Tygerberg_1263/2021\|EPI_ISL_3032472\|2021-06-28 | 21A (Delta) |
| hCoV-19/South_Africa/Tygerberg_1264/2021\|EPI_ISL_3032473\|2021-06-26 | 21A (Delta) |
| hCoV-19/South_Africa/Tygerberg_1265/2021\|EPI_ISL_3032474\|2021-06-29 | 21A (Delta) |
| hCoV-19/South_Africa/Tygerberg_1266/2021\|EPI_ISL_3032475\|2021-06-30 | 20H (Beta, V2) |
| hCoV-19/South_Africa/Tygerberg_1267/2021\|EPI_ISL_3032476\|2021-06-30 | 21A (Delta) |
| hCoV-19/South_Africa/Tygerberg_1268/2021\|EPI_ISL_3032477\|2021-06-30 | 21A (Delta) |
| hCoV-19/South_Africa/Tygerberg_1269/2021\|EPI_ISL_3032478\|2021-06-30 | 21A (Delta) |
| hCoV-19/South_Africa/Tygerberg_1270/2021\|EPI_ISL_3032479\|2021-07-07 | 21A (Delta) |

| | |
|---|---|
| hCoV-19/South_Africa/Tygerberg_1271/2021\|EPI_ISL_3032480\|2021-07-06 | 21A (Delta) |
| hCoV-19/South_Africa/Tygerberg_1272/2021\|EPI_ISL_3032481\|2021-07-07 | 21A (Delta) |
| hCoV-19/South_Africa/Tygerberg_1273/2021\|EPI_ISL_3032482\|2021-07-07 | 21A (Delta) |
| hCoV-19/South_Africa/Tygerberg_1274/2021\|EPI_ISL_3032483\|2021-07-07 | 21A (Delta) |
| hCoV-19/South_Africa/Tygerberg_1276/2021\|EPI_ISL_3032485\|2021-07-06 | 21A (Delta) |
| hCoV-19/South_Africa/Tygerberg_1277/2021\|EPI_ISL_3032486\|2021-07-07 | 21A (Delta) |
| hCoV-19/South_Africa/Tygerberg_1278/2021\|EPI_ISL_3032487\|2021-07-07 | 21A (Delta) |
| hCoV-19/South_Africa/Tygerberg_1279/2021\|EPI_ISL_3032488\|2021-07-01 | 21A (Delta) |
| hCoV-19/South_Africa/Tygerberg_1280/2021\|EPI_ISL_3032489\|2021-07-02 | 21A (Delta) |
| hCoV-19/South_Africa/Tygerberg_1281/2021\|EPI_ISL_3032490\|2021-07-02 | 21A (Delta) |
| hCoV-19/South_Africa/Tygerberg_1282/2021\|EPI_ISL_3032491\|2021-07-06 | 21A (Delta) |
| hCoV-19/South_Africa/Tygerberg_1283/2021\|EPI_ISL_3032492\|2021-07-06 | 21A (Delta) |
| hCoV-19/South_Africa/Tygerberg_1284/2021\|EPI_ISL_3032493\|2021-07-06 | 21A (Delta) |
| hCoV-19/South_Africa/Tygerberg_1285/2021\|EPI_ISL_3032494\|2021-07-05 | 21A (Delta) |
| hCoV-19/South_Africa/Tygerberg_1286/2021\|EPI_ISL_3032495\|2021-07-06 | 21A (Delta) |
| hCoV-19/South_Africa/Tygerberg_1287/2021\|EPI_ISL_3032496\|2021-07-06 | 21A (Delta) |
| hCoV-19/South_Africa/Tygerberg_1288/2021\|EPI_ISL_3032497\|2021-07-06 | 21A (Delta) |
| hCoV-19/South_Africa/Tygerberg_1289/2021\|EPI_ISL_3032498\|2021-07-06 | 21A (Delta) |
| hCoV-19/South_Africa/Tygerberg_1290/2021\|EPI_ISL_3032499\|2021-07-06 | 20H (Beta, V2) |
| hCoV-19/South_Africa/Tygerberg_1291/2021\|EPI_ISL_3032500\|2021-07-06 | 21A (Delta) |
| hCoV-19/South_Africa/Tygerberg_1292/2021\|EPI_ISL_3032501\|2021-07-06 | 21A (Delta) |
| hCoV-19/South_Africa/Tygerberg_1293/2021\|EPI_ISL_3032502\|2021-07-06 | 21A (Delta) |
| hCoV-19/South_Africa/Tygerberg_1294/2021\|EPI_ISL_3032503\|2021-07-05 | 21A (Delta) |
| hCoV-19/South_Africa/Tygerberg_1295/2021\|EPI_ISL_3032504\|2021-07-06 | 21A (Delta) |
| hCoV-19/South_Africa/Tygerberg_1296/2021\|EPI_ISL_3032505\|2021-06-17 | 20H (Beta, V2) |
| hCoV-19/South_Africa/Tygerberg_1297/2021\|EPI_ISL_3032506\|2021-07-07 | 21A (Delta) |
| hCoV-19/South_Africa/Tygerberg_1298/2021\|EPI_ISL_3032507\|2021-07-09 | 21A (Delta) |
| hCoV-19/South_Africa/Tygerberg_1299/2021\|EPI_ISL_3032508\|2021-07-09 | 21A (Delta) |
| hCoV-19/South_Africa/Tygerberg_1300/2021\|EPI_ISL_3032509\|2021-07-09 | 21A (Delta) |
| hCoV-19/South_Africa/Tygerberg_1301/2021\|EPI_ISL_3032510\|2021-07-09 | 21A (Delta) |
| hCoV-19/South_Africa/Tygerberg_1302/2021\|EPI_ISL_3032511\|2021-07-08 | 21A (Delta) |
| hCoV-19/South_Africa/Tygerberg_1303/2021\|EPI_ISL_3032512\|2021-07-09 | 20B |
| hCoV-19/South_Africa/Tygerberg_1304/2021\|EPI_ISL_3032513\|2021-07-08 | 21A (Delta) |
| hCoV-19/South_Africa/Tygerberg_1305/2021\|EPI_ISL_3032514\|2021-07-09 | 21A (Delta) |
| hCoV-19/South_Africa/Tygerberg_1306/2021\|EPI_ISL_3032515\|2021-07-08 | 21A (Delta) |
| hCoV-19/South_Africa/Tygerberg_1307/2021\|EPI_ISL_3032516\|2021-07-09 | 21A (Delta) |
| hCoV-19/South_Africa/Tygerberg_1308/2021\|EPI_ISL_3032517\|2021-07-08 | 21A (Delta) |
| hCoV-19/South_Africa/Tygerberg_1310/2021\|EPI_ISL_3032519\|2021-07-09 | 21A (Delta) |
| hCoV-19/South_Africa/Tygerberg_1311/2021\|EPI_ISL_3032520\|2021-07-07 | 21A (Delta) |
| hCoV-19/South_Africa/Tygerberg_1312/2021\|EPI_ISL_3032521\|2021-07-07 | 21A (Delta) |
| hCoV-19/South_Africa/Tygerberg_1314/2021\|EPI_ISL_3032522\|2021-07-03 | 21A (Delta) |
| hCoV-19/South_Africa/Tygerberg_1316/2021\|EPI_ISL_3032523\|2021-07-06 | 20H (Beta, V2) |
| hCoV-19/South_Africa/Tygerberg_1317/2021\|EPI_ISL_3032524\|2021-07-08 | 20H (Beta, V2) |
| hCoV-19/South_Africa/Tygerberg_1318/2021\|EPI_ISL_3032525\|2021-07-06 | 21A (Delta) |
| hCoV-19/South_Africa/Tygerberg_1319/2021\|EPI_ISL_3032526\|2021-07-06 | 21A (Delta) |
| hCoV-19/South_Africa/Tygerberg_1320/2021\|EPI_ISL_3032527\|2021-07-06 | 21A (Delta) |
| hCoV-19/South_Africa/Tygerberg_1322/2021\|EPI_ISL_3032528\|2021-06-22 | 20H (Beta, V2) |
| hCoV-19/South_Africa/Tygerberg_1323/2021\|EPI_ISL_3032529\|2021-06-11 | 20H (Beta, V2) |
| hCoV-19/South_Africa/Tygerberg_1325/2021\|EPI_ISL_3032531\|2021-06-11 | 21A (Delta) |
| hCoV-19/South_Africa/Tygerberg_1326/2021\|EPI_ISL_3032532\|2021-06-23 | 20H (Beta, V2) |

| | |
|---|---|
| hCoV-19/South_Africa/Tygerberg_1327/2021\|EPI_ISL_3032533\|2021-06-23 | 21A (Delta) |
| hCoV-19/South_Africa/Tygerberg_1328/2021\|EPI_ISL_3032534\|2021-06-23 | 21A (Delta) |
| hCoV-19/South_Africa/Tygerberg_1329/2021\|EPI_ISL_3032535\|2021-06-23 | 21A (Delta) |
| hCoV-19/South_Africa/Tygerberg_1330/2021\|EPI_ISL_3032536\|2021-06-23 | 21A (Delta) |
| hCoV-19/South_Africa/Tygerberg_1331/2021\|EPI_ISL_3032537\|2021-06-17 | 21A (Delta) |
| hCoV-19/South_Africa/Tygerberg_1338/2021\|EPI_ISL_3118698\|2021-07-11 | 21A (Delta) |
| hCoV-19/South_Africa/Tygerberg_1339/2021\|EPI_ISL_3118709\|2021-07-11 | 21A (Delta) |
| hCoV-19/South_Africa/Tygerberg_1340/2021\|EPI_ISL_3118720\|2021-07-11 | 20H (Beta, V2) |
| hCoV-19/South_Africa/Tygerberg_1341/2021\|EPI_ISL_3118731\|2021-07-11 | 21A (Delta) |
| hCoV-19/South_Africa/Tygerberg_1342/2021\|EPI_ISL_3118742\|2021-07-11 | 21A (Delta) |
| hCoV-19/South_Africa/Tygerberg_1343/2021\|EPI_ISL_3118753\|2021-07-11 | 21A (Delta) |
| hCoV-19/South_Africa/Tygerberg_1344/2021\|EPI_ISL_3118763\|2021-07-11 | 21A (Delta) |
| hCoV-19/South_Africa/Tygerberg_1345/2021\|EPI_ISL_3118774\|2021-07-10 | 21A (Delta) |
| hCoV-19/South_Africa/Tygerberg_1346/2021\|EPI_ISL_3118699\|2021-07-11 | 21A (Delta) |
| hCoV-19/South_Africa/Tygerberg_1347/2021\|EPI_ISL_3118710\|2021-07-11 | 21A (Delta) |
| hCoV-19/South_Africa/Tygerberg_1348/2021\|EPI_ISL_3118721\|2021-07-11 | 21A (Delta) |
| hCoV-19/South_Africa/Tygerberg_1349/2021\|EPI_ISL_3118732\|2021-07-11 | 21A (Delta) |
| hCoV-19/South_Africa/Tygerberg_1350/2021\|EPI_ISL_3118743\|2021-07-10 | 21A (Delta) |
| hCoV-19/South_Africa/Tygerberg_1351/2021\|EPI_ISL_3118754\|2021-07-11 | 21A (Delta) |
| hCoV-19/South_Africa/Tygerberg_1352/2021\|EPI_ISL_3118764\|2021-07-11 | 21A (Delta) |
| hCoV-19/South_Africa/Tygerberg_1353/2021\|EPI_ISL_3118775\|2021-07-11 | 21A (Delta) |
| hCoV-19/South_Africa/Tygerberg_1354/2021\|EPI_ISL_3118700\|2021-07-11 | 21A (Delta) |
| hCoV-19/South_Africa/Tygerberg_1355/2021\|EPI_ISL_3118711\|2021-07-11 | 21A (Delta) |
| hCoV-19/South_Africa/Tygerberg_1356/2021\|EPI_ISL_3118722\|2021-07-12 | 21A (Delta) |
| hCoV-19/South_Africa/Tygerberg_1357/2021\|EPI_ISL_3118733\|2021-07-12 | 21A (Delta) |
| hCoV-19/South_Africa/Tygerberg_1358/2021\|EPI_ISL_3118744\|2021-07-12 | 21A (Delta) |
| hCoV-19/South_Africa/Tygerberg_1361/2021\|EPI_ISL_3118776\|2021-07-11 | 21A (Delta) |
| hCoV-19/South_Africa/Tygerberg_1362/2021\|EPI_ISL_3118701\|2021-07-12 | 21A (Delta) |
| hCoV-19/South_Africa/Tygerberg_1363/2021\|EPI_ISL_3118712\|2021-07-12 | 21A (Delta) |
| hCoV-19/South_Africa/Tygerberg_1364/2021\|EPI_ISL_3118723\|2021-07-12 | 21A (Delta) |
| hCoV-19/South_Africa/Tygerberg_1365/2021\|EPI_ISL_3118734\|2021-07-12 | 21A (Delta) |
| hCoV-19/South_Africa/Tygerberg_1366/2021\|EPI_ISL_3118745\|2021-07-12 | 21A (Delta) |
| hCoV-19/South_Africa/Tygerberg_1367/2021\|EPI_ISL_3118755\|2021-07-12 | 21A (Delta) |
| hCoV-19/South_Africa/Tygerberg_1368/2021\|EPI_ISL_3118766\|2021-07-12 | 21A (Delta) |
| hCoV-19/South_Africa/Tygerberg_1369/2021\|EPI_ISL_3118777\|2021-07-13 | 21A (Delta) |
| hCoV-19/South_Africa/Tygerberg_1370/2021\|EPI_ISL_3118702\|2021-07-12 | 21A (Delta) |
| hCoV-19/South_Africa/Tygerberg_1371/2021\|EPI_ISL_3118713\|2021-07-12 | 21A (Delta) |
| hCoV-19/South_Africa/Tygerberg_1372/2021\|EPI_ISL_3118724\|2021-07-12 | 21A (Delta) |
| hCoV-19/South_Africa/Tygerberg_1373/2021\|EPI_ISL_3118735\|2021-07-12 | 21A (Delta) |
| hCoV-19/South_Africa/Tygerberg_1374/2021\|EPI_ISL_3118746\|2021-07-12 | 21A (Delta) |
| hCoV-19/South_Africa/Tygerberg_1375/2021\|EPI_ISL_3118756\|2021-07-12 | 21A (Delta) |
| hCoV-19/South_Africa/Tygerberg_1376/2021\|EPI_ISL_3118767\|2021-07-11 | 21A (Delta) |
| hCoV-19/South_Africa/Tygerberg_1378/2021\|EPI_ISL_3118703\|2021-07-13 | 21A (Delta) |
| hCoV-19/South_Africa/Tygerberg_1379/2021\|EPI_ISL_3118714\|2021-07-13 | 21A (Delta) |
| hCoV-19/South_Africa/Tygerberg_1380/2021\|EPI_ISL_3118725\|2021-07-13 | 21A (Delta) |
| hCoV-19/South_Africa/Tygerberg_1381/2021\|EPI_ISL_3118736\|2021-07-13 | 21A (Delta) |
| hCoV-19/South_Africa/Tygerberg_1382/2021\|EPI_ISL_3118747\|2021-07-13 | 21A (Delta) |
| hCoV-19/South_Africa/Tygerberg_1383/2021\|EPI_ISL_3118757\|2021-07-13 | 21A (Delta) |
| hCoV-19/South_Africa/Tygerberg_1384/2021\|EPI_ISL_3118768\|2021-07-13 | 21A (Delta) |
| hCoV-19/South_Africa/Tygerberg_1385/2021\|EPI_ISL_3118779\|2021-07-13 | 21A (Delta) |

| | |
|---|---|
| hCoV-19/South_Africa/Tygerberg_1386/2021\|EPI_ISL_3118704\|2021-07-12 | 21A (Delta) |
| hCoV-19/South_Africa/Tygerberg_1387/2021\|EPI_ISL_3118715\|2021-07-12 | 21A (Delta) |
| hCoV-19/South_Africa/Tygerberg_1389/2021\|EPI_ISL_3118737\|2021-07-12 | 21A (Delta) |
| hCoV-19/South_Africa/Tygerberg_1390/2021\|EPI_ISL_3118748\|2021-07-12 | 21A (Delta) |
| hCoV-19/South_Africa/Tygerberg_1391/2021\|EPI_ISL_3118758\|2021-07-12 | 21A (Delta) |
| hCoV-19/South_Africa/Tygerberg_1392/2021\|EPI_ISL_3118769\|2021-07-12 | 21A (Delta) |
| hCoV-19/South_Africa/Tygerberg_1393/2021\|EPI_ISL_3118780\|2021-07-12 | 21A (Delta) |
| hCoV-19/South_Africa/Tygerberg_1394/2021\|EPI_ISL_3118705\|2021-07-12 | 21A (Delta) |
| hCoV-19/South_Africa/Tygerberg_1395/2021\|EPI_ISL_3118716\|2021-07-13 | 21A (Delta) |
| hCoV-19/South_Africa/Tygerberg_1396/2021\|EPI_ISL_3118727\|2021-07-13 | 21A (Delta) |
| hCoV-19/South_Africa/Tygerberg_1397/2021\|EPI_ISL_3118738\|2021-07-13 | 21A (Delta) |
| hCoV-19/South_Africa/Tygerberg_1398/2021\|EPI_ISL_3118749\|2021-07-13 | 21A (Delta) |
| hCoV-19/South_Africa/Tygerberg_1399/2021\|EPI_ISL_3118759\|2021-07-13 | 21A (Delta) |
| hCoV-19/South_Africa/Tygerberg_1400/2021\|EPI_ISL_3118770\|2021-07-13 | 21A (Delta) |
| hCoV-19/South_Africa/Tygerberg_1401/2021\|EPI_ISL_3118781\|2021-07-14 | 21A (Delta) |
| hCoV-19/South_Africa/Tygerberg_1402/2021\|EPI_ISL_3118706\|2021-07-14 | 21A (Delta) |
| hCoV-19/South_Africa/Tygerberg_1403/2021\|EPI_ISL_3118717\|2021-07-14 | 21A (Delta) |
| hCoV-19/South_Africa/Tygerberg_1404/2021\|EPI_ISL_3118728\|2021-07-14 | 21A (Delta) |
| hCoV-19/South_Africa/Tygerberg_1405/2021\|EPI_ISL_3118739\|2021-07-14 | 21A (Delta) |
| hCoV-19/South_Africa/Tygerberg_1406/2021\|EPI_ISL_3118750\|2021-07-14 | 21A (Delta) |
| hCoV-19/South_Africa/Tygerberg_1407/2021\|EPI_ISL_3118760\|2021-07-13 | 21A (Delta) |
| hCoV-19/South_Africa/Tygerberg_1408/2021\|EPI_ISL_3118771\|2021-07-14 | 21A (Delta) |
| hCoV-19/South_Africa/Tygerberg_1409/2021\|EPI_ISL_3118782\|2021-07-14 | 21A (Delta) |
| hCoV-19/South_Africa/Tygerberg_1410/2021\|EPI_ISL_3118707\|2021-07-14 | 21A (Delta) |
| hCoV-19/South_Africa/Tygerberg_1411/2021\|EPI_ISL_3118718\|2021-07-14 | 21A (Delta) |
| hCoV-19/South_Africa/Tygerberg_1412/2021\|EPI_ISL_3118729\|2021-07-13 | 21A (Delta) |
| hCoV-19/South_Africa/Tygerberg_1413/2021\|EPI_ISL_3118740\|2021-07-13 | 21A (Delta) |
| hCoV-19/South_Africa/Tygerberg_1414/2021\|EPI_ISL_3118751\|2021-07-14 | 21A (Delta) |
| hCoV-19/South_Africa/Tygerberg_1415/2021\|EPI_ISL_3118761\|2021-07-14 | 21A (Delta) |
| hCoV-19/South_Africa/Tygerberg_1416/2021\|EPI_ISL_3118772\|2021-07-14 | 21A (Delta) |
| hCoV-19/South_Africa/Tygerberg_1417/2021\|EPI_ISL_3118783\|2021-07-14 | 21A (Delta) |
| hCoV-19/South_Africa/Tygerberg_1418/2021\|EPI_ISL_3118708\|2021-07-14 | 21A (Delta) |
| hCoV-19/South_Africa/Tygerberg_1420/2021\|EPI_ISL_3118730\|2021-07-14 | 21A (Delta) |
| hCoV-19/South_Africa/Tygerberg_1421/2021\|EPI_ISL_3118741\|2021-07-14 | 21A (Delta) |
| hCoV-19/South_Africa/Tygerberg_1422/2021\|EPI_ISL_3118752\|2021-07-14 | 20H (Beta, V2) |
| hCoV-19/South_Africa/Tygerberg_1423/2021\|EPI_ISL_3118762\|2021-07-14 | 21A (Delta) |
| hCoV-19/South_Africa/Tygerberg_1428/2021\|EPI_ISL_3247084\|2021-06-21 | 21A (Delta) |
| hCoV-19/South_Africa/Tygerberg_1430/2021\|EPI_ISL_3247106\|2021-06-22 | 21A (Delta) |
| hCoV-19/South_Africa/Tygerberg_1431/2021\|EPI_ISL_3247116\|2021-06-23 | 20H (Beta, V2) |
| hCoV-19/South_Africa/Tygerberg_1437/2021\|EPI_ISL_3247095\|2021-06-26 | 20B |
| hCoV-19/South_Africa/Tygerberg_1438/2021\|EPI_ISL_3247107\|2021-06-26 | 21A (Delta) |
| hCoV-19/South_Africa/Tygerberg_1439/2021\|EPI_ISL_3247117\|2021-07-19 | 21A (Delta) |
| hCoV-19/South_Africa/Tygerberg_1440/2021\|EPI_ISL_3247126\|2021-07-19 | 21A (Delta) |
| hCoV-19/South_Africa/Tygerberg_1441/2021\|EPI_ISL_3247135\|2021-07-19 | 21A (Delta) |
| hCoV-19/South_Africa/Tygerberg_1444/2021\|EPI_ISL_3247086\|2021-07-19 | 21A (Delta) |
| hCoV-19/South_Africa/Tygerberg_1445/2021\|EPI_ISL_3247096\|2021-07-19 | 21A (Delta) |
| hCoV-19/South_Africa/Tygerberg_1446/2021\|EPI_ISL_3247108\|2021-07-19 | 21A (Delta) |
| hCoV-19/South_Africa/Tygerberg_1448/2021\|EPI_ISL_3247127\|2021-07-19 | 21A (Delta) |
| hCoV-19/South_Africa/Tygerberg_1449/2021\|EPI_ISL_3247136\|2021-07-19 | 21A (Delta) |
| hCoV-19/South_Africa/Tygerberg_1450/2021\|EPI_ISL_3247075\|2021-07-19 | 21A (Delta) |

| | |
|---|---|
| hCoV-19/South_Africa/Tygerberg_1451/2021\|EPI_ISL_3247087\|2021-07-19 | 21A (Delta) |
| hCoV-19/South_Africa/Tygerberg_1452/2021\|EPI_ISL_3247097\|2021-07-20 | 21A (Delta) |
| hCoV-19/South_Africa/Tygerberg_1455/2021\|EPI_ISL_3247128\|2021-07-19 | 21A (Delta) |
| hCoV-19/South_Africa/Tygerberg_1457/2021\|EPI_ISL_3247146\|2021-07-19 | 21A (Delta) |
| hCoV-19/South_Africa/Tygerberg_1458/2021\|EPI_ISL_3247076\|2021-07-19 | 21A (Delta) |
| hCoV-19/South_Africa/Tygerberg_1459/2021\|EPI_ISL_3247088\|2021-07-19 | 21A (Delta) |
| hCoV-19/South_Africa/Tygerberg_1460/2021\|EPI_ISL_3247098\|2021-07-19 | 21A (Delta) |
| hCoV-19/South_Africa/Tygerberg_1461/2021\|EPI_ISL_3247109\|2021-07-19 | 21A (Delta) |
| hCoV-19/South_Africa/Tygerberg_1462/2021\|EPI_ISL_3247119\|2021-07-19 | 21A (Delta) |
| hCoV-19/South_Africa/Tygerberg_1463/2021\|EPI_ISL_3247129\|2021-07-19 | 21A (Delta) |
| hCoV-19/South_Africa/Tygerberg_1464/2021\|EPI_ISL_3247138\|2021-07-19 | 21A (Delta) |
| hCoV-19/South_Africa/Tygerberg_1465/2021\|EPI_ISL_3247147\|2021-07-19 | 21A (Delta) |
| hCoV-19/South_Africa/Tygerberg_1466/2021\|EPI_ISL_3247077\|2021-07-19 | 21A (Delta) |
| hCoV-19/South_Africa/Tygerberg_1467/2021\|EPI_ISL_3247089\|2021-07-19 | 21A (Delta) |
| hCoV-19/South_Africa/Tygerberg_1468/2021\|EPI_ISL_3247099\|2021-07-19 | 21A (Delta) |
| hCoV-19/South_Africa/Tygerberg_1469/2021\|EPI_ISL_3247110\|2021-07-19 | 21A (Delta) |
| hCoV-19/South_Africa/Tygerberg_1472/2021\|EPI_ISL_3247139\|2021-07-19 | 21A (Delta) |
| hCoV-19/South_Africa/Tygerberg_1473/2021\|EPI_ISL_3247148\|2021-07-19 | 21A (Delta) |
| hCoV-19/South_Africa/Tygerberg_1474/2021\|EPI_ISL_3247078\|2021-07-19 | 21A (Delta) |
| hCoV-19/South_Africa/Tygerberg_1475/2021\|EPI_ISL_3247090\|2021-07-19 | 21A (Delta) |
| hCoV-19/South_Africa/Tygerberg_1476/2021\|EPI_ISL_3247100\|2021-07-19 | 21A (Delta) |
| hCoV-19/South_Africa/Tygerberg_1477/2021\|EPI_ISL_3247111\|2021-07-19 | 21A (Delta) |
| hCoV-19/South_Africa/Tygerberg_1478/2021\|EPI_ISL_3247121\|2021-07-19 | 21A (Delta) |
| hCoV-19/South_Africa/Tygerberg_1479/2021\|EPI_ISL_3247130\|2021-07-19 | 20H (Beta, V2) |
| hCoV-19/South_Africa/Tygerberg_1480/2021\|EPI_ISL_3247140\|2021-07-19 | 21A (Delta) |
| hCoV-19/South_Africa/Tygerberg_1481/2021\|EPI_ISL_3247149\|2021-07-19 | 21A (Delta) |
| hCoV-19/South_Africa/Tygerberg_1482/2021\|EPI_ISL_3247079\|2021-07-19 | 21A (Delta) |
| hCoV-19/South_Africa/Tygerberg_1483/2021\|EPI_ISL_3247091\|2021-07-19 | 21A (Delta) |
| hCoV-19/South_Africa/Tygerberg_1484/2021\|EPI_ISL_3247101\|2021-07-19 | 21A (Delta) |
| hCoV-19/South_Africa/Tygerberg_1485/2021\|EPI_ISL_3247112\|2021-07-19 | 21A (Delta) |
| hCoV-19/South_Africa/Tygerberg_1487/2021\|EPI_ISL_3247131\|2021-07-19 | 21A (Delta) |
| hCoV-19/South_Africa/Tygerberg_1488/2021\|EPI_ISL_3247141\|2021-07-19 | 21A (Delta) |
| hCoV-19/South_Africa/Tygerberg_1489/2021\|EPI_ISL_3247150\|2021-07-19 | 21A (Delta) |
| hCoV-19/South_Africa/Tygerberg_1490/2021\|EPI_ISL_3247080\|2021-07-19 | 21A (Delta) |
| hCoV-19/South_Africa/Tygerberg_1491/2021\|EPI_ISL_3247092\|2021-07-19 | 21A (Delta) |
| hCoV-19/South_Africa/Tygerberg_1492/2021\|EPI_ISL_3247102\|2021-07-19 | 21A (Delta) |
| hCoV-19/South_Africa/Tygerberg_1493/2021\|EPI_ISL_3247113\|2021-07-19 | 21A (Delta) |
| hCoV-19/South_Africa/Tygerberg_1494/2021\|EPI_ISL_3247123\|2021-07-19 | 21A (Delta) |
| hCoV-19/South_Africa/Tygerberg_1495/2021\|EPI_ISL_3247132\|2021-07-19 | 21A (Delta) |
| hCoV-19/South_Africa/Tygerberg_1496/2021\|EPI_ISL_3247142\|2021-07-19 | 21A (Delta) |
| hCoV-19/South_Africa/Tygerberg_1497/2021\|EPI_ISL_3247151\|2021-07-19 | 21A (Delta) |
| hCoV-19/South_Africa/Tygerberg_1498/2021\|EPI_ISL_3247081\|2021-07-19 | 21A (Delta) |
| hCoV-19/South_Africa/Tygerberg_1499/2021\|EPI_ISL_3247093\|2021-07-19 | 21A (Delta) |
| hCoV-19/South_Africa/Tygerberg_1501/2021\|EPI_ISL_3247114\|2021-07-19 | 21A (Delta) |
| hCoV-19/South_Africa/Tygerberg_1502/2021\|EPI_ISL_3247124\|2021-07-19 | 21A (Delta) |
| hCoV-19/South_Africa/Tygerberg_1503/2021\|EPI_ISL_3247133\|2021-07-19 | 21A (Delta) |
| hCoV-19/South_Africa/Tygerberg_1505/2021\|EPI_ISL_3247152\|2021-07-19 | 21A (Delta) |
| hCoV-19/South_Africa/Tygerberg_1506/2021\|EPI_ISL_3247082\|2021-07-19 | 21A (Delta) |
| hCoV-19/South_Africa/Tygerberg_1508/2021\|EPI_ISL_3247104\|2021-07-19 | 21A (Delta) |
| hCoV-19/South_Africa/Tygerberg_1509/2021\|EPI_ISL_3247115\|2021-07-19 | 21A (Delta) |

| | |
|---|---|
| hCoV-19/South_Africa/Tygerberg_1510/2021\|EPI_ISL_3247125\|2021-07-19 | 21A (Delta) |
| hCoV-19/South_Africa/Tygerberg_1511/2021\|EPI_ISL_3247134\|2021-07-19 | 21A (Delta) |
| hCoV-19/South_Africa/Tygerberg_1512/2021\|EPI_ISL_3247144\|2021-07-20 | 21A (Delta) |
| hCoV-19/South_Africa/Tygerberg_1514/2021\|EPI_ISL_3247083\|2021-07-20 | 21A (Delta) |
| hCoV-19/South_Africa/Tygerberg_1515/2021\|EPI_ISL_3247094\|2021-07-19 | 21A (Delta) |
| hCoV-19/South_Africa/Tygerberg_1516/2021\|EPI_ISL_3247105\|2021-07-19 | 21A (Delta) |
| hCoV-19/South_Africa/Tygerberg_1517/2021\|EPI_ISL_3482512\|2021-07-22 | 21A (Delta) |
| hCoV-19/South_Africa/Tygerberg_1519/2021\|EPI_ISL_3482516\|2021-07-22 | 21A (Delta) |
| hCoV-19/South_Africa/Tygerberg_1520/2021\|EPI_ISL_3482518\|2021-07-22 | 21A (Delta) |
| hCoV-19/South_Africa/Tygerberg_1522/2021\|EPI_ISL_3482521\|2021-07-22 | 21A (Delta) |
| hCoV-19/South_Africa/Tygerberg_1524/2021\|EPI_ISL_3482523\|2021-07-23 | 21A (Delta) |
| hCoV-19/South_Africa/Tygerberg_1525/2021\|EPI_ISL_3482524\|2021-07-23 | 21A (Delta) |
| hCoV-19/South_Africa/Tygerberg_1526/2021\|EPI_ISL_3482526\|2021-07-23 | 21A (Delta) |
| hCoV-19/South_Africa/Tygerberg_1527/2021\|EPI_ISL_3482528\|2021-07-23 | 21A (Delta) |
| hCoV-19/South_Africa/Tygerberg_1528/2021\|EPI_ISL_3482529\|2021-07-23 | 21A (Delta) |
| hCoV-19/South_Africa/Tygerberg_1529/2021\|EPI_ISL_3482531\|2021-07-22 | 21A (Delta) |
| hCoV-19/South_Africa/Tygerberg_1530/2021\|EPI_ISL_3482533\|2021-07-23 | 21A (Delta) |
| hCoV-19/South_Africa/Tygerberg_1531/2021\|EPI_ISL_3482534\|2021-07-23 | 21A (Delta) |
| hCoV-19/South_Africa/Tygerberg_1533/2021\|EPI_ISL_3482536\|2021-07-23 | 20D |
| hCoV-19/South_Africa/Tygerberg_1534/2021\|EPI_ISL_3482538\|2021-07-23 | 21A (Delta) |
| hCoV-19/South_Africa/Tygerberg_1535/2021\|EPI_ISL_3482539\|2021-07-23 | 21A (Delta) |
| hCoV-19/South_Africa/Tygerberg_1536/2021\|EPI_ISL_3482541\|2021-07-23 | 21A (Delta) |
| hCoV-19/South_Africa/Tygerberg_1537/2021\|EPI_ISL_3482542\|2021-07-23 | 21A (Delta) |
| hCoV-19/South_Africa/Tygerberg_1538/2021\|EPI_ISL_3482544\|2021-07-24 | 21A (Delta) |
| hCoV-19/South_Africa/Tygerberg_1539/2021\|EPI_ISL_3482546\|2021-07-24 | 21A (Delta) |
| hCoV-19/South_Africa/Tygerberg_1540/2021\|EPI_ISL_3482547\|2021-07-24 | 21A (Delta) |
| hCoV-19/South_Africa/Tygerberg_1541/2021\|EPI_ISL_3482549\|2021-07-25 | 21A (Delta) |
| hCoV-19/South_Africa/Tygerberg_1542/2021\|EPI_ISL_3482551\|2021-07-26 | 21A (Delta) |
| hCoV-19/South_Africa/Tygerberg_1543/2021\|EPI_ISL_3482552\|2021-07-26 | 21A (Delta) |
| hCoV-19/South_Africa/Tygerberg_1544/2021\|EPI_ISL_3482554\|2021-07-26 | 21A (Delta) |
| hCoV-19/South_Africa/Tygerberg_1545/2021\|EPI_ISL_3482556\|2021-07-26 | 21A (Delta) |
| hCoV-19/South_Africa/Tygerberg_1546/2021\|EPI_ISL_3482557\|2021-07-26 | 21A (Delta) |
| hCoV-19/South_Africa/Tygerberg_1547/2021\|EPI_ISL_3482559\|2021-07-26 | 21A (Delta) |
| hCoV-19/South_Africa/Tygerberg_1548/2021\|EPI_ISL_3485265\|2021-07-26 | 21A (Delta) |
| hCoV-19/South_Africa/Tygerberg_1549/2021\|EPI_ISL_3482561\|2021-07-26 | 21A (Delta) |
| hCoV-19/South_Africa/Tygerberg_1550/2021\|EPI_ISL_3482562\|2021-07-26 | 21A (Delta) |
| hCoV-19/South_Africa/Tygerberg_1551/2021\|EPI_ISL_3482564\|2021-07-26 | 21A (Delta) |
| hCoV-19/South_Africa/Tygerberg_1552/2021\|EPI_ISL_3482565\|2021-07-26 | 21A (Delta) |
| hCoV-19/South_Africa/Tygerberg_1553/2021\|EPI_ISL_3482567\|2021-07-26 | 21A (Delta) |
| hCoV-19/South_Africa/Tygerberg_1554/2021\|EPI_ISL_3482569\|2021-07-26 | 21A (Delta) |
| hCoV-19/South_Africa/Tygerberg_1555/2021\|EPI_ISL_3482571\|2021-07-26 | 21A (Delta) |
| hCoV-19/South_Africa/Tygerberg_1556/2021\|EPI_ISL_3482572\|2021-07-26 | 21A (Delta) |
| hCoV-19/South_Africa/Tygerberg_1557/2021\|EPI_ISL_3482574\|2021-07-26 | 21A (Delta) |
| hCoV-19/South_Africa/Tygerberg_1558/2021\|EPI_ISL_3482575\|2021-07-24 | 21A (Delta) |
| hCoV-19/South_Africa/Tygerberg_1559/2021\|EPI_ISL_3482577\|2021-06-30 | 21A (Delta) |
| hCoV-19/South_Africa/Tygerberg_1561/2021\|EPI_ISL_3482579\|2021-06-29 | 21A (Delta) |
| hCoV-19/South_Africa/Tygerberg_1562/2021\|EPI_ISL_3482580\|2021-06-29 | 21A (Delta) |
| hCoV-19/South_Africa/Tygerberg_1563/2021\|EPI_ISL_3482582\|2021-06-28 | 21A (Delta) |
| hCoV-19/South_Africa/Tygerberg_1565/2021\|EPI_ISL_3482583\|2021-06-29 | 21A (Delta) |
| hCoV-19/South_Africa/Tygerberg_1566/2021\|EPI_ISL_3482585\|2021-06-29 | 21A (Delta) |

| | |
|---|---|
| hCoV-19/South_Africa/Tygerberg_1567/2021\|EPI_ISL_3482587\|2021-06-29 | 21A (Delta) |
| hCoV-19/South_Africa/Tygerberg_1568/2021\|EPI_ISL_3482588\|2021-06-29 | 20H (Beta, V2) |
| hCoV-19/South_Africa/Tygerberg_1569/2021\|EPI_ISL_3482590\|2021-06-20 | 21A (Delta) |
| hCoV-19/South_Africa/Tygerberg_1570/2021\|EPI_ISL_3482592\|2021-06-19 | 20H (Beta, V2) |
| hCoV-19/South_Africa/Tygerberg_1571/2021\|EPI_ISL_3482593\|2021-06-22 | 21A (Delta) |
| hCoV-19/South_Africa/Tygerberg_1572/2021\|EPI_ISL_3482595\|2021-07-22 | 21A (Delta) |
| hCoV-19/South_Africa/Tygerberg_1574/2021\|EPI_ISL_3482598\|2021-06-21 | 21A (Delta) |
| hCoV-19/South_Africa/Tygerberg_1575/2021\|EPI_ISL_3482600\|2021-06-18 | 20H (Beta, V2) |
| hCoV-19/South_Africa/Tygerberg_1576/2021\|EPI_ISL_3482601\|2021-06-18 | 21A (Delta) |
| hCoV-19/South_Africa/Tygerberg_1577/2021\|EPI_ISL_3482603\|2021-07-26 | 21A (Delta) |
| hCoV-19/South_Africa/Tygerberg_1582/2021\|EPI_ISL_3482605\|2021-07-22 | 21A (Delta) |
| hCoV-19/South_Africa/Tygerberg_1583/2021\|EPI_ISL_3482606\|2021-07-22 | 21A (Delta) |
| hCoV-19/South_Africa/Tygerberg_1584/2021\|EPI_ISL_3482608\|2021-07-22 | 21A (Delta) |
| hCoV-19/South_Africa/Tygerberg_1585/2021\|EPI_ISL_3482610\|2021-07-22 | 21A (Delta) |
| hCoV-19/South_Africa/Tygerberg_1586/2021\|EPI_ISL_3482613\|2021-07-23 | 21A (Delta) |
| hCoV-19/South_Africa/Tygerberg_1587/2021\|EPI_ISL_3482615\|2021-07-26 | 21A (Delta) |
| hCoV-19/South_Africa/Tygerberg_1588/2021\|EPI_ISL_3482617\|2021-07-26 | 21A (Delta) |
| hCoV-19/South_Africa/Tygerberg_1589/2021\|EPI_ISL_3482620\|2021-07-26 | 21A (Delta) |
| hCoV-19/South_Africa/Tygerberg_1590/2021\|EPI_ISL_3482623\|2021-07-26 | 21A (Delta) |
| hCoV-19/South_Africa/Tygerberg_1591/2021\|EPI_ISL_3482625\|2021-07-23 | 21A (Delta) |
| hCoV-19/South_Africa/Tygerberg_1592/2021\|EPI_ISL_3482628\|2021-06-28 | 21A (Delta) |
| hCoV-19/South_Africa/Tygerberg_1593/2021\|EPI_ISL_3482630\|2021-06-20 | 21A (Delta) |
| hCoV-19/South_Africa/Tygerberg_1969/2021\|EPI_ISL_3666141\|2021-06-16 | 21A (Delta) |
| hCoV-19/South_Africa/Tygerberg_1971/2021\|EPI_ISL_3824214\|2021-06-15 | 20B |
| hCoV-19/South_Africa/Tygerberg_1972/2021\|EPI_ISL_3824215\|2021-06-17 | 21A (Delta) |
| hCoV-19/South_Africa/Tygerberg_1973/2021\|EPI_ISL_3824216\|2021-06-14 | 21A (Delta) |
| hCoV-19/South_Africa/Tygerberg_1974/2021\|EPI_ISL_3824217\|2021-06-18 | 21A (Delta) |
| hCoV-19/South_Africa/Tygerberg_1975/2021\|EPI_ISL_3824218\|2021-06-12 | 20H (Beta, V2) |
| hCoV-19/South_Africa/Tygerberg_1976/2021\|EPI_ISL_3824219\|2021-06-12 | 20I (Alpha, V1) |
| hCoV-19/South_Africa/Tygerberg_1977/2021\|EPI_ISL_3666130\|2021-06-12 | 21A (Delta) |
| hCoV-19/South_Africa/Tygerberg_1978/2021\|EPI_ISL_3666142\|2021-06-11 | 21A (Delta) |
| hCoV-19/South_Africa/Tygerberg_1979/2021\|EPI_ISL_3666163\|2021-06-11 | 20H (Beta, V2) |
| hCoV-19/South_Africa/Tygerberg_1980/2021\|EPI_ISL_3666159\|2021-06-13 | 20I (Alpha, V1) |
| hCoV-19/South_Africa/Tygerberg_1981/2021\|EPI_ISL_3666164\|2021-06-09 | 20H (Beta, V2) |
| hCoV-19/South_Africa/Tygerberg_1982/2021\|EPI_ISL_3666143\|2021-06-16 | 21A (Delta) |
| hCoV-19/South_Africa/Tygerberg_1983/2021\|EPI_ISL_3666129\|2021-06-27 | 21A (Delta) |
| hCoV-19/South_Africa/Tygerberg_1985/2021\|EPI_ISL_3666131\|2021-06-02 | 20H (Beta, V2) |
| hCoV-19/South_Africa/Tygerberg_1987/2021\|EPI_ISL_3666133\|2021-06-04 | 21A (Delta) |
| hCoV-19/South_Africa/Tygerberg_1988/2021\|EPI_ISL_3666136\|2021-06-29 | 21A (Delta) |
| hCoV-19/South_Africa/Tygerberg_1989/2021\|EPI_ISL_3666165\|2021-06-30 | 20H (Beta, V2) |
| hCoV-19/South_Africa/Tygerberg_1990/2021\|EPI_ISL_3666144\|2021-06-24 | 21A (Delta) |
| hCoV-19/South_Africa/Tygerberg_1991/2021\|EPI_ISL_3666134\|2021-08-09 | 21A (Delta) |
| hCoV-19/South_Africa/Tygerberg_1992/2021\|EPI_ISL_3666145\|2021-08-09 | 21A (Delta) |
| hCoV-19/South_Africa/Tygerberg_1993/2021\|EPI_ISL_3666146\|2021-08-09 | 21A (Delta) |
| hCoV-19/South_Africa/Tygerberg_1994/2021\|EPI_ISL_3666137\|2021-08-09 | 21A (Delta) |
| hCoV-19/South_Africa/Tygerberg_1995/2021\|EPI_ISL_3666147\|2021-08-09 | 21A (Delta) |
| hCoV-19/South_Africa/Tygerberg_1996/2021\|EPI_ISL_3666148\|2021-08-09 | 21A (Delta) |
| hCoV-19/South_Africa/Tygerberg_1997/2021\|EPI_ISL_3666149\|2021-08-09 | 21A (Delta) |
| hCoV-19/South_Africa/Tygerberg_1998/2021\|EPI_ISL_3666161\|2021-08-09 | 21A (Delta) |
| hCoV-19/South_Africa/Tygerberg_1999/2021\|EPI_ISL_3666150\|2021-08-09 | 21A (Delta) |

| | |
|---|---|
| hCoV-19/South_Africa/Tygerberg_2000/2021\|EPI_ISL_3666138\|2021-08-09 | 21A (Delta) |
| hCoV-19/South_Africa/Tygerberg_2001/2021\|EPI_ISL_3666128\|2021-08-09 | 21A (Delta) |
| hCoV-19/South_Africa/Tygerberg_2002/2021\|EPI_ISL_3666132\|2021-08-09 | 21A (Delta) |
| hCoV-19/South_Africa/Tygerberg_2003/2021\|EPI_ISL_3666139\|2021-08-09 | 21A (Delta) |
| hCoV-19/South_Africa/Tygerberg_2004/2021\|EPI_ISL_3666151\|2021-08-09 | 21A (Delta) |
| hCoV-19/South_Africa/Tygerberg_2005/2021\|EPI_ISL_3666152\|2021-08-09 | 21A (Delta) |
| hCoV-19/South_Africa/Tygerberg_2006/2021\|EPI_ISL_3666160\|2021-08-09 | 21A (Delta) |
| hCoV-19/South_Africa/Tygerberg_2007/2021\|EPI_ISL_3666153\|2021-08-09 | 21A (Delta) |
| hCoV-19/South_Africa/Tygerberg_2008/2021\|EPI_ISL_3666154\|2021-08-09 | 21A (Delta) |
| hCoV-19/South_Africa/Tygerberg_2009/2021\|EPI_ISL_3666140\|2021-08-09 | 21A (Delta) |
| hCoV-19/South_Africa/Tygerberg_2010/2021\|EPI_ISL_3666135\|2021-08-09 | 21A (Delta) |
| hCoV-19/South_Africa/Tygerberg_2011/2021\|EPI_ISL_3666158\|2021-08-09 | 21A (Delta) |
| hCoV-19/South_Africa/Tygerberg_2012/2021\|EPI_ISL_3666155\|2021-08-09 | 21A (Delta) |
| hCoV-19/South_Africa/Tygerberg_2013/2021\|EPI_ISL_3666156\|2021-08-09 | 21A (Delta) |
| hCoV-19/South_Africa/Tygerberg_2015/2021\|EPI_ISL_3666157\|2021-08-09 | 21A (Delta) |
| hCoV-19/South_Africa/Tygerberg_2025/2021\|EPI_ISL_3666714\|2021-08-02 | 21A (Delta) |
| hCoV-19/South_Africa/Tygerberg_2026/2021\|EPI_ISL_3666715\|2021-08-03 | 21A (Delta) |
| hCoV-19/South_Africa/Tygerberg_2027/2021\|EPI_ISL_3666716\|2021-08-04 | 21A (Delta) |
| hCoV-19/South_Africa/Tygerberg_2028/2021\|EPI_ISL_3666717\|2021-08-03 | 21A (Delta) |
| hCoV-19/South_Africa/Tygerberg_2029/2021\|EPI_ISL_3666718\|2021-08-03 | 21A (Delta) |
| hCoV-19/South_Africa/Tygerberg_2030/2021\|EPI_ISL_3666719\|2021-08-03 | 21A (Delta) |
| hCoV-19/South_Africa/Tygerberg_2031/2021\|EPI_ISL_3666720\|2021-08-03 | 21A (Delta) |
| hCoV-19/South_Africa/Tygerberg_2032/2021\|EPI_ISL_3666721\|2021-08-04 | 21A (Delta) |
| hCoV-19/South_Africa/Tygerberg_2033/2021\|EPI_ISL_3666722\|2021-08-05 | 21A (Delta) |
| hCoV-19/South_Africa/Tygerberg_2034/2021\|EPI_ISL_3666723\|2021-08-05 | 21A (Delta) |
| hCoV-19/South_Africa/Tygerberg_2035/2021\|EPI_ISL_3666724\|2021-08-05 | 21A (Delta) |
| hCoV-19/South_Africa/Tygerberg_2036/2021\|EPI_ISL_3666725\|2021-08-05 | 21A (Delta) |
| hCoV-19/South_Africa/Tygerberg_2037/2021\|EPI_ISL_3666726\|2021-08-05 | 21A (Delta) |
| hCoV-19/South_Africa/Tygerberg_2038/2021\|EPI_ISL_3666727\|2021-08-05 | 21A (Delta) |
| hCoV-19/South_Africa/Tygerberg_2039/2021\|EPI_ISL_3870856\|2021-08-05 | 21A (Delta) |
| hCoV-19/South_Africa/Tygerberg_2040/2021\|EPI_ISL_3666728\|2021-08-05 | 21A (Delta) |
| hCoV-19/South_Africa/Tygerberg_2041/2021\|EPI_ISL_3666729\|2021-08-05 | 21A (Delta) |
| hCoV-19/South_Africa/Tygerberg_2042/2021\|EPI_ISL_3666730\|2021-08-05 | 21A (Delta) |
| hCoV-19/South_Africa/Tygerberg_2043/2021\|EPI_ISL_3666731\|2021-08-05 | 21A (Delta) |
| hCoV-19/South_Africa/Tygerberg_2044/2021\|EPI_ISL_3666732\|2021-08-05 | 21A (Delta) |
| hCoV-19/South_Africa/Tygerberg_2045/2021\|EPI_ISL_3666733\|2021-08-05 | 21A (Delta) |
| hCoV-19/South_Africa/Tygerberg_2048/2021\|EPI_ISL_3666734\|2021-08-05 | 21A (Delta) |
| hCoV-19/South_Africa/Tygerberg_2049/2021\|EPI_ISL_3666735\|2021-08-05 | 21A (Delta) |
| hCoV-19/South_Africa/Tygerberg_2050/2021\|EPI_ISL_3666736\|2021-08-05 | 21A (Delta) |
| hCoV-19/South_Africa/Tygerberg_2051/2021\|EPI_ISL_3666737\|2021-08-05 | 21A (Delta) |
| hCoV-19/South_Africa/Tygerberg_2052/2021\|EPI_ISL_3666738\|2021-08-05 | 21A (Delta) |
| hCoV-19/South_Africa/Tygerberg_2053/2021\|EPI_ISL_3666739\|2021-08-05 | 21A (Delta) |
| hCoV-19/South_Africa/Tygerberg_2054/2021\|EPI_ISL_3666740\|2021-08-05 | 21A (Delta) |
| hCoV-19/South_Africa/Tygerberg_2055/2021\|EPI_ISL_3666741\|2021-08-05 | 21A (Delta) |
| hCoV-19/South_Africa/Tygerberg_2056/2021\|EPI_ISL_3666742\|2021-08-05 | 21A (Delta) |
| hCoV-19/South_Africa/Tygerberg_2057/2021\|EPI_ISL_3666743\|2021-08-05 | 21A (Delta) |
| hCoV-19/South_Africa/Tygerberg_2058/2021\|EPI_ISL_3666744\|2021-08-05 | 21A (Delta) |
| hCoV-19/South_Africa/Tygerberg_2059/2021\|EPI_ISL_3666745\|2021-08-05 | 21A (Delta) |
| hCoV-19/South_Africa/Tygerberg_2060/2021\|EPI_ISL_3666746\|2021-08-05 | 21A (Delta) |
| hCoV-19/South_Africa/Tygerberg_2061/2021\|EPI_ISL_3666747\|2021-08-05 | 21A (Delta) |

| | |
|---|---|
| hCoV-19/South_Africa/Tygerberg_2062/2021\|EPI_ISL_3666748\|2021-08-05 | 21A (Delta) |
| hCoV-19/South_Africa/Tygerberg_2063/2021\|EPI_ISL_3666749\|2021-08-05 | 21A (Delta) |
| hCoV-19/South_Africa/Tygerberg_2064/2021\|EPI_ISL_3666750\|2021-08-05 | 21A (Delta) |
| hCoV-19/South_Africa/Tygerberg_2065/2021\|EPI_ISL_3666751\|2021-08-05 | 21A (Delta) |
| hCoV-19/South_Africa/Tygerberg_2066/2021\|EPI_ISL_3666752\|2021-08-05 | 21A (Delta) |
| hCoV-19/South_Africa/Tygerberg_2067/2021\|EPI_ISL_3666753\|2021-08-05 | 21A (Delta) |
| hCoV-19/South_Africa/Tygerberg_2068/2021\|EPI_ISL_3666754\|2021-08-05 | 21A (Delta) |
| hCoV-19/South_Africa/Tygerberg_2069/2021\|EPI_ISL_3870858\|2021-08-05 | 21A (Delta) |
| hCoV-19/South_Africa/Tygerberg_2070/2021\|EPI_ISL_3666755\|2021-08-05 | 21A (Delta) |
| hCoV-19/South_Africa/Tygerberg_2071/2021\|EPI_ISL_3666756\|2021-08-05 | 21A (Delta) |
| hCoV-19/South_Africa/Tygerberg_2072/2021\|EPI_ISL_3666757\|2021-08-05 | 21A (Delta) |
| hCoV-19/South_Africa/Tygerberg_2073/2021\|EPI_ISL_3666758\|2021-08-05 | 21A (Delta) |
| hCoV-19/South_Africa/Tygerberg_2074/2021\|EPI_ISL_3666759\|2021-08-05 | 21A (Delta) |
| hCoV-19/South_Africa/Tygerberg_2075/2021\|EPI_ISL_3666760\|2021-08-05 | 21A (Delta) |
| hCoV-19/South_Africa/Tygerberg_2076/2021\|EPI_ISL_3666761\|2021-08-05 | 21A (Delta) |
| hCoV-19/South_Africa/Tygerberg_2077/2021\|EPI_ISL_3666762\|2021-08-05 | 21A (Delta) |
| hCoV-19/South_Africa/Tygerberg_2078/2021\|EPI_ISL_3666763\|2021-08-05 | 21A (Delta) |
| hCoV-19/South_Africa/Tygerberg_2079/2021\|EPI_ISL_3666764\|2021-08-05 | 21A (Delta) |
| hCoV-19/South_Africa/Tygerberg_2080/2021\|EPI_ISL_3666765\|2021-08-05 | 21A (Delta) |
| hCoV-19/South_Africa/Tygerberg_2081/2021\|EPI_ISL_3666766\|2021-08-05 | 21A (Delta) |
| hCoV-19/South_Africa/Tygerberg_2082/2021\|EPI_ISL_3666767\|2021-08-05 | 21A (Delta) |
| hCoV-19/South_Africa/Tygerberg_2083/2021\|EPI_ISL_3666768\|2021-08-06 | 21A (Delta) |
| hCoV-19/South_Africa/Tygerberg_2084/2021\|EPI_ISL_3666769\|2021-08-06 | 21A (Delta) |
| hCoV-19/South_Africa/Tygerberg_2085/2021\|EPI_ISL_3666770\|2021-08-05 | 21A (Delta) |
| hCoV-19/South_Africa/Tygerberg_2086/2021\|EPI_ISL_3666771\|2021-08-05 | 21A (Delta) |
| hCoV-19/South_Africa/Tygerberg_2087/2021\|EPI_ISL_3666772\|2021-08-05 | 21A (Delta) |
| hCoV-19/South_Africa/Tygerberg_2088/2021\|EPI_ISL_3666773\|2021-08-05 | 21A (Delta) |
| hCoV-19/South_Africa/Tygerberg_2089/2021\|EPI_ISL_3666774\|2021-08-05 | 21A (Delta) |
| hCoV-19/South_Africa/Tygerberg_2090/2021\|EPI_ISL_3666775\|2021-08-05 | 21A (Delta) |
| hCoV-19/South_Africa/Tygerberg_2091/2021\|EPI_ISL_3666776\|2021-08-05 | 21A (Delta) |
| hCoV-19/South_Africa/Tygerberg_2092/2021\|EPI_ISL_3666777\|2021-08-05 | 21A (Delta) |
| hCoV-19/South_Africa/Tygerberg_2093/2021\|EPI_ISL_3666778\|2021-08-05 | 21A (Delta) |
| hCoV-19/South_Africa/Tygerberg_2094/2021\|EPI_ISL_3666779\|2021-08-05 | 21A (Delta) |
| hCoV-19/South_Africa/Tygerberg_2095/2021\|EPI_ISL_3666780\|2021-08-05 | 21A (Delta) |
| hCoV-19/South_Africa/Tygerberg_2096/2021\|EPI_ISL_3666781\|2021-08-05 | 21A (Delta) |
| hCoV-19/South_Africa/Tygerberg_2097/2021\|EPI_ISL_3666782\|2021-08-05 | 21A (Delta) |
| hCoV-19/South_Africa/Tygerberg_2098/2021\|EPI_ISL_3666783\|2021-08-05 | 21A (Delta) |
| hCoV-19/South_Africa/Tygerberg_2099/2021\|EPI_ISL_3666784\|2021-08-05 | 21A (Delta) |
| hCoV-19/South_Africa/Tygerberg_2100/2021\|EPI_ISL_3666785\|2021-08-05 | 21A (Delta) |
| hCoV-19/South_Africa/Tygerberg_2101/2021\|EPI_ISL_3666786\|2021-08-05 | 21A (Delta) |
| hCoV-19/South_Africa/Tygerberg_2102/2021\|EPI_ISL_3666787\|2021-08-05 | 21A (Delta) |
| hCoV-19/South_Africa/Tygerberg_2103/2021\|EPI_ISL_3666788\|2021-08-05 | 21A (Delta) |
| hCoV-19/South_Africa/Tygerberg_2104/2021\|EPI_ISL_3666789\|2021-08-06 | 21A (Delta) |
| hCoV-19/South_Africa/Tygerberg_2105/2021\|EPI_ISL_3666790\|2021-08-05 | 21A (Delta) |
| hCoV-19/South_Africa/Tygerberg_2106/2021\|EPI_ISL_3666791\|2021-08-06 | 21A (Delta) |
| hCoV-19/South_Africa/Tygerberg_2107/2021\|EPI_ISL_3666792\|2021-08-06 | 21A (Delta) |
| hCoV-19/South_Africa/Tygerberg_2108/2021\|EPI_ISL_3666793\|2021-08-06 | 21A (Delta) |
| hCoV-19/South_Africa/Tygerberg_2109/2021\|EPI_ISL_3666794\|2021-08-10 | 21A (Delta) |
| hCoV-19/South_Africa/Tygerberg_2111/2021\|EPI_ISL_3666795\|2021-08-10 | 21A (Delta) |
| hCoV-19/South_Africa/Tygerberg_2112/2021\|EPI_ISL_3666796\|2021-08-10 | 21A (Delta) |

| | |
|---|---|
| hCoV-19/South_Africa/Tygerberg_2120/2021\|EPI_ISL_4193881\|2021-08-11 | 21A (Delta) |
| hCoV-19/South_Africa/Tygerberg_2128/2021\|EPI_ISL_4193888\|2021-08-10 | 21A (Delta) |
| hCoV-19/South_Africa/Tygerberg_2146/2021\|EPI_ISL_4193897\|2021-08-10 | 21A (Delta) |
| hCoV-19/South_Africa/Tygerberg_2152/2021\|EPI_ISL_4193914\|2021-08-10 | 21A (Delta) |
| hCoV-19/South_Africa/Tygerberg_2154/2021\|EPI_ISL_4193855\|2021-08-10 | 21A (Delta) |
| hCoV-19/South_Africa/Tygerberg_2160/2021\|EPI_ISL_4193863\|2021-08-11 | 21A (Delta) |
| hCoV-19/South_Africa/Tygerberg_2168/2021\|EPI_ISL_4193872\|2021-08-11 | 21A (Delta) |
| hCoV-19/South_Africa/Tygerberg_2176/2021\|EPI_ISL_4193882\|2021-08-10 | 21A (Delta) |
| hCoV-19/South_Africa/Tygerberg_2184/2021\|EPI_ISL_4193889\|2021-08-10 | 21A (Delta) |
| hCoV-19/South_Africa/Tygerberg_2192/2021\|EPI_ISL_4193898\|2021-08-11 | 21A (Delta) |
| hCoV-19/South_Africa/Tygerberg_2200/2021\|EPI_ISL_4193906\|2021-08-11 | 21A (Delta) |
| hCoV-19/South_Africa/Tygerberg_2203/2021\|EPI_ISL_4193915\|2021-08-12 | 21A (Delta) |
| hCoV-19/South_Africa/Tygerberg_2206/2021\|EPI_ISL_4193856\|2021-08-12 | 21A (Delta) |
| hCoV-19/South_Africa/Tygerberg_2226/2021\|EPI_ISL_4193864\|2021-08-16 | 21A (Delta) |
| hCoV-19/South_Africa/Tygerberg_2234/2021\|EPI_ISL_4193873\|2021-08-15 | 21A (Delta) |
| hCoV-19/South_Africa/Tygerberg_2236/2021\|EPI_ISL_3872142\|2021-08-14 | 21A (Delta) |
| hCoV-19/South_Africa/Tygerberg_2242/2021\|EPI_ISL_4193919\|2021-08-16 | 21A (Delta) |
| hCoV-19/South_Africa/Tygerberg_2251/2021\|EPI_ISL_4193890\|2021-08-16 | 21A (Delta) |
| hCoV-19/South_Africa/Tygerberg_2259/2021\|EPI_ISL_4193907\|2021-08-16 | 21A (Delta) |
| hCoV-19/South_Africa/Tygerberg_2266/2021\|EPI_ISL_4193923\|2021-08-17 | 21A (Delta) |
| hCoV-19/South_Africa/Tygerberg_2267/2021\|EPI_ISL_4193922\|2021-08-17 | 21A (Delta) |
| hCoV-19/South_Africa/Tygerberg_2274/2021\|EPI_ISL_4193874\|2021-08-18 | 21A (Delta) |
| hCoV-19/South_Africa/Tygerberg_2291/2021\|EPI_ISL_4193891\|2021-08-20 | 21A (Delta) |
| hCoV-19/South_Africa/Tygerberg_2296/2021\|EPI_ISL_4193853\|2021-08-23 | 21A (Delta) |
| hCoV-19/South_Africa/Tygerberg_2297/2021\|EPI_ISL_4193859\|2021-08-20 | 21A (Delta) |
| hCoV-19/South_Africa/Tygerberg_2298/2021\|EPI_ISL_4193867\|2021-08-23 | 21A (Delta) |
| hCoV-19/South_Africa/Tygerberg_2299/2021\|EPI_ISL_4193877\|2021-08-22 | 21A (Delta) |
| hCoV-19/South_Africa/Tygerberg_2300/2021\|EPI_ISL_4193885\|2021-08-19 | 21A (Delta) |
| hCoV-19/South_Africa/Tygerberg_2301/2021\|EPI_ISL_4193894\|2021-08-22 | 21A (Delta) |
| hCoV-19/South_Africa/Tygerberg_2302/2021\|EPI_ISL_4193902\|2021-08-20 | 21A (Delta) |
| hCoV-19/South_Africa/Tygerberg_2303/2021\|EPI_ISL_4193911\|2021-08-20 | 21A (Delta) |
| hCoV-19/South_Africa/Tygerberg_2305/2021\|EPI_ISL_4193860\|2021-08-22 | 21A (Delta) |
| hCoV-19/South_Africa/Tygerberg_2306/2021\|EPI_ISL_4193868\|2021-08-21 | 21A (Delta) |
| hCoV-19/South_Africa/Tygerberg_2307/2021\|EPI_ISL_4193878\|2021-06-22 | 21A (Delta) |
| hCoV-19/South_Africa/Tygerberg_2308/2021\|EPI_ISL_4193886\|2021-08-20 | 21A (Delta) |
| hCoV-19/South_Africa/Tygerberg_2309/2021\|EPI_ISL_4193895\|2021-08-20 | 21A (Delta) |
| hCoV-19/South_Africa/Tygerberg_2310/2021\|EPI_ISL_4193903\|2021-08-20 | 21A (Delta) |
| hCoV-19/South_Africa/Tygerberg_2311/2021\|EPI_ISL_4193912\|2021-08-22 | 21A (Delta) |
| hCoV-19/South_Africa/Tygerberg_2313/2021\|EPI_ISL_4193861\|2021-08-23 | 21A (Delta) |
| hCoV-19/South_Africa/Tygerberg_2314/2021\|EPI_ISL_4193869\|2021-08-20 | 21A (Delta) |
| hCoV-19/South_Africa/Tygerberg_2315/2021\|EPI_ISL_4193879\|2021-08-21 | 21A (Delta) |
| hCoV-19/South_Africa/Tygerberg_2316/2021\|EPI_ISL_4193887\|2021-08-22 | 21A (Delta) |
| hCoV-19/South_Africa/Tygerberg_2317/2021\|EPI_ISL_4193896\|2021-08-22 | 21A (Delta) |
| hCoV-19/South_Africa/Tygerberg_2319/2021\|EPI_ISL_4193913\|2021-06-23 | 20H (Beta, V2) |
| hCoV-19/South_Africa/Tygerberg_2320/2021\|EPI_ISL_4193854\|2021-08-22 | 21A (Delta) |
| hCoV-19/South_Africa/Tygerberg_2321/2021\|EPI_ISL_4193862\|2021-08-19 | 21A (Delta) |
| hCoV-19/South_Africa/Tygerberg_2322/2021\|EPI_ISL_4193870\|2021-08-19 | 21A (Delta) |
| hCoV-19/South_Africa/Tygerberg_2323/2021\|EPI_ISL_4193880\|2021-08-21 | 21A (Delta) |
| hCoV-19/South_Africa/Tygerberg_2325/2021\|EPI_ISL_4193904\|2021-06-25 | 21A (Delta) |
| hCoV-19/South_Africa/Tygerberg_2327/2021\|EPI_ISL_4193920\|2021-06-30 | 21A (Delta) |

| | |
|---|---|
| hCoV-19/South_Africa/Tygerberg_2328/2021\|EPI_ISL_4193921\|2021-06-30 | 20H (Beta, V2) |
| hCoV-19/South_Africa/Tygerberg_2329/2021\|EPI_ISL_4193871\|2021-08-27 | 21A (Delta) |
| hCoV-19/South_Africa/Tygerberg_2330/2021\|EPI_ISL_4193899\|2021-08-21 | 21A (Delta) |
| hCoV-19/South_Africa/Tygerberg_2331/2021\|EPI_ISL_4193908\|2021-08-21 | 21A (Delta) |
| hCoV-19/South_Africa/Tygerberg_2332/2021\|EPI_ISL_4193916\|2021-08-22 | 21A (Delta) |
| hCoV-19/South_Africa/Tygerberg_2333/2021\|EPI_ISL_4193857\|2021-08-22 | 21A (Delta) |
| hCoV-19/South_Africa/Tygerberg_2334/2021\|EPI_ISL_4193865\|2021-08-23 | 21A (Delta) |
| hCoV-19/South_Africa/Tygerberg_2335/2021\|EPI_ISL_4193875\|2021-08-23 | 21A (Delta) |
| hCoV-19/South_Africa/Tygerberg_2336/2021\|EPI_ISL_4193883\|2021-08-23 | 21A (Delta) |
| hCoV-19/South_Africa/Tygerberg_2337/2021\|EPI_ISL_4193892\|2021-08-23 | 21A (Delta) |
| hCoV-19/South_Africa/Tygerberg_2338/2021\|EPI_ISL_4193900\|2021-08-23 | 21A (Delta) |
| hCoV-19/South_Africa/Tygerberg_2339/2021\|EPI_ISL_4193909\|2021-08-23 | 21A (Delta) |
| hCoV-19/South_Africa/Tygerberg_2340/2021\|EPI_ISL_4193917\|2021-08-23 | 21A (Delta) |
| hCoV-19/South_Africa/Tygerberg_2342/2021\|EPI_ISL_4193866\|2021-08-24 | 21A (Delta) |
| hCoV-19/South_Africa/Tygerberg_2343/2021\|EPI_ISL_4193876\|2021-08-25 | 21A (Delta) |
| hCoV-19/South_Africa/Tygerberg_2344/2021\|EPI_ISL_4193884\|2021-08-25 | 21A (Delta) |
| hCoV-19/South_Africa/Tygerberg_2345/2021\|EPI_ISL_4193893\|2021-08-25 | 21A (Delta) |
| hCoV-19/South_Africa/Tygerberg_2346/2021\|EPI_ISL_4193901\|2021-08-25 | 21A (Delta) |
| hCoV-19/South_Africa/Tygerberg_2347/2021\|EPI_ISL_4193910\|2021-08-27 | 21A (Delta) |
| hCoV-19/South_Africa/Tygerberg_2348/2021\|EPI_ISL_4193918\|2021-08-27 | 21A (Delta) |
| hCoV-19/South_Africa/Tygerberg_2349/2021\|EPI_ISL_4193858\|2021-08-28 | 21A (Delta) |
| hCoV-19/South_Africa/Tygerberg_2350/2021\|EPI_ISL_4195417\|2021-08-30 | 21A (Delta) |
| hCoV-19/South_Africa/Tygerberg_2351/2021\|EPI_ISL_4195418\|2021-08-30 | 21A (Delta) |
| hCoV-19/South_Africa/Tygerberg_2352/2021\|EPI_ISL_4195419\|2021-08-30 | 21A (Delta) |
| hCoV-19/South_Africa/Tygerberg_2353/2021\|EPI_ISL_4195420\|2021-08-30 | 21A (Delta) |
| hCoV-19/South_Africa/Tygerberg_2354/2021\|EPI_ISL_4195421\|2021-08-30 | 21A (Delta) |
| hCoV-19/South_Africa/Tygerberg_2355/2021\|EPI_ISL_4195422\|2021-08-30 | 21A (Delta) |
| hCoV-19/South_Africa/Tygerberg_2356/2021\|EPI_ISL_4195423\|2021-08-30 | 21A (Delta) |
| hCoV-19/South_Africa/Tygerberg_2357/2021\|EPI_ISL_4195424\|2021-08-30 | 21A (Delta) |
| hCoV-19/South_Africa/Tygerberg_2359/2021\|EPI_ISL_4195426\|2021-08-30 | 21A (Delta) |
| hCoV-19/South_Africa/Tygerberg_2360/2021\|EPI_ISL_4195427\|2021-08-30 | 21A (Delta) |
| hCoV-19/South_Africa/Tygerberg_2361/2021\|EPI_ISL_4195428\|2021-08-30 | 21A (Delta) |
| hCoV-19/South_Africa/Tygerberg_2366/2021\|EPI_ISL_4195432\|2021-08-28 | 21A (Delta) |
| hCoV-19/South_Africa/Tygerberg_2369/2021\|EPI_ISL_4195435\|2021-08-31 | 21A (Delta) |
| hCoV-19/South_Africa/Tygerberg_2370/2021\|EPI_ISL_4195436\|2021-08-31 | 21A (Delta) |
| hCoV-19/South_Africa/Tygerberg_2371/2021\|EPI_ISL_4195437\|2021-08-31 | 21A (Delta) |
| hCoV-19/South_Africa/Tygerberg_2372/2021\|EPI_ISL_4195438\|2021-08-31 | 21A (Delta) |
| hCoV-19/South_Africa/Tygerberg_2373/2021\|EPI_ISL_4195439\|2021-08-31 | 21A (Delta) |
| hCoV-19/South_Africa/Tygerberg_2374/2021\|EPI_ISL_4195440\|2021-08-31 | 21A (Delta) |
| hCoV-19/South_Africa/Tygerberg_2375/2021\|EPI_ISL_4195441\|2021-08-31 | 21A (Delta) |
| hCoV-19/South_Africa/Tygerberg_2376/2021\|EPI_ISL_4195442\|2021-08-31 | 21A (Delta) |
| hCoV-19/South_Africa/Tygerberg_2390/2021\|EPI_ISL_4195456\|2021-08-30 | 21A (Delta) |
| hCoV-19/South_Africa/Tygerberg_2391/2021\|EPI_ISL_4195457\|2021-08-30 | 21A (Delta) |
| hCoV-19/South_Africa/Tygerberg_2392/2021\|EPI_ISL_4195458\|2021-08-30 | 21A (Delta) |
| hCoV-19/South_Africa/Tygerberg_2393/2021\|EPI_ISL_4195459\|2021-08-30 | 21A (Delta) |
| hCoV-19/South_Africa/Tygerberg_2394/2021\|EPI_ISL_4195460\|2021-08-30 | 21A (Delta) |
| hCoV-19/South_Africa/Tygerberg_2395/2021\|EPI_ISL_4195461\|2021-08-29 | 21A (Delta) |
| hCoV-19/South_Africa/Tygerberg_2396/2021\|EPI_ISL_4195462\|2021-08-27 | 21A (Delta) |
| hCoV-19/South_Africa/Tygerberg_2397/2021\|EPI_ISL_4195463\|2021-08-30 | 21A (Delta) |
| hCoV-19/South_Africa/Tygerberg_2398/2021\|EPI_ISL_4195464\|2021-08-30 | 21A (Delta) |

| | |
|---|---|
| hCoV-19/South_Africa/Tygerberg_2399/2021\|EPI_ISL_4195465\|2021-08-30 | 21A (Delta) |
| hCoV-19/South_Africa/Tygerberg_2400/2021\|EPI_ISL_4195466\|2021-08-30 | 21A (Delta) |
| hCoV-19/South_Africa/Tygerberg_2401/2021\|EPI_ISL_4195467\|2021-08-30 | 21A (Delta) |
| hCoV-19/South_Africa/Tygerberg_2402/2021\|EPI_ISL_4195468\|2021-08-30 | 21A (Delta) |
| hCoV-19/South_Africa/Tygerberg_2403/2021\|EPI_ISL_4195469\|2021-08-26 | 21A (Delta) |
| hCoV-19/South_Africa/Tygerberg_2404/2021\|EPI_ISL_4195470\|2021-08-31 | 21A (Delta) |
| hCoV-19/South_Africa/Tygerberg_2405/2021\|EPI_ISL_4195471\|2021-08-27 | 21A (Delta) |
| hCoV-19/South_Africa/Tygerberg_2406/2021\|EPI_ISL_4195472\|2021-08-31 | 21A (Delta) |
| hCoV-19/South_Africa/Tygerberg_2407/2021\|EPI_ISL_4195473\|2021-08-31 | 21A (Delta) |
| hCoV-19/South_Africa/Tygerberg_2408/2021\|EPI_ISL_4195474\|2021-08-30 | 21A (Delta) |
| hCoV-19/South_Africa/Tygerberg_2409/2021\|EPI_ISL_4195475\|2021-08-30 | 21A (Delta) |
| hCoV-19/South_Africa/Tygerberg_2410/2021\|EPI_ISL_4195476\|2021-08-30 | 21A (Delta) |
| hCoV-19/South_Africa/Tygerberg_2417/2021\|EPI_ISL_4195483\|2021-08-30 | 21A (Delta) |
| hCoV-19/South_Africa/Tygerberg_2420/2021\|EPI_ISL_4195486\|2021-08-31 | 21A (Delta) |
| hCoV-19/South_Africa/Tygerberg_2429/2021\|EPI_ISL_4195495\|2021-08-31 | 21A (Delta) |
| hCoV-19/South_Africa/Tygerberg_2430/2021\|EPI_ISL_4195496\|2021-08-31 | 21A (Delta) |
| hCoV-19/South_Africa/Tygerberg_2434/2021\|EPI_ISL_4195500\|2021-08-29 | 21A (Delta) |
| hCoV-19/South_Africa/Tygerberg_2436/2021\|EPI_ISL_4195501\|2021-08-28 | 21A (Delta) |
| hCoV-19/South_Africa/Tygerberg_2437/2021\|EPI_ISL_4195502\|2021-08-28 | 21A (Delta) |
| hCoV-19/South_Africa/Tygerberg_2438/2021\|EPI_ISL_4195503\|2021-08-28 | 21A (Delta) |
| hCoV-19/South_Africa/Tygerberg_2439/2021\|EPI_ISL_4195504\|2021-08-28 | 21A (Delta) |
| hCoV-19/South_Africa/Tygerberg_2440/2021\|EPI_ISL_4195505\|2021-08-29 | 21A (Delta) |
| hCoV-19/South_Africa/Tygerberg_2441/2021\|EPI_ISL_4195506\|2021-08-31 | 21A (Delta) |
| hCoV-19/South_Africa/Tygerberg_2442/2021\|EPI_ISL_4195507\|2021-08-28 | 21A (Delta) |
| hCoV-19/South_Africa/Tygerberg_2445/2021\|EPI_ISL_4250647\|2021-07-23 | 21A (Delta) |
| hCoV-19/South_Africa/Tygerberg_2446/2021\|EPI_ISL_4250693\|2021-07-19 | 21A (Delta) |
| hCoV-19/South_Africa/Tygerberg_2447/2021\|EPI_ISL_4250416\|2021-07-16 | 21A (Delta) |
| hCoV-19/South_Africa/Tygerberg_2448/2021\|EPI_ISL_4250745\|2021-07-24 | 21A (Delta) |
| hCoV-19/South_Africa/Tygerberg_2449/2021\|EPI_ISL_4250405\|2021-07-17 | 21A (Delta) |
| hCoV-19/South_Africa/Tygerberg_2451/2021\|EPI_ISL_4250683\|2021-07-17 | 21A (Delta) |
| hCoV-19/South_Africa/Tygerberg_2452/2021\|EPI_ISL_4250755\|2021-07-21 | 21A (Delta) |
| hCoV-19/South_Africa/Tygerberg_2453/2021\|EPI_ISL_4250539\|2021-07-19 | 21A (Delta) |
| hCoV-19/South_Africa/Tygerberg_2454/2021\|EPI_ISL_4250526\|2021-07-19 | 21A (Delta) |
| hCoV-19/South_Africa/Tygerberg_2455/2021\|EPI_ISL_4250431\|2021-07-30 | 21A (Delta) |
| hCoV-19/South_Africa/Tygerberg_2456/2021\|EPI_ISL_4250686\|2021-07-29 | 21A (Delta) |
| hCoV-19/South_Africa/Tygerberg_2457/2021\|EPI_ISL_4250550\|2021-07-29 | 21A (Delta) |
| hCoV-19/South_Africa/Tygerberg_2458/2021\|EPI_ISL_4250661\|2021-07-27 | 21A (Delta) |
| hCoV-19/South_Africa/Tygerberg_2459/2021\|EPI_ISL_4250644\|2021-07-28 | 21A (Delta) |
| hCoV-19/South_Africa/Tygerberg_2460/2021\|EPI_ISL_4250434\|2021-07-28 | 21A (Delta) |
| hCoV-19/South_Africa/Tygerberg_2461/2021\|EPI_ISL_4250638\|2021-07-25 | 21A (Delta) |
| hCoV-19/South_Africa/Tygerberg_2462/2021\|EPI_ISL_4250668\|2021-07-30 | 21A (Delta) |
| hCoV-19/South_Africa/Tygerberg_2463/2021\|EPI_ISL_4250408\|2021-07-26 | 21A (Delta) |
| hCoV-19/South_Africa/Tygerberg_2464/2021\|EPI_ISL_4250705\|2021-07-15 | 21A (Delta) |
| hCoV-19/South_Africa/Tygerberg_2465/2021\|EPI_ISL_4250506\|2021-07-07 | 21A (Delta) |
| hCoV-19/South_Africa/Tygerberg_2466/2021\|EPI_ISL_4250562\|2021-07-12 | 21A (Delta) |
| hCoV-19/South_Africa/Tygerberg_2467/2021\|EPI_ISL_4250742\|2021-07-13 | 21A (Delta) |
| hCoV-19/South_Africa/Tygerberg_2468/2021\|EPI_ISL_4250438\|2021-07-13 | 21A (Delta) |
| hCoV-19/South_Africa/Tygerberg_2469/2021\|EPI_ISL_4250772\|2021-07-14 | 21A (Delta) |
| hCoV-19/South_Africa/Tygerberg_2470/2021\|EPI_ISL_4250441\|2021-07-14 | 21A (Delta) |
| hCoV-19/South_Africa/Tygerberg_2471/2021\|EPI_ISL_4250718\|2021-07-14 | 21A (Delta) |

| | |
|---|---|
| hCoV-19/South_Africa/Tygerberg_2472/2021\|EPI_ISL_4250651\|2021-07-15 | 21A (Delta) |
| hCoV-19/South_Africa/Tygerberg_2474/2021\|EPI_ISL_4250671\|2021-07-26 | 21A (Delta) |
| hCoV-19/South_Africa/Tygerberg_2475/2021\|EPI_ISL_4250445\|2021-07-01 | 21A (Delta) |
| hCoV-19/South_Africa/Tygerberg_2476/2021\|EPI_ISL_4250448\|2021-07-01 | 21A (Delta) |
| hCoV-19/South_Africa/Tygerberg_2477/2021\|EPI_ISL_4250573\|2021-08-06 | 21A (Delta) |
| hCoV-19/South_Africa/Tygerberg_2478/2021\|EPI_ISL_4250419\|2021-08-02 | 21A (Delta) |
| hCoV-19/South_Africa/Tygerberg_2479/2021\|EPI_ISL_4250677\|2021-08-02 | 21A (Delta) |
| hCoV-19/South_Africa/Tygerberg_2480/2021\|EPI_ISL_4250583\|2021-08-02 | 21A (Delta) |
| hCoV-19/South_Africa/Tygerberg_2481/2021\|EPI_ISL_4250412\|2021-08-02 | 21A (Delta) |
| hCoV-19/South_Africa/Tygerberg_2482/2021\|EPI_ISL_4250516\|2021-08-02 | 21A (Delta) |
| hCoV-19/South_Africa/Tygerberg_2483/2021\|EPI_ISL_4250519\|2021-08-02 | 21A (Delta) |
| hCoV-19/South_Africa/Tygerberg_2484/2021\|EPI_ISL_4250641\|2021-08-02 | 21A (Delta) |
| hCoV-19/South_Africa/Tygerberg_2485/2021\|EPI_ISL_4250586\|2021-08-02 | 21A (Delta) |
| hCoV-19/South_Africa/Tygerberg_2486/2021\|EPI_ISL_4250721\|2021-08-02 | 21A (Delta) |
| hCoV-19/South_Africa/Tygerberg_2487/2021\|EPI_ISL_4250674\|2021-08-02 | 21A (Delta) |
| hCoV-19/South_Africa/Tygerberg_2488/2021\|EPI_ISL_4250451\|2021-08-05 | 21A (Delta) |
| hCoV-19/South_Africa/Tygerberg_2489/2021\|EPI_ISL_4250454\|2021-08-05 | 21A (Delta) |
| hCoV-19/South_Africa/Tygerberg_2491/2021\|EPI_ISL_4250590\|2021-08-04 | 21A (Delta) |
| hCoV-19/South_Africa/Tygerberg_2492/2021\|EPI_ISL_4250457\|2021-08-12 | 21A (Delta) |
| hCoV-19/South_Africa/Tygerberg_2493/2021\|EPI_ISL_4250402\|2021-08-06 | 21A (Delta) |
| hCoV-19/South_Africa/Tygerberg_2494/2021\|EPI_ISL_4250593\|2021-08-06 | 21A (Delta) |
| hCoV-19/South_Africa/Tygerberg_2495/2021\|EPI_ISL_4250738\|2021-08-08 | 21A (Delta) |
| hCoV-19/South_Africa/Tygerberg_2496/2021\|EPI_ISL_4250597\|2021-08-08 | 21A (Delta) |
| hCoV-19/South_Africa/Tygerberg_2497/2021\|EPI_ISL_4250460\|2021-08-10 | 21A (Delta) |
| hCoV-19/South_Africa/Tygerberg_2498/2021\|EPI_ISL_4250779\|2021-08-10 | 21A (Delta) |
| hCoV-19/South_Africa/Tygerberg_2499/2021\|EPI_ISL_4250654\|2021-08-10 | 21A (Delta) |
| hCoV-19/South_Africa/Tygerberg_2502/2021\|EPI_ISL_4250463\|2021-08-11 | 21A (Delta) |
| hCoV-19/South_Africa/Tygerberg_2503/2021\|EPI_ISL_4250607\|2021-08-14 | 21A (Delta) |
| hCoV-19/South_Africa/Tygerberg_2504/2021\|EPI_ISL_4250466\|2021-08-13 | 21A (Delta) |
| hCoV-19/South_Africa/Tygerberg_2505/2021\|EPI_ISL_4250529\|2021-08-16 | 21A (Delta) |
| hCoV-19/South_Africa/Tygerberg_2506/2021\|EPI_ISL_4250766\|2021-08-16 | 21A (Delta) |
| hCoV-19/South_Africa/Tygerberg_2507/2021\|EPI_ISL_4250618\|2021-08-16 | 21A (Delta) |
| hCoV-19/South_Africa/Tygerberg_2508/2021\|EPI_ISL_4250658\|2021-08-16 | 21A (Delta) |
| hCoV-19/South_Africa/Tygerberg_2509/2021\|EPI_ISL_4250470\|2021-08-14 | 21A (Delta) |
| hCoV-19/South_Africa/Tygerberg_2510/2021\|EPI_ISL_4250732\|2021-08-14 | 21A (Delta) |
| hCoV-19/South_Africa/Tygerberg_2512/2021\|EPI_ISL_4250680\|2021-08-14 | 21A (Delta) |
| hCoV-19/South_Africa/Tygerberg_2513/2021\|EPI_ISL_4250711\|2021-08-16 | 21A (Delta) |
| hCoV-19/South_Africa/Tygerberg_2515/2021\|EPI_ISL_4250736\|2021-08-16 | 21A (Delta) |
| hCoV-19/South_Africa/Tygerberg_2516/2021\|EPI_ISL_4250628\|2021-08-16 | 21A (Delta) |
| hCoV-19/South_Africa/Tygerberg_2517/2021\|EPI_ISL_4250483\|2021-08-18 | 21A (Delta) |
| hCoV-19/South_Africa/Tygerberg_2518/2021\|EPI_ISL_4250390\|2021-08-18 | 21A (Delta) |
| hCoV-19/South_Africa/Tygerberg_2519/2021\|EPI_ISL_4250769\|2021-08-17 | 21A (Delta) |
| hCoV-19/South_Africa/Tygerberg_2520/2021\|EPI_ISL_4250522\|2021-08-19 | 21A (Delta) |
| hCoV-19/South_Africa/Tygerberg_2521/2021\|EPI_ISL_4250495\|2021-08-17 | 21A (Delta) |
| hCoV-19/South_Africa/Tygerberg_2522/2021\|EPI_ISL_4250665\|2021-08-20 | 21A (Delta) |
| hCoV-19/South_Africa/Tygerberg_2523/2021\|EPI_ISL_4250690\|2021-08-20 | 21A (Delta) |
| hCoV-19/South_Africa/Tygerberg_2524/2021\|EPI_ISL_4250714\|2021-08-20 | 21A (Delta) |
| hCoV-19/South_Africa/Tygerberg_2525/2021\|EPI_ISL_4250398\|2021-08-29 | 21A (Delta) |
| hCoV-19/South_Africa/Tygerberg_2526/2021\|EPI_ISL_4250708\|2021-08-26 | 21A (Delta) |
| hCoV-19/South_Africa/Tygerberg_2529/2021\|EPI_ISL_4250782\|2021-08-30 | 21A (Delta) |

| | |
|---|---|
| hCoV-19/South_Africa/Tygerberg_2595/2021\|EPI_ISL_4498660\|2021-05-27 | 20H (Beta, V2) |
| hCoV-19/South_Africa/Tygerberg_2596/2021\|EPI_ISL_4498661\|2021-05-27 | 20H (Beta, V2) |
| hCoV-19/South_Africa/Tygerberg_2597/2021\|EPI_ISL_4498662\|2021-05-27 | 20H (Beta, V2) |
| hCoV-19/South_Africa/Tygerberg_2598/2021\|EPI_ISL_4498663\|2021-05-27 | 20H (Beta, V2) |
| hCoV-19/South_Africa/Tygerberg_487/2021\|EPI_ISL_1591359\|2021-01-01 | 20H (Beta, V2) |
| hCoV-19/South_Africa/Tygerberg_488/2021\|EPI_ISL_1608099\|2021-01-02 | 20H (Beta, V2) |
| hCoV-19/South_Africa/Tygerberg_531/2021\|EPI_ISL_912401\|2021-01-09 | 20I (Alpha, V1) |
| hCoV-19/South_Africa/Tygerberg_555/2021\|EPI_ISL_1591380\|2021-01-26 | 20H (Beta, V2) |
| hCoV-19/South_Africa/Tygerberg_595/2021\|EPI_ISL_1591396\|2021-02-05 | 20H (Beta, V2) |
| hCoV-19/South_Africa/Tygerberg_596/2021\|EPI_ISL_1591397\|2021-02-09 | 20H (Beta, V2) |
| hCoV-19/South_Africa/Tygerberg_597/2021\|EPI_ISL_1591398\|2021-02-09 | 20H (Beta, V2) |
| hCoV-19/South_Africa/Tygerberg_598/2021\|EPI_ISL_1591399\|2021-02-09 | 20H (Beta, V2) |
| hCoV-19/South_Africa/Tygerberg_599/2021\|EPI_ISL_1591400\|2021-02-09 | 20H (Beta, V2) |
| hCoV-19/South_Africa/Tygerberg_600/2021\|EPI_ISL_1591401\|2021-02-09 | 20H (Beta, V2) |
| hCoV-19/South_Africa/Tygerberg_601/2021\|EPI_ISL_1591402\|2021-02-09 | 20H (Beta, V2) |
| hCoV-19/South_Africa/Tygerberg_602/2021\|EPI_ISL_1591403\|2021-02-09 | 20H (Beta, V2) |
| hCoV-19/South_Africa/Tygerberg_610/2021\|EPI_ISL_1591410\|2021-02-01 | 20H (Beta, V2) |
| hCoV-19/South_Africa/Tygerberg_611/2021\|EPI_ISL_1608102\|2021-02-01 | 20H (Beta, V2) |
| hCoV-19/South_Africa/Tygerberg_613/2021\|EPI_ISL_1591412\|2021-02-01 | 20H (Beta, V2) |
| hCoV-19/South_Africa/Tygerberg_614/2021\|EPI_ISL_1591413\|2021-02-01 | 20H (Beta, V2) |
| hCoV-19/South_Africa/Tygerberg_615/2021\|EPI_ISL_1591414\|2021-02-01 | 20H (Beta, V2) |
| hCoV-19/South_Africa/Tygerberg_619/2021\|EPI_ISL_1591418\|2021-02-03 | 20H (Beta, V2) |
| hCoV-19/South_Africa/Tygerberg_620/2021\|EPI_ISL_1591419\|2021-02-03 | 20H (Beta, V2) |
| hCoV-19/South_Africa/Tygerberg_621/2021\|EPI_ISL_1591420\|2021-02-03 | 20H (Beta, V2) |
| hCoV-19/South_Africa/Tygerberg_622/2021\|EPI_ISL_1591421\|2021-02-03 | 20H (Beta, V2) |
| hCoV-19/South_Africa/Tygerberg_623/2021\|EPI_ISL_1591422\|2021-02-03 | 20H (Beta, V2) |
| hCoV-19/South_Africa/Tygerberg_624/2021\|EPI_ISL_1591423\|2021-02-04 | 20H (Beta, V2) |
| hCoV-19/South_Africa/Tygerberg_625/2021\|EPI_ISL_1591424\|2021-02-04 | 20H (Beta, V2) |
| hCoV-19/South_Africa/Tygerberg_626/2021\|EPI_ISL_1591425\|2021-02-04 | 20H (Beta, V2) |
| hCoV-19/South_Africa/Tygerberg_629/2021\|EPI_ISL_1591428\|2021-02-10 | 20H (Beta, V2) |
| hCoV-19/South_Africa/Tygerberg_630/2021\|EPI_ISL_1591429\|2021-02-10 | 20H (Beta, V2) |
| hCoV-19/South_Africa/Tygerberg_634/2021\|EPI_ISL_1591433\|2021-02-11 | 20H (Beta, V2) |
| hCoV-19/South_Africa/Tygerberg_635/2021\|EPI_ISL_1591434\|2021-02-11 | 20H (Beta, V2) |
| hCoV-19/South_Africa/Tygerberg_639/2021\|EPI_ISL_1591438\|2021-02-12 | 20H (Beta, V2) |
| hCoV-19/South_Africa/Tygerberg_640/2021\|EPI_ISL_1591439\|2021-02-12 | 20H (Beta, V2) |
| hCoV-19/South_Africa/Tygerberg_642/2021\|EPI_ISL_1591441\|2021-02-12 | 20H (Beta, V2) |
| hCoV-19/South_Africa/Tygerberg_643/2021\|EPI_ISL_1591442\|2021-02-12 | 20H (Beta, V2) |
| hCoV-19/South_Africa/Tygerberg_644/2021\|EPI_ISL_1591443\|2021-02-12 | 20H (Beta, V2) |
| hCoV-19/South_Africa/Tygerberg_976/2021\|EPI_ISL_3066524\|2021-05-03 | 20H (Beta, V2) |
| hCoV-19/South_Africa/VIDA-KRISP-K010665/2021\|EPI_ISL_2360431\|2021-01-06 | 20H (Beta, V2) |
| hCoV-19/South_Africa/VIDA-KRISP-K010666/2021\|EPI_ISL_2360432\|2021-01-15 | 20H (Beta, V2) |
| hCoV-19/South_Africa/VIDA-KRISP-K010675/2021\|EPI_ISL_2360439\|2021-01-06 | 20H (Beta, V2) |
| hCoV-19/South_Africa/VIDA-KRISP-K010676/2021\|EPI_ISL_2360440\|2021-01-20 | 20H (Beta, V2) |
| hCoV-19/South_Africa/VIDA-KRISP-K010677/2021\|EPI_ISL_2360441\|2021-01-06 | 20H (Beta, V2) |
| hCoV-19/South_Africa/VIDA-KRISP-K010678/2021\|EPI_ISL_2360442\|2021-01-06 | 20H (Beta, V2) |
| hCoV-19/South_Africa/VIDA-KRISP-K010683/2021\|EPI_ISL_2360446\|2021-01-13 | 20H (Beta, V2) |
| hCoV-19/South_Africa/VIDA-KRISP-K010686/2021\|EPI_ISL_2360448\|2021-01-07 | 20H (Beta, V2) |
| hCoV-19/South_Africa/VIDA-KRISP-K011193/2021\|EPI_ISL_2360559\|2021-01-12 | 20H (Beta, V2) |
| hCoV-19/South_Africa/VIDA-KRISP-K011238/2021\|EPI_ISL_2360600\|2021-02-02 | 20H (Beta, V2) |
| hCoV-19/South_Africa/VIDA-KRISP-K011240/2021\|EPI_ISL_2360602\|2021-02-02 | 20H (Beta, V2) |

| | |
|---|---|
| hCoV-19/South_Africa/VIDA-KRISP-K011244/2021\|EPI_ISL_2360606\|2021-01-06 | 20H (Beta, V2) |
| hCoV-19/South_Africa/VIDA-KRISP-K011245/2021\|EPI_ISL_2360607\|2021-01-15 | 20H (Beta, V2) |
| hCoV-19/South_Africa/VIDA-KRISP-K011254/2021\|EPI_ISL_2360616\|2021-02-02 | 20H (Beta, V2) |
| hCoV-19/South_Africa/VIDA-KRISP-K011329/2021\|EPI_ISL_2360683\|2021-01-07 | 20H (Beta, V2) |
| hCoV-19/South_Africa/VIDA-KRISP-K011403/2021\|EPI_ISL_2360718\|2021-02-08 | 20H (Beta, V2) |
| hCoV-19/South_Africa/VIDA-KRISP-K011409/2021\|EPI_ISL_2360724\|2021-01-22 | 20C |
| hCoV-19/South_Africa/VIDA-KRISP-K011410/2021\|EPI_ISL_2360725\|2021-01-22 | 20H (Beta, V2) |
| hCoV-19/South_Africa/VIDA-KRISP-K011411/2021\|EPI_ISL_2360726\|2021-01-22 | 20H (Beta, V2) |
| hCoV-19/South_Africa/VIDA-KRISP-K011476/2021\|EPI_ISL_2360786\|2021-02-05 | 20H (Beta, V2) |
| hCoV-19/South_Africa/VIDA-KRISP-K011479/2021\|EPI_ISL_2360788\|2021-02-17 | 20H (Beta, V2) |
| hCoV-19/South_Africa/VIDA-KRISP-K012159/2021\|EPI_ISL_2494486\|2021-01-19 | 20H (Beta, V2) |
| hCoV-19/South_Africa/VIDA-KRISP-K012472/2021\|EPI_ISL_2494519\|2021-02-10 | 20H (Beta, V2) |
| hCoV-19/South_Africa/VIDA-KRISP-K012615/2021\|EPI_ISL_2494414\|2021-02-10 | 20H (Beta, V2) |
| hCoV-19/South_Africa/VIDA-KRISP-V001005/2021\|EPI_ISL_940864\|2021-01-08 | 20H (Beta, V2) |
